# Dynamic analysis of the individual patterns of intakes, voids, and bladder sensations reported in bladder diaries collected in the LURN study

**DOI:** 10.1101/2023.04.05.23288100

**Authors:** Victor P. Andreev, Margaret E. Helmuth, Abigail R. Smith, Anna Zisman, Anne P. Cameron, John O. L. DeLancey, Wade A. Bushman

## Abstract

The goal of this study was to perform an in-depth dynamic analysis of individual bladder diaries to inform which behavioral modifications would best reduce lower urinary tract symptoms, such as frequency and urgency. Three-day bladder diaries containing data on timing, volumes, and types of fluid intake, as well as timing, volumes, and bladder sensation at voids were analyzed for 197 participants with lower urinary tract symptoms. A novel dynamic analytic approach to bladder diary time series data was proposed and developed, including intra-subject correlations between time-varying variables: rates of intake, bladder filling rate, and urge growth rate. Grey-box models of bladder filling rate and multivariable linear regression models of urge growth rate were developed for individual diaries. These models revealed that bladder filling rate, rather than urine volume, was the primary determinant of urinary frequency and urgency growth rate in the majority of participants. Simulations performed with the developed models predicted that the most beneficial behavioral modifications to reduce the number of urgency episodes are those that smooth profiles of bladder filling rate, which might include behaviors such as exclusion of caffeine and alcohol and/or other measures, e.g., increasing number and decreasing volumes of intakes.

## Introduction

Bladder diaries (BDs) are a useful tool for diagnosis and treatment of patients with lower urinary tract symptoms (LUTS). Studies have shown that BDs provide accurate data on functional bladder capacity [1], incontinence episodes, frequency, nocturia, and daily urgency [2-6], and correlate well with symptom scores and urodynamics [7-8]. BDs can also provide timing and volumes of fluid intake, which is the primary driver of urinary output. The time lapse between intake and output is not straightforward. Although intake and output ultimately equilibrate over time, there is significant variability between individuals in the timing of output. Commonly used metrics from BDs ignore the timing of voids and tend to focus on summary statistics of voided volume (VV) and void frequency; however, inopportune timing of the urgent need to void can be bothersome to patients.

Mathematical modeling can be used to leverage the rich time series data available in BDs. There are several mathematical models analyzing renal physiology and urine production, including neural control of kidney function [9-16]. These models provide helpful insights for understanding physiology of urination; however, they do not allow for the prediction of adverse urinary events or identification of behavioral modifications to avoid such events.

The goal of this paper is to fill this gap by introducing a dynamic approach to the analysis of BDs, complementary to existing approaches that average dynamics of fluid intake and voiding over time. We investigate how drinking patterns and types of fluid intake affect the rate at which urine is produced by the kidneys. We also investigate how bladder filling rate (BFR) affects bladder sensations. Unlike the time-averaged approach, we are interested not only in the number of voids per day and how it correlates with the total volume of the intake, but also in how the time interval between voids and the bladder sensations at voids change during the day and how they associate with timing, volumes, and composition of fluid consumed. Since one of the goals of the paper is to introduce and justify the new analytic approach, the Methods section is rather detailed and includes not only the description of, but also the thought process for selection of proposed methods.

## Materials and Methods

### Data

Data were obtained from the Symptoms of Lower Urinary Tract Dysfunction Research Network (LURN) Observational Cohort study, which collected self-reported urinary symptoms, BDs, and physical examination data on 1064 care-seeking female and male participants across six tertiary care centers [17-18]. Three-day BDs were collected using a modified International Consultation on Incontinence Questionnaire (ICIQ) bladder diary [19]. Details of the study and quality of the data have been reported [20]. Diaries included information on the timing and volumes of fluid intake and voids, as well as bladder sensations during voiding and types of fluids consumed, across 3 consecutive days.

Briefly, 448 participants returned BDs that had data on 3 days, had no missing intake and void volumes, and had a physiologically plausible fluid imbalance (<3 L across all 3 days). The dynamic analysis approach of this paper requires detailed data; therefore, we additionally excluded BDs that had missing voiding, drinking, waking, and sleeping times, types of fluid consumed, and bladder sensations. We did not attempt to impute these missing data, but limited ourselves to the analysis of complete BDs, since imputations inherently add variability, and since we had enough complete cases. We also excluded participants with post-void residual (PVR) >50 mL. This resulted in 197 BDs suitable for dynamic analysis belonging to 99 male and 98 female participants. Of these, 74 had at least one incontinence episode in 3 days, 165 consumed caffeinated drinks, and 65 consumed drinks containing alcohol.

Reported information on the types of fluid consumed was used to estimate osmolality, caffeine, and alcohol content of the fluid based on literature review [21-22]. Estimates for durations of fluid intake (time taken to drink the fluid) were based on expert opinion of the authors who independently reviewed a representative list of beverages and produced estimates of time for consumption for each type of beverage. Estimated durations varied from 1 minute to 20 minutes (hot beverages and alcoholic beverages are usually consumed slower than water, for example). Bladder sensations were reported on a rating scale from 0 to 4, with 0 meaning “no sensation of needing to pass urine, but passed urine for social reasons; 1=“normal desire to pass urine and no urgency”; 2=“had urgency, but it had passed before patient went to the toilet”; 3=“had urgency but managed to get to the toilet, still with urgency, but did not leak urine”, 4=“had urgency and could not get to the toilet in time so leaked urine” [19]. The 197 3-day BD reported a total of 5124 voids. Of these, 342 were with urge level=0; 2341 with urge level=1; 732 with urge level=2; 1287 with urge level=3; and 422 with urge level=4, i.e., participants experienced sensation of urgency during 47.6% of voids, normal desire to pass urine in 45.7% of voids, and in 6.7% cases voided for social reasons.

### Ethical guidelines and consent

As described in the Data section above, this is a secondary data analysis of LURN bladder diaries. The authors confirm all relevant ethical guidelines have been followed, and all research has been conducted according to the principles expressed in the Declaration of Helsinki. Written informed consent was obtained from participants enrolled in the LURN study from 2015 to 2017, with each participant signing a confidential consent form witnessed and signed by the site research coordinator. This paper’s authors did not have access to information that could identify individual participants during or after data collection. Institutional Review Board (IRB) approval was obtained from: Ethical and Independent Review Services (E&I) IRB, an Association for the Accreditation of Human Research Protection Programs (AAHRPP) Accredited Board, Registration #IRB 00007807.

### Methods

#### Overview of the multistep analysis of BDs

First, we performed a traditional time-averaged analysis of the BDs. We examined the distributions and inter-subject correlations of demographic and time-averaged BD variables across the cohort of 197 participants with LUTS. Then, we introduced a dynamic approach to the analysis of BDs by examining intra-subject correlations between BD variables across the 3-day duration of the diaries, investigated cross-correlation functions and delays between these variables, and finally developed and evaluated dynamic models for individual BDs (Figure 1).

**Figure 1.**
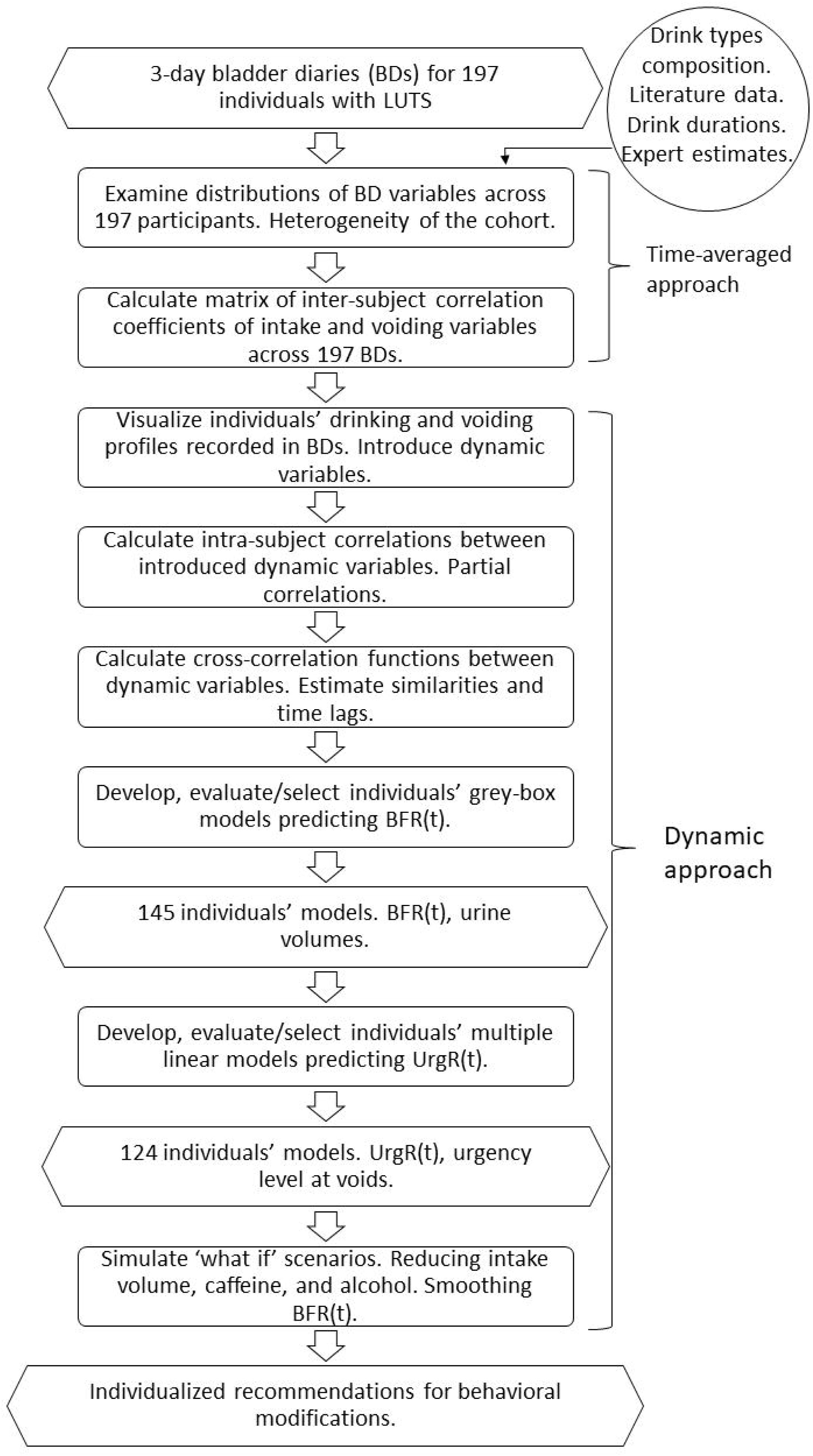
Flowchart of the multistep analysis of BDs. Analysis steps are in rectangles, data and results are in the hexagons. Steps 1-2 constitute time-averaged analysis; steps 3-8 constitute a dynamic approach. First, we examined the distributions of BD variables across 197 participants. Second, we performed the time-averaged analysis of BDs by examining the matrix of inter-subject correlation coefficients of intake and voiding variables across 197 BDs. Third, we visualized individual BD by presenting graphs of drinking and voiding events and introduced dynamic variables describing drinking and voiding patterns. Fourth, we examined the intra­subject correlations between the introduced dynamic variables. Fifth, we calculated cross­correlation functions between the introduced dynamic variables to estimate both their similarities and time lags. Sixth, we developed and evaluated the individuals’ grey-box models predicting BFR using data on intake profiles and composition of the drinks. Seventh, we developed multivariable linear regression models for each individual urge growth rate (UrgR). Finally, we proposed behavioral modifications, i.e., restrictions of intake volume, caffeine, and alcohol consumption and simulated the effect of these modifications on UrgR and urge levels at voids.

#### Justification of dynamic approach

A dynamic approach to understanding reality, i.e., description of relationships between changes in variables in time rather than relationships between the values of the variables, proves to be productive in quantitative sciences, starting with Newton’s laws of motion, where acceleration, i.e., change in velocity of the object, was shown to be proportional to the force acting on the object. We propose to use a dynamic approach to the analysis of BDs, by exploring how changes in blood volume, osmolality, and concentrations of caffeine and alcohol, caused by fluid consumption affect (change) the rate of urine production by the kidneys. We propose to investigate how the sensation of urinary urge changes in time and how it is affected by the bladder filling rate and urine volume. Using this dynamic approach, we introduce urge growth rate (UrgR) as defined in the next section. We believe that UrgR often (with the exception of social or convenience voids) drives voiding behavior. For example, time interval between voids is determined by the time required to reach certain urge levels, and therefore is proportional to the reciprocal of the UrgR. In a sense, UrgR is more important than urge level at void. Any healthy person deprived of the opportunity to void will eventually get to a high level of urge, e.g., urge=3; however, it will likely take a healthy person much longer to get to this level than a person with overactive bladder (OAB). This makes UrgR a useful metric to evaluate the severity of OAB.

While implementing this dynamic approach to the analysis and modeling of BDs and to the simulation of potential behavioral modifications, we kept in mind that information available for modeling, as well as our understanding of the involved physiological processes, are incomplete. For instance, were have no information on physical activities or changes in heart rate that might affect the urine production rate by the kidney. We do not have food diaries and therefore do not have information on the fluids consumed in food (e.g., watermelon or soup). We do not know how much salt is consumed with food and when, which influences water retention. We do not know about the presence of psychological distractions and triggers that might influence the sensation of urge. All this incompleteness of information makes our task of predicting urgency episodes less like Newtonian mechanics calculation of the position of planets and more like the task of the captain determining the course of the ship having an approximate knowledge of the strength of the winds and currents, and therefore regularly measuring the position of the ship relative to the sun and the stars or using global positioning system (GPS) signals. Similarly, we try to minimize the propagation of the errors along the modeling process and, whenever possible, use the measured values of variables recorded in the BDs.

#### Dynamic variables

BDs report the values of intake variables (volumes and types of drinks) at times of intakes and voiding variables (volumes and urge levels) at times of voids, which do not coincide. Dynamic analysis requires the knowledge of intake variables and voiding variables at the same and preferably equidistant time points. To meet this requirement and to analyze dynamics of drinking and voiding patterns and bladder sensations, we defined several dynamic (time­dependent) variables across the whole duration of the 3-day BDs. We calculated rate of intake, bladder filling rate (BFR), urge growth rate (UrgR), and time-dependent frequency of voiding (F), as shown below. Stepwise approximations were used to derive these variables from the BD data. They were considered constant during the time intervals (durations of fluid intake and time intervals between voids), with changes occurring only at the boundaries of these intervals.

Fluid intake rate (*IR(t))* was approximated as i^th^ intake volume (*IV_i_*) divided by the estimated duration of i^th^ intake (*Di*) and assumed zero everywhere except during reported intake, where i=1,..M, M-number of reported intakes. No fluid intake was assumed or attributed to the food the participant ate; only fluids recorded on the BD were included in the analysis.

Frequency of voiding (*F(t))* was defined as:

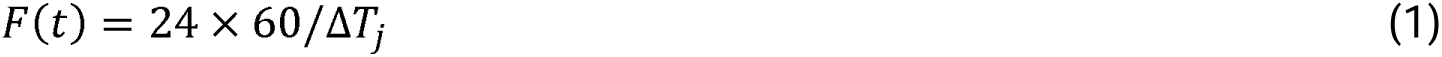

where Δ*T_j_ = T_j_ – T_j_*_–1_, time interval from previous void in minutes, *T_j_*, j=1,..N – time of each of N voids reported by an individual. For easier interpretation, the 24 × 60 multiplier was introduced to make frequency defined by eq. 1 comparable with the averaged daily frequency, i.e., number of voids per day. Note that F defined by eq. 1 depends on time, unlike the frequency of voiding averaged across the day or several days typically used in the urologic literature [23,24]. For example, consider an individual who voided at 8:00 am, 9:00 am, 9:20 am, 9:40 am, 12:00 noon, 2:00 pm, 5:00 pm, 7:00 pm, 8:00 pm, and 11:00 pm. They would typically be described as voiding 10 times a day, or voiding every 90 minutes; however, there was a time during the day when they voided every 20 minutes, which is not reflected by the time-averaged definition of frequency, but is reflected by the peak (*F(t=9:20 am)=72*) of the time-dependent frequency (eq. 1). Note that the presence of such high-frequency voids at inopportune times could be much more bothersome for the individual than voiding 11 versus 10 times a day; therefore, it is desirable to have tools to predict such high-frequency episodes, as well as high-urgency episodes.

Urge was assumed to be zero immediately after each void, so *UrgR(t)* was defined as urge *(U_j_*) reported at the *j^th^* void divided by *ΔT_j_,* assuming a linear growth of urge with time, which may not always be correct but is close to sigmoidal growth observed in several studies measuring real-time bladder sensations in individuals with and without OAB [25,26]. Even when the assumption of linear growth of urge is not valid, the variable *UrgR(t))* defined as *U_j_*/Δ*T_j_* is of importance since it reflects how much time it takes to reach the certain level of urge. The time required to reach urgency is even more important than level of urge. For instance, an individual without OAB driving on the highway (with limited access to a toilet) might get a sensation of urge *U_j_* =3 and need to exit the highway. However, an individual with OAB will get to urge *U_j_* =3 much faster and would need to exit more often. Variable *U_j_*/Δ*T_j_* is different from frequency defined by eq. 1, since some of the voids could be social or convenience ones and occur at the low level of *U_j_*.

The BFR, sometimes referred to as the rate of diuresis [27,28], was defined as: *BFR*(*t*) *= VV_j_*/Δ*T_j_*, where *VV_j_* is the *j^th^* voided volume (VV). Here, we assumed that the variation of PVR from void to void is small relative to the VV (justified in participants with PVR<50mL selected for this analysis); therefore, the VV can approximate the change of the urine stored in the bladder between voids.

Figure 2 illustrates the 3-day BD data (intake and VVs, and urge levels reported at voids) and its relationship with the dynamic variables derived from these data (BFR, time-dependent (F), and UrgR) for a typical study participant A. Supplemental Figures S1-S9 provide similar information for nine other representative participants (B-J). More detailed discussion of the figures is provided in the Results section; however, we think it is beneficial to introduce these figures here for better illustration of our dynamic approach to the BD analysis.

**Figure 2.**
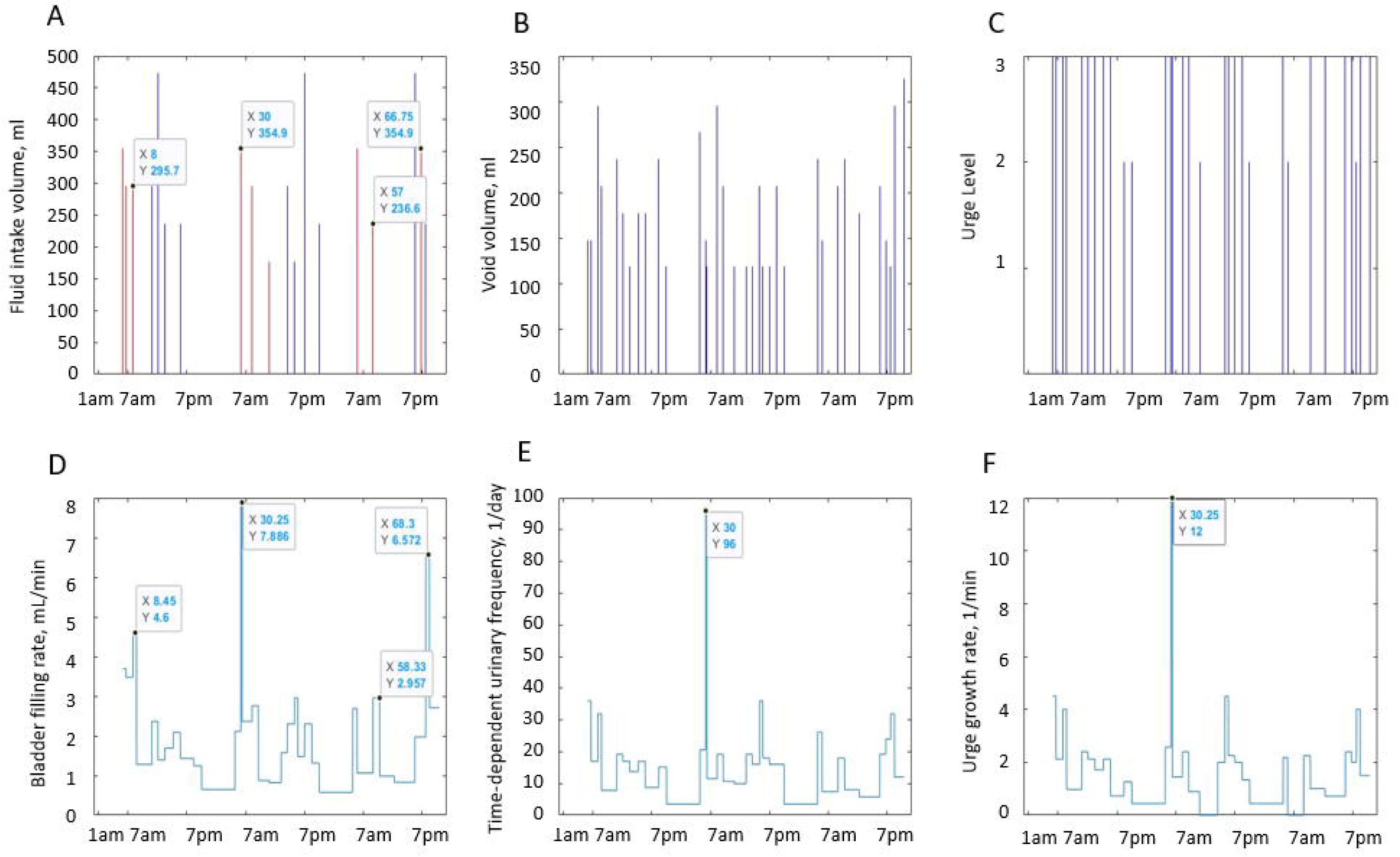
Intake and voiding profiles recorded in the 3-day BD of patient A and dynamic variables derived from the profiles. Figure 2A: fluid intake volumes. Figure 2B: voided volumes (VV). Figure 2C: urge levels reported at voids. Data in Figures 2A-2C are presented in the form of stems instead of dots to illustrate the discrete nature of drinking and voiding events and to emphasize the impossibility of connecting the dots, e.g., intake volumes are equal to zero between intakes, and void volumes are equal to zero between voids. Figure 2D: bladder filling rate (BFR(t)). Figure 2E: time-dependent frequency of voiding (F(t)). Figure 2F: urge growth rate (UrgR(t)). Caffeine containing intakes are shown in Figure 2A in red. Note that peaks of BFR(t), F(t), and UrgR(t) are collocated with intakes of caffeine and with frequent and/or high-volume drinks. Some of these peaks are labeled to emphasize their collocation (X=time in hours from midnight of the first day of the diary; Y=values of the plotted variables). Note high (several fold) time variability of BFR.

#### Inter-subject and intra-subject correlations

Both inter-subject and intra-subject correlation coefficients of BD variables were calculated. The inter-subject correlations analysis was included in the time-averaged approach (step 2 in Figure 1). It was performed by first averaging the dynamic BD variables over time (3 days of the BD) and then calculating the matrix of the Pearson correlation coefficients of the time-averaged variables across 197 BDs. The intra-subject correlation analysis was part of dynamic approach (step 4 of Figure 1). It was performed by calculating matrices of Pearson correlation coefficients of dynamic variables for each of the individuals and then averaging the individual correlation matrices across all 197 individuals. Matrices of Pearson correlation coefficients were calculated using the MATLAB function corrcoef.m (MathWorks, MA).

Both for inter-subject and intra-subject correlations, we also calculated matrices of partial correlation coefficients, while controlling for certain variables. This type of analysis helps when there are three or more codependent variables (e.g., A, B, C), and we want to know how variable A responds to the changes in variable B, which are not caused by the changes in variable C. It is similar to controlling for variables in linear regression models [29] and was implemented using the MATLAB function partialcorr.m.

#### Cross-correlation function

Reactions of organisms to changes in external factors are not instantaneous but take time, which may be short or long depending on the physiological processes involved and may differ across individuals. For instance, it may take different time in different individuals for the BFR to react to the increase in the intake due to consumed fluid. Intra-subject correlations do not allow for capturing similarities in the presence of unknown time delays between the variables. To evaluate similarities and time lags between the functions representing the dynamic variables, we used cross-correlation function, which is routinely used to compare signals in signal processing [30] and is defined as:

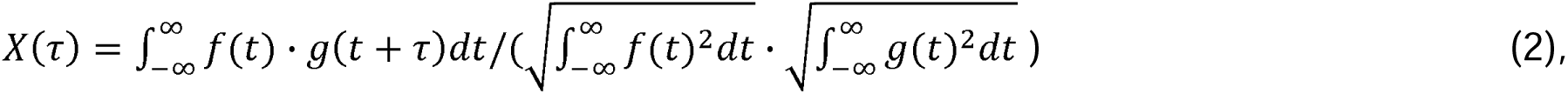

where *f*(*t*) and *g*(*t*) are two dynamic variables under comparison. The maximum possible value of cross-correlation function is *X*(*τ_max_*) = 1, which happens when *g*(*t + τ_max_*) *= f*(*t*). Therefore, the value of *X*(*τ_max_*) represents the level of similarity, while *τ_max_* represents the time lag between the two functions. We used cross-correlation function to compare the dynamic variables derived from the BD data. We also used cross-correlation function to compare BFR profiles predicted by the developed models and derived from the BDs. Cross-correlation function was implemented as MATLAB function xcorr.m.

#### Grey-box model of the urine formation rate

We created mathematical models for each individual participant, describing the dynamics of the BFR, which is equal to the rate of urine formation by the kidneys. In doing so, we followed a nonlinear grey-box modeling approach. Grey-box model is a type of model where the equations describing the processes are assumed known, while the parameters/coefficients of the equations have physical or physiological meaning and are determined by minimizing differences between observed and modeled processes. We described the model by a set of nonlinear ordinary differential equations (ODEs), with free parameters determined through minimization of the difference between urine formation rate profile predicted by the model and BFR profile derived from the BD. We assumed that the structure of the model was the same for all participants, while the parameters of the models were participant-specific. The core of our model is the modified model of Bighamian et al [31] describing the dynamics of the distribution of added water between blood and interstitial fluid, which is essential for determination of time lags between intakes and voids. We generalized this model by adding two more compartments (stomach and intestine) and by describing transport, kinetics, and redistribution of electrolytes, caffeine, and alcohol between blood and interstitial fluid. We also allowed for urine formation rate dependency on blood plasma volume, osmolality, and caffeine and alcohol concentrations. The model was implemented with System Identification Toolbox (MATLAB 2021a), using function idnlgrey.m. Details on the assumptions, equations, and implementation of the grey-box models of urine formation rate are presented in the Supplemental Material.

#### Comparing modeled and observed dynamic variables by evaluating locations and amplitudes of the peaks

The traditional metric to compare modeled and observed variables is called “fit percent” and is calculated as:

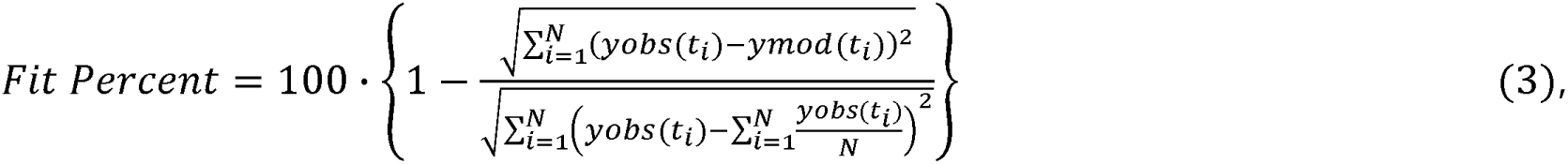

where *yobs*(*t_i_*) and *ymod*(*t_i_*) are the values of observed and modeled dynamic variable *y* at all points *t_i_,* where this variable was observed and modeled. Note that this metric compares differences in the observed and modeled values of the variable (numerator) with the overall time-variability of the observed variable (denominator), which makes it, in general, suitable for evaluating dynamic models. However, given the presence of high and narrow peaks in the variables of interest in the BDs (see BFR(t), F(t) and UrgR(t) in Figure 2 and Supplemental Figures S1-S9), which could be caused by high-volume or/and high caffeine content intakes, this metric might be unsatisfactory. For instance, it could happen that narrow peaks in *yobs*(*t_i obs_*) and *ymod*(*t_i mod_*) are closely collocated but not overlapping. In this case, the *Fit Percent* value would be lower than if the peak of interest was completely absent in the modeled function, which would misrepresent properties of the model. To avoid this situation, we introduced a new metric named *Peak Fit*, similar to *Fit Percent*, but allowing for some lags in the peaks of the modeled function; i.e., we identified ten highest observed and modeled peaks and searched for the modeled and observed maxima in the specified vicinity (30 minutes) of these peaks. The below equation for *Peak Fit* is similar to equation for *Fit Percent* but uses only the peak apex points compared with the maxima in the vicinity of the peaks. This way, it allows for limited lags in the modeled function relative to the observed function and punishes both the cases of missing peaks and of false modeled peaks, which are absent in the observed function.

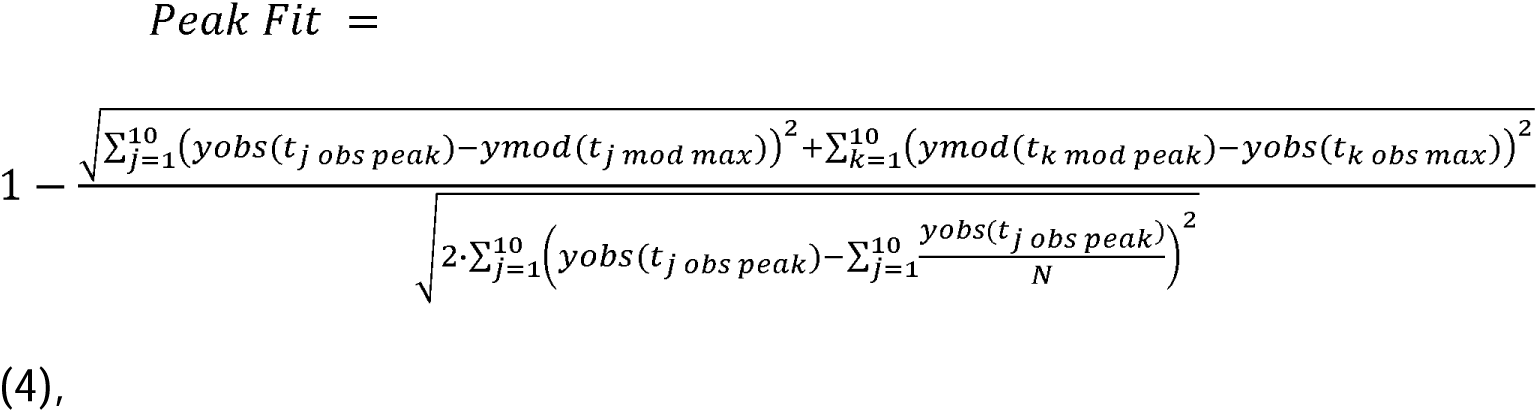

where *t_j obs peak_* and *t_k mod peak_* are positions of the apexes of the peaks in the observed and modeled functions, while *t_j mod max_* and *t_k obs max_* are the maximum values of modeled and observed functions in the vicinity of the above peaks.

To evaluate the average lag between observed and modeled peaks, we introduced another metric:

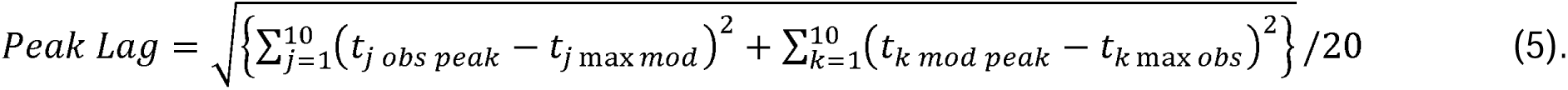

Note that this metric is similar to the *τ_max_*-lag provided by the cross-correlation function; however, the latter is dominated by the lag of the highest peak, while the former provides lag averaged across multiple peaks.

#### Evaluation and selection of grey-box models

An important quality of any model is its ability to describe/predict observations in the time range outside of the time range used for parameter fitting. To estimate this ability, we derived not only the models using 3 days of BD data (3-day model), but also the models using only first 2 days of BD data (2-day model). Then, we used the parameters of the 2-day models and 3-day intake data to predict BFR(t) during all 3 days. Details of the comparison of 2-day and 3-day models are provided in the Results section and are briefly summarized below. We evaluated the developed 197 individuals’ grey-box models of urine production rate by using the *Peak Fit* metric defined by eq. 4. We selected for further analysis and use only the individuals with *Peak Fit>0.9* both for 3-day and 2-day grey box models of urine production rate. Models for 145 individuals satisfied these criteria. The mean value of Peak Fit for these models of BFR(t) was 0.94, and the mean value of Peak Lag was 6 minutes. We also compared the “full models”, including information on caffeine and alcohol content of the intakes, with the “no caffeine no alcohol” models ignoring this information. It was shown that “full models” provide substantially better fit.

#### Regression models of UrgR

Physiology of bladder sensations is less clear than that of urine formation by the kidneys; therefore, creation of the grey-box model of UrgR does not seem feasible. It is known, however, that feeling of urinary urge is initiated by stretching of the bladder wall [32]. We assumed that the reaction of the nervous system to wall stretching was nearly instantaneous, or at least happened at a much shorter time scale (seconds) than the time scale of the BD records (time intervals between drinks and voids – minutes) and the typical time required to establish equilibrium distribution of water between blood and interstitial fluid (minutes or hours). Given the assumption of the nearly instantaneous reaction of the nervous system to the input variables, we used the multiple linear regression approach to create personalized models of urinary UrgR for each participant. Analysis of the intra-subject correlations performed as described in the “Inter-subject and intra-subject correlations” subsection above and in the Results section below provided evidence of different levels of correlations of UrgR with BFR and with VV, which correspond to sensitivity, not only to the level of bladder wall stretch, but also to the velocity of stretching.

Given these observations, the inputs of the model included BFR (*x_1_*=BFR(t)) and volume (x_2_=V(t)) of the urine in the bladder (calculated as integral of the BFR from the time of previous void to the given moment plus PVR volume 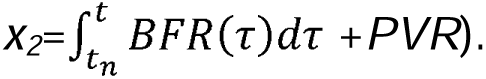 The last input (*x_3_*) was binary and contained information on whether the participant was awake or asleep at the given moment, which allowed for possible bladder sensation differences in these two states. The formula for model specification can be presented as:

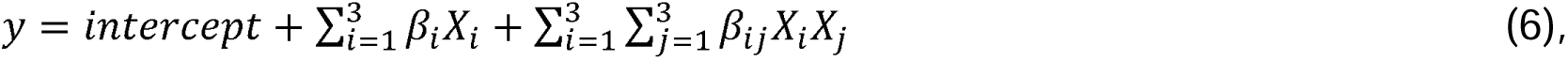

where *y* = *UrgR(t),* while 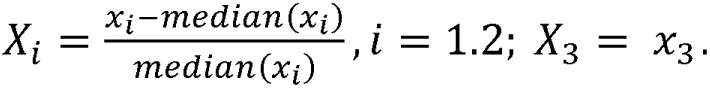 Scaling of the variables is not necessary in multivariable linear regression but was implemented for further comparison of the relative role of different input variables in predicting UrgR. For instance, intercept in this formulation represents the value of UrgR in the hypothetical case when all the input variables are kept constant during the 3-day BD and equal to the median values of these variables during these 3 days. The values of coefficients *β*_1_*, β*_2_ represent the increase in the UrgR in response to the change in the values of input variables equal to their medians. The above specification (eq 6) means that the model allows not only for linear dependence of UrgR(t) on the input terms, but also on their interactions. The values of UrgR(t) and BFR(t) were determined from the BDs and assumed constant during the time intervals between voids; asleep/awake data was also taken directly from the BDs. This method was chosen to describe the inputs of the model as accurately as possible. BFR (t) can be determined from the grey-box model, however, less accurately than from the BDs since the model does not take into account the changes in heart rate or the consumption of salted food or diuretics, which could contribute to changes in BFR observed from BDs.

Multivariable linear regression was performed using MATLAB function stepwiseglm.m, which uses forward and backward stepwise regression to determine the values of coefficients *β_i_* and *β_¡j_*. At each step, the function adds or removes terms to the model, based on the value of the ‘criterion’ chosen. In this situation, the default value of ‘criterion’ was used, i.e., the p-value for an F-test of the change in the deviance that results from adding or removing the term. The quality of the individual final models was evaluated by their adjusted R squared value (Adj. R^2^). Only the models with Adj. R^2^ >0.3 were selected for further analysis and simulation.

#### Calculating urge level using the predicted UrgR: Predicting urgency episodes

Timings of voids are not completely determined by the intake information, since some of the voids are “social” or “convenience” voids, which occur without urge or at the low level of urge and cannot be predicted by renal physiology-based models. Urge sensation is supposed to be zero at the moments right after voids; therefore, timings of voids are necessary as independent data to determine the level of urge with UrgR predicted by the developed regression models. Therefore, we used BD data on the timings of voids (*t_i_*) together with the predicted by the regression models UrgR (*pUrgR(t))* to predict the values of urge levels at the times of voids:

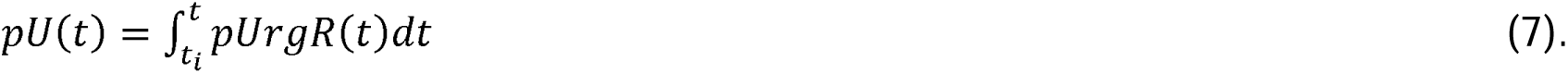

Note that *pUrgR*(*t*) predicted by the model (unlike UrgR estimated from the BD) is not a necessary constant in the interval between the adjacent voids; therefore, integration rather than multiplication by the time interval is warranted. We then used eq. 7 to calculate urge levels at the time of each void and evaluated percentage of true positive and false positive predictions of urge episodes by comparing with urge levels reported in BD. Note that, while observed/reported values of urge at the moments of void have discrete integer values 0,1,2,3,4, the predicted values of urge pU(t_i_) can have any positive value. Given the previously-outlined definitions of urge levels [19], we counted all voids where U(t_i_)=2,3, or 4 as urgency episodes. In comparison, for the predicted urgency episodes, we implemented a “personalized urgency threshold” approach by allowing the threshold level for each patient to change from 1.05 to 2 and selecting for each patient the threshold that minimized the sum of percentages of false negatives and false positives. This is equivalent to adding one more parameter to the individual’s model of sensation of urge.

#### Simulation of “what if’ scenarios and personalized behavioral modifications

We used the developed models to simulate personalized behavioral modifications aimed to avoid or minimize urgency episodes. Simulation is different from model development since it does not involve parameter fitting. Parameters of each individual model were already determined at the stage of model development. In simulation, parameters of the models are fixed, inputs of the models are modified, and the changes of the outputs caused by the modifications of the inputs are reported. Simulating modifications of intake patterns involve running with the modified intake data the developed individual grey-box models of urine production and the regression models of UrgR. In particular, some of the simulated modifications excluded use of alcohol and/or caffeinated drinks, substituting them with the alcohol-free and decaffeinated drinks with the same osmolality and volume. The BFR(t) profiles with proposed modified inputs and with the initial true inputs are calculated using the grey-box models; the difference in two profiles is determined and added to the BFR(t) profile determined from the BD. The result constitutes our best guess of the BFR(t) profile and serves as the first input variable for the developed multivariable linear regression model of UrgR(t), while the second input variable is determined by the time integral of the first one. Note that these simulations are different from “no caffeine no alcohol” models discussed above. While the comparison of “no caffeine no alcohol models” with “full models” served to demonstrate the importance of considering the effect of caffeine and alcohol on BFR for achieving adequate fit of modeled and observed profiles, the simulations with excluded caffeine and alcohol containing beverages served to examine if abstinence of these beverages could help to avoid urgency episodes.

## Results

### Time-averaged analysis of BD data

#### Distributions of the BD variables across 197 participants

Our cohort of 197 participants with BDs suitable for dynamic analysis is quite heterogeneous. There is substantial variability across the cohort by age, body mass index (BMI), 3-day intake volumes, mean intake volumes, types of drinks (osmolality, caffeine and alcohol content), voided volumes (VVs), bladder capacity (estimated as maximum VV), urge levels, and numbers of leaks, as illustrated by Table 1 and histograms in Figure 3.

**Figure 3.**
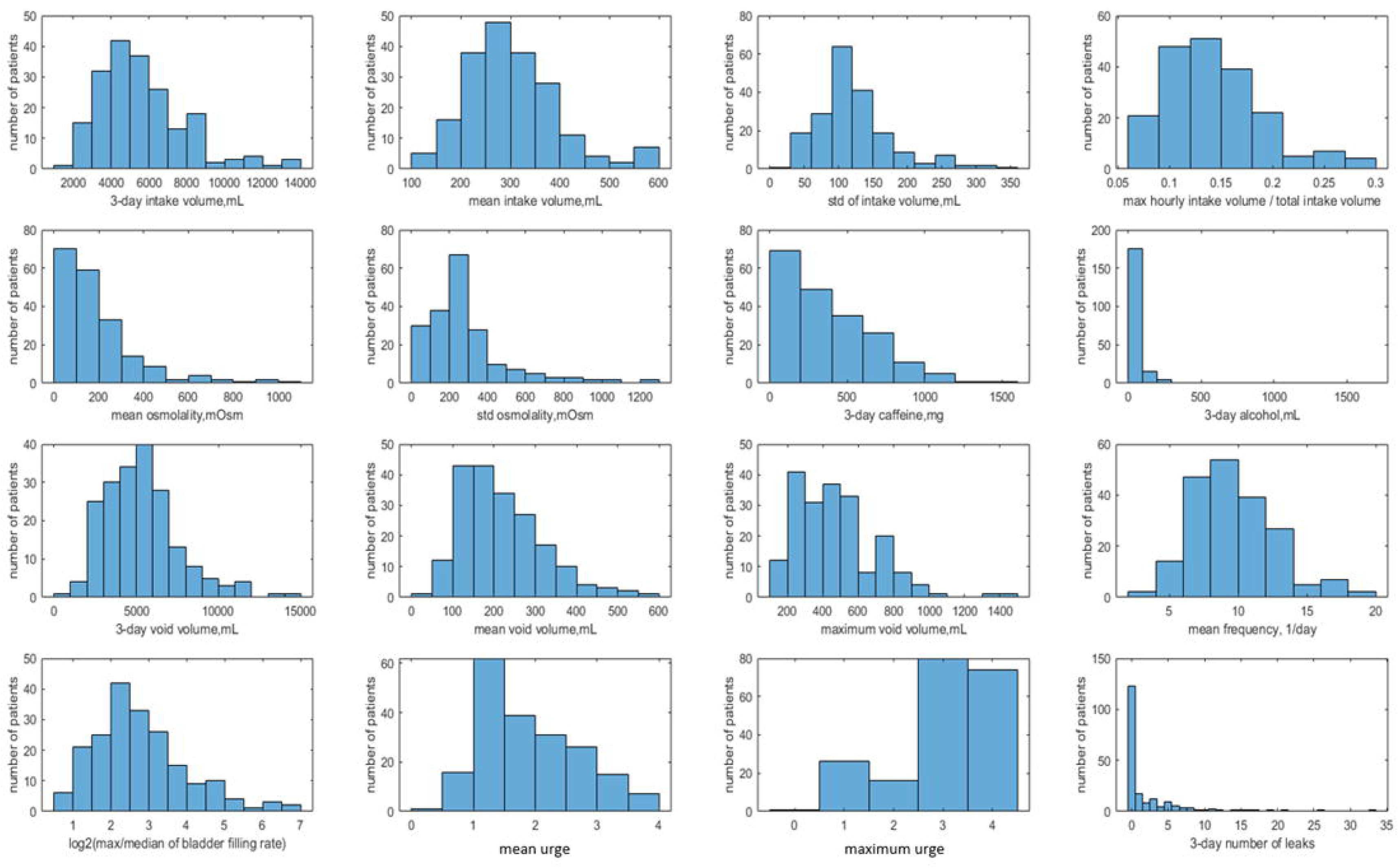
Distributions of the BD data and the derived dynamic variables across the 197 participants. Rows 1-2: intake properties. First row: 3-day intake volume (mL), mean intake volume (mL), standard deviation of intake volume (mL), ratio of maximum hourly intake volume, and total intake volume. Second row: mean osmolality of consumed drinks (mOsm), standard deviation of osmolality of the drinks, total consumed caffeine (mg), total consumed alcohol (mL). Rows 3-4: voids properties. Third row: 3-day void volume (mL), mean void volume (mL), maximum void volume, i.e., bladder capacity (mL), number of voids per day. Fourth row: log2 (max BFR(t)/median (BFR(t))) – measure of BFR versus time variability, mean urge, maximum urge, 3-day number of leaks. Note that our cohort is quite heterogeneous, with 3-day intake volume as low as 2L for some patients and as high as 14L for others, osmolality of the drinks varying from 0 to 1200 mOsm, both within the BD for a given participant and across participants. Bladder capacity varied from 200mL to 1400 mL, number of voids per day from 3 to 19, ratio of maximum to median BFR(t) from 1.6 to 128, mean urge from 0 to 4, and number of leaks during 3 days from zero to 33.

**Table 1.** Distribution of age, PVR, BMI, and time-averaged BD variables across 197 participants.

|  | cohort<br>minimum | cohort 25<br>percentile | cohort<br>median | cohort 75<br>percentile | cohort<br>maximum |
| --- | --- | --- | --- | --- | --- |
| Age, years | 20 | 49 | 61 | 70 | 84 |
| PVR, mL | 0 | 0 | 13 | 27 | 50 |
| BMI | 19.2 | 25.5 | 29.1 | 33.5 | 55.6 |
| 3-day intake volume, mL | 1567.4 | 4081.2 | 5205 | 6688.9 | 13722.3 |
| mean intake volume, mL | 109.2 | 242.3 | 291.2 | 351.7 | 597.4 |
| max hourly intake volume / total intake volume | 0.07 | 0.11 | 0.14 | 0.17 | 0.3 |
| mean osmolality, mOsm | 0 | 72.5 | 142.9 | 253.4 | 1049.2 |
| 3-day caffeine, mg | 0 | 100 | 300 | 542.5 | 1450 |
| 3-day alcohol, mL | 0 | 0 | 0 | 27.7 | 1632.5 |
| 3-day void volume, mL | 828.1 | 3672.7 | 5027.6 | 6458.2 | 14195.5 |
| mean void volume, mL | 48.7 | 142.5 | 198.1 | 276.5 | 563.6 |
| maximum void volume, mL | 103.5 | 295.7 | 414 | 591.5 | 1419.6 |
| mean/max void volume | 0.18 | 0.38 | 0.48 | 0.58 | 0.88 |
| mean frequency, 1/day | 3 | 7.6 | 9.4 | 11.2 | 18.9 |
| max/median of BFR | 1.6 | 3.8 | 6 | 10.1 | 128 |
| mean urge rating | 0 | 1.1 | 1.7 | 2.4 | 3.9 |
| max urge rating | 0 | 3 | 3 | 4 | 4 |
| number of leaks | 0 | 0 | 0 | 2.3 | 33 |

Intake volume variability during the day differed substantially across participants, with the ratio of maximum hourly intake volume to total 3-day intake volume ranging from 7% to 30%. Time-variability of void volumes during the day, measured as the ratio of mean to maximum void volume ranges from 0.18 to 0.88, with cohort median equal 0.48 and 75 percentile equal 0.58, supporting the observation [33] that the majority of people void when their bladder is at most 50% full. The ratio of maximum to median BFR for a given participant varies from a rather low 1.6 to extremely high value 128. Specifically, max/median BFR >4 in 70% of participants and >8 in 36% of participants, demonstrating that the high variability of BFR observed in Figure 2 and Supplemental Figures S1-S9 is a general feature of the cohort.

Table 2 provides values of significant correlation coefficients between the variables from Table1, sorted from stronger to weaker, and their partial correlation coefficients calculated with the rest of the variables kept constant.

**Table 2.**
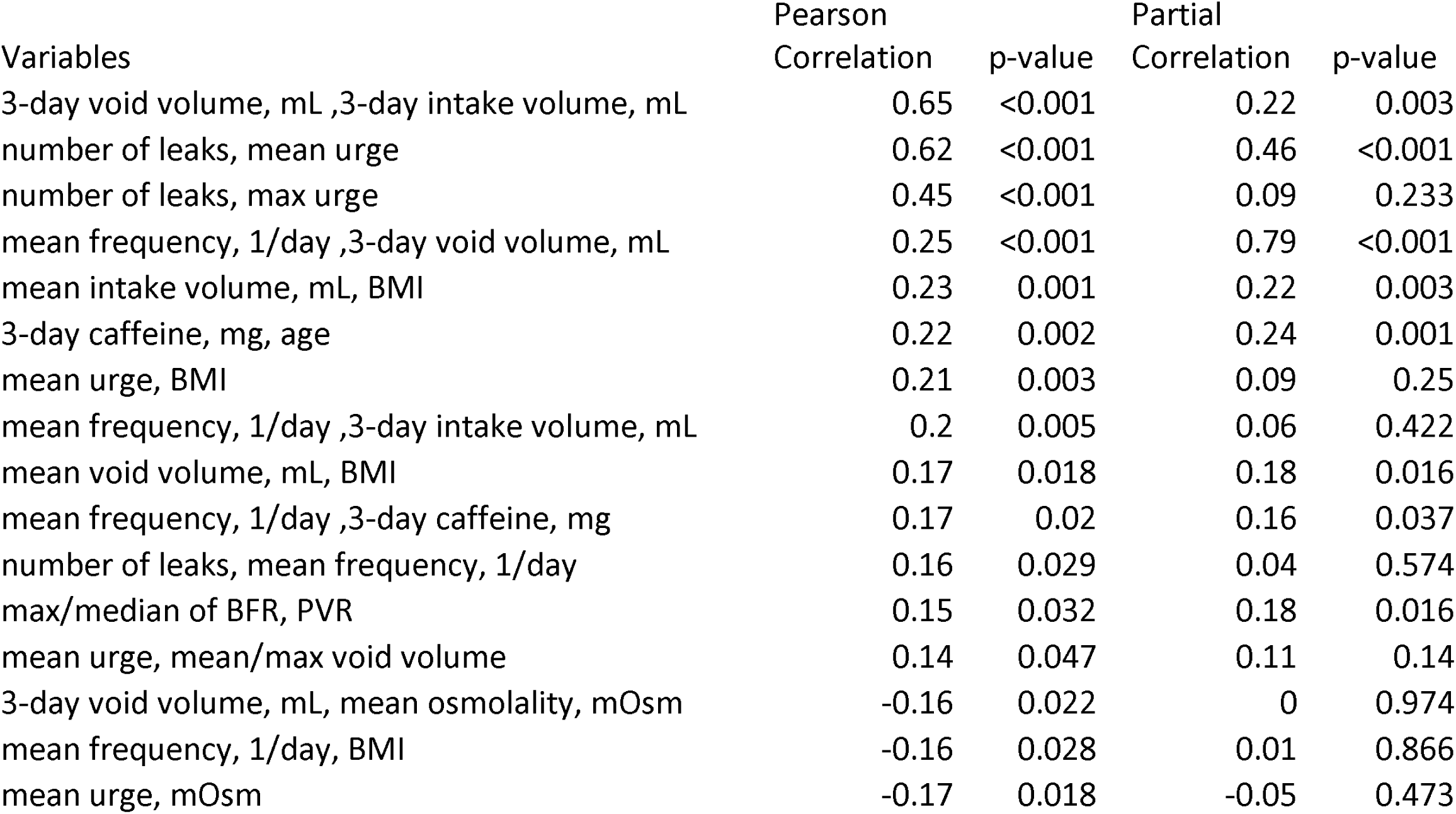
Inter-subject correlation coefficients between variables of Table 1.

| Variables | Pearson<br>Correlation | p-value | Partial<br>Correlation | p-value |
| --- | --- | --- | --- | --- |
| 3-day void volume, mL ,3-day intake volume, mL | 0.65 | <0.001 | 0.22 | 0.003 |
| number of leaks, mean urge | 0.62 | <0.001 | 0.46 | <0.001 |
| number of leaks, max urge | 0.45 | <0.001 | 0.09 | 0.233 |
| mean frequency, 1/day ,3-day void volume, mL | 0.25 | <0.001 | 0.79 | <0.001 |
| mean intake volume, mL, BMI | 0.23 | 0.001 | 0.22 | 0.003 |
| 3-day caffeine, mg, age | 0.22 | 0.002 | 0.24 | 0.001 |
| mean urge, BMI | 0.21 | 0.003 | 0.09 | 0.25 |
| mean frequency, 1/day ,3-day intake volume, mL | 0.2 | 0.005 | 0.06 | 0.422 |
| mean void volume, mL, BMI | 0.17 | 0.018 | 0.18 | 0.016 |
| mean frequency, 1/day ,3-day caffeine, mg | 0.17 | 0.02 | 0.16 | 0.037 |
| number of leaks, mean frequency, 1/day | 0.16 | 0.029 | 0.04 | 0.574 |
| max/median of BFR, PVR | 0.15 | 0.032 | 0.18 | 0.016 |
| mean urge, mean/max void volume | 0.14 | 0.047 | 0.11 | 0.14 |
| 3-day void volume, mL, mean osmolality, mOsm | -0.16 | 0.022 | 0 | 0.974 |
| mean frequency, 1/day, BMI | -0.16 | 0.028 | 0.01 | 0.866 |
| mean urge, mOsm | -0.17 | 0.018 | -0.05 | 0.473 |

Most of the inter-subject correlations are either trivial (e.g., correlations between 3-day intake and void volumes, number of leaks and mean urgency during 3 days, or mean daily frequency and 3-day void volume), or weak, or become weak when controlled for other variables, therefore not allowing for identification of definitive predictors of frequency, urgency, and incontinence episodes in our heterogeneous cohort and demonstrating the need for a dynamic approach to the analysis of individual BDs.

More interesting information is provided by the analysis of the distribution of VVs divided by the individual bladder capacity (estimated as the maximum VV during the 3-day BD). First, we performed analysis similar to [33] by creating histograms of the number of voids with the given ratio of VV to the individual bladder capacity. Then, we focused the analysis by looking only at the “very urgent” voids, i.e., voids with *U_j_* >=3. Finally, we investigated how many individuals had low-volume voids 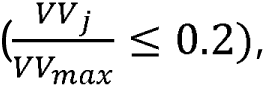 and low-volume “very urgent” voids. Results of this analysis are presented in Figure 4. Low-volume voids appeared to be quite prevalent: 134 of 197 (68%) individuals had low-volume voids, while 52 of 197 (26%) individuals had low-volume “very urgent” voids, i.e., voided with urge *U_j_* >=3, when only 20% or less of their bladder volume were filled with urine. While for the majority of individuals, low-volume “very urgent” voids were infrequent (20% or less of their voids), for some individuals, such voids happened as often as in 50% of cases. This analysis indicates that, at least for some of the individuals, the fraction of bladder volume filled with urine is not the only or even the main determinant of urinary urge. Next, we performed dynamic analysis of individual BDs in search of such determinants.

**Figure 4.**
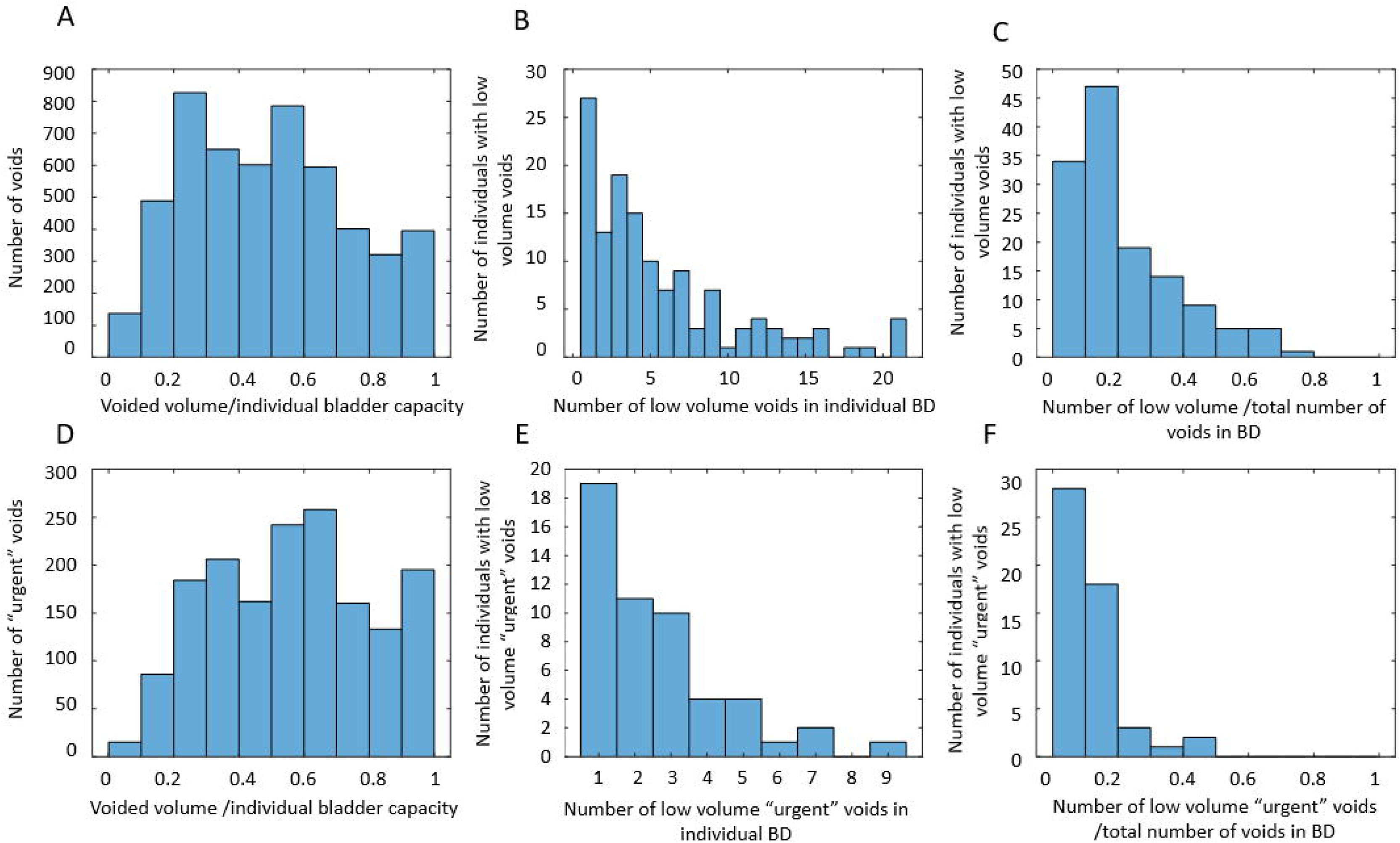
Distributions of the ratio of the voided volumes (VVs) to individual bladder capacity. Figure 4A: number of voids with the given ratio of VV to the individual bladder capacity in the cohort of 197 individuals. Figure 4B: number of individuals with low-volume voids (ratio<=20%) versus the number of such voids in the 3-day BD. Figure 4C: number of individuals with low-volume voids (ratio<=20%) versus the fraction of such voids in the 3-day BD. Figure 4D: number of “very urgent” voids (*U_j_* >=3) with the given ratio of VV to individual bladder capacity in the cohort of 197 individuals. Figure 4E: number of individuals with low-volume “very urgent” voids (ratio<=20%) versus the number of such voids in the 3-day BD.

Figure 4F: number of individuals with low-volume “very urgent” voids (ratio<=20%) versus the fraction of such voids in the 3-day BD.

### Dynamic analysis of BD data

In this section, we concentrate on dynamic analysis of individual BDs by visualizing individual intake and voiding patterns, calculating intra-subject correlations between dynamic BD variables, investigating similarities and time delays in the dynamics of these variables, and creating mathematical models of the individual BDs.

#### Intake and voiding profiles

Three main observations can be made from visualization of the individual intake and voiding profiles and dynamic variables derived from the BDs. Here, we demonstrate it for a typical participant A (Figure 2) and for nine other representative participants B to J (Supplemental Figures S1-S9). First, dynamic variables, i.e., bladder-filling rate (BFR), instantaneous time-dependent frequency of voiding (F), and urge growth rate (UrgR) can vary several-fold (e.g., 12-fold [Figure 2], 16-fold [Supplemental Figures S2, S3, S5, S7, S9]) during the day. Second, peaks of these functions occur at time points collocated with fluid intakes. The highest peaks of BFR(t), F(t), and UrgR(t) are closely following consumption of caffeine and/or alcohol, e.g., in participant A (Figure 2), and in participants B, C, D, and I (Supplemental Figures S2, S3, S4, S9). Other peaks are collocated with frequent and/or high-volume drinks. Third, periods of low BFR(t), F(t), and UrgR(t) are collocated with time intervals with no fluid consumption, usually at nighttime.

Next steps serve to formalize and quantify these observations by examining intra-subject correlations and cross-correlation functions of the above dynamic BD variables, and by developing personalized models of BDs.

#### Intra-subject correlations of dynamic BD variables

In this section, we will show how correlation analysis of time-dependent (dynamic) variables reveals a powerful correlation of BFR with UrgR and F. We will also show, in contrast, a lack of significant correlation of urge level at void with either BFR or VV. BD data for a typical participant A (Figure 2) can be presented in a different way by plotting UrgR versus BFR (Figure 5A) and versus VV (Figure 5B). Similarly, F can be plotted versus BFR (Figure 5C) and versus VV (Figure 5D). In this presentation, information regarding the sequence of events is lost; however, presence or absence of correlations between variables becomes more visible. For instance, for participant A, BFR is strongly and significantly correlated with UrgR (R=0.88, p= 3 · 10^-11^) and with F (R=0.85, p= 2 · 10^-10^), while correlation of these variables with VV is weak and unsignificant (R=-0.19, p=0.29; R=-0.30, p=0.08). Correlations of urge level at the time of void (URG) is weak and unsignificant with VV (R=0.32, p=0.06) and BFR (R=0.31, p=0.08) (Figures 5E-5F). As shown in Figure 6, these observations hold for the majority of 197 patients. While correlations of UrgR and F with BFR are almost always positive and are above 0.5 for the majority of participants, correlations of these variables with VV can be both positive and negative and are almost all weak (|R|<0.5). Figure 6C presents the correlations of URG with VV and BFR. Unlike UrgR and F, where correlation with BFR was a strong positive for almost all participants, there are participants with positive, zero, and negative correlations of URG with BFR. Similarly, there are participants with positive, zero, and negative correlations of URG with VV. Four extreme cases with the strongest positive and negative correlations are presented in Table 3. Note that the strongest negative correlation of URG with VV is observed for participant Y, consuming high osmolality drinks as well as caffeinated and alcohol-containing drinks. Negative correlations with BFR and VV are observed in participant Z, consuming the highest amount of caffeine. Likely, for these participants, urge level is determined not by the urine volume but by the composition of the drinks. The strongest positive correlation of URG with VV is observed for participant W, who consumed low osmolality drinks without caffeine and alcohol. The strongest positive correlation of URG with BFR was observed for participant X, with the highest time variability of BFR (max/median BFR=95.8). Observed differences in the correlations of URG with BFR and VV across participants suggest the differences in mechanism of urge, which could be potentially used for subtyping of patients with LUTS.

**Figure 5.**
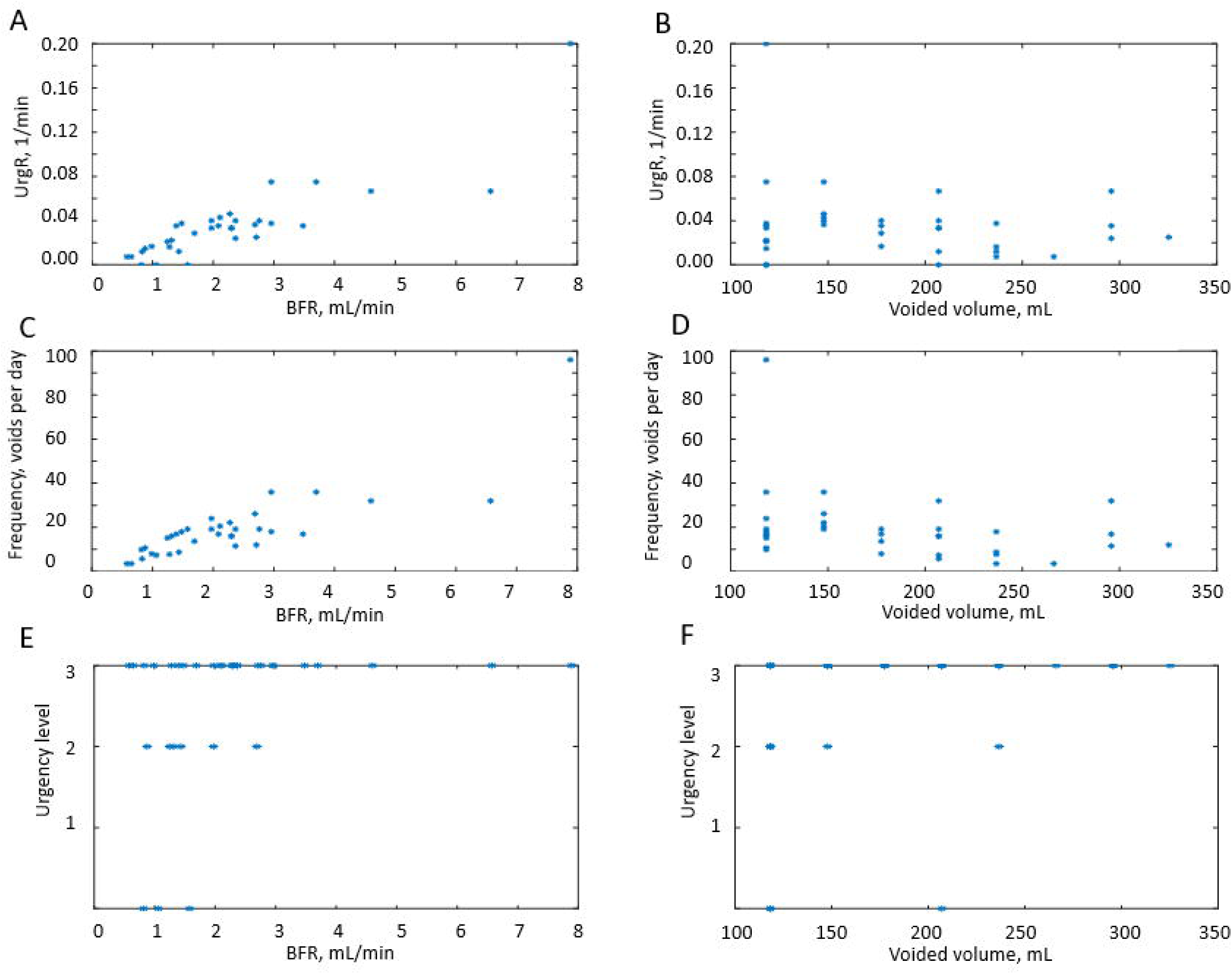
Example for a typical participant A of the correlations between the dynamic BD variables. Figure 5A: UrgR versus BFR (R=0.88, p= 3 · 10^-11^). Figure 5B: UrgR versus VV (R=-0.19, p=0.29). Figure 5C: F versus BFR (R=0.85, p= 2 · 10^-10^). Figure 5D: F versus VV (R=-0.30, p=0.08). Figure 5E: URG versus BFR (R=0.31, p=0.08). Figure 5F: URG versus VV (R=0.32, p=0.06).

**Figure 6.**
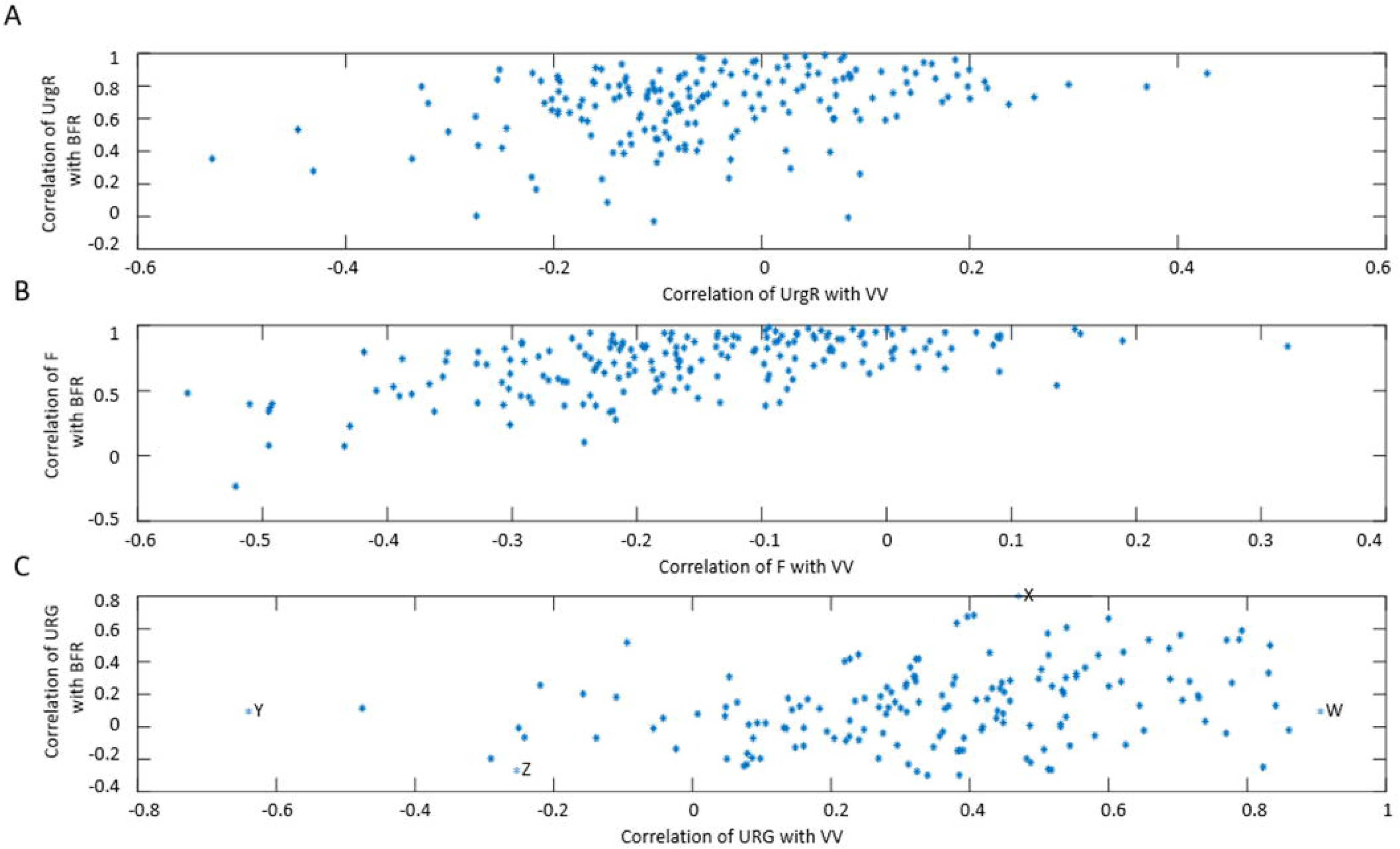
Scatter plots representing the values of correlation coefficients between dynamic variables for 197 participants. Figure 6A: correlation of UrgR with BFR (mean R=0.73) versus correlation of UrgR with VV (mean R=-0.07). Figure 6B: correlation of F with BFR (mean R=0.75) versus correlation of F with VV (mean R=-0.17). Figure 6C: correlation of URG with BFR (mean R=0.12) versus correlation of URG with VV (mean R=0.36). Four cases (W, X, Y, and Z with the highest and lowest values of R) are labeled and described in Table 3.

**Table 3.** Four participants with the strongest positive and negative correlations of URG with VV and BFR.

| ID | W | X | Y | Z |
| --- | --- | --- | --- | --- |
| Sex | Female | Female | Female | Male |
| Number of voids | 15 | 30 | 22 | 36 |
| Number of leaks | 2 | 0 | 1 | 6 |
| Total VV, mL | 7097.8 | 3645.1 | 5352.9 | 6713.3 |
| Mean URG | 2.4 | 1.21 | 2.86 | 1.53 |
| Max URG | 4 | 3 | 4 | 4 |
| Number of intakes | 15 | 9 | 19 | 21 |
| Total intake volume, mL | 4864.9 | 3312.3 | 4199.5 | 5648.6 |
| Total caffeine, mg | 0 | 150 | 200 | 700 |
| Total alcohol, mL | 0 | 0 | 45 | 0 |
| Mean osmolality, mOsm | 133.3 | 240 | 404.2 | 130 |
| Max/median BFR | 3.12 | 95.83 | 6.36 | 5.69 |
| Correlation of URG with VV | 0.91 | 0.48 | -0.63 | -0.24 |
| Correlation of URG with BFR | 0.05 | 0.76 | 0.09 | -0.32 |

Table 4 presents the values of intra-subject correlation coefficients between dynamic BD variables calculated for each participant and then averaged across 197 participants. As seen, BFR, F, and UrgR are strongly and significantly correlated (correlations are preserved when controlled for other variables; see values of partial correlation coefficients); correlations of these variables with VV are weak, indicating that BFR rather than VV is the main determinant of urinary frequency and UrgR. This conclusion corroborates the observation of numerous low-volume “urgent” voids illustrated by Figure 4. There are two aspects of the correlation of UrgR with BFR. The first is obvious: A higher BFR results in faster bladder-filling to a level that produces a sensation of urge and a greater frequency of urination. The second aspect is revealed when we compare the full correlation coefficients with the partial correlation coefficients calculated when certain variables are fixed. For instance, partial correlation of BFR with UrgR (R=0.758) with fixed VV is even higher than full correlation of BFR with UrgR (R=0.733), while partial correlation of BFR with UrgR with fixed urinary frequency is relatively weak (R=0.3, p-value<0.001), indicating that the role of high BFR cannot be reduced to the time necessary to fill the bladder to a certain level. We further investigate the role of BFR and urine volume in the regression models of UrgR.

**Table 4.** Intra-subject correlations between dynamic BD variables averaged across 197 participants.

| Variables | Pearson<br>Correlation | p-value | Partial<br>Corr,<br>VV<br>fixed | p-value | Partial<br>Corr,<br>BFR<br>fixed | p-value | Partial<br>Corr,<br>IR<br>fixed | p-value | Partial<br>Corr,<br>F<br>fixed | p-value |
| --- | --- | --- | --- | --- | --- | --- | --- | --- | --- | --- |
| F, UrgR | 0.807 | <0.001 | 0.807 | <0.001 | 0.597 | <0.001 | 0.803 | <0.001 |  |  |
| BFR, F | 0.752 | <0.001 | 0.797 | <0.001 |  |  | 0.753 | <0.001 |  |  |
| BFR, UrgR | 0.733 | <0.001 | 0.758 | <0.001 |  |  | 0.728 | <0.001 | 0.3 | <0.001 |
| BFR, VV | 0.151 | <0.001 |  |  |  |  | 0.16 | <0.001 | 0.395 | <0.001 |
| VV, UrgR | -0.07 | <0.001 |  |  | -0.231 | <0.001 | -0.061 | <0.001 | 0.103 | <0.001 |
| VV, F | -0.167 | <0.001 |  |  | -0.395 | <0.001 | -0.159 | <0.001 |  |  |

#### Investigating similarities between BD variables using cross-correlation functions

Intra-subject correlation analysis in the previous section helped to reveal important relationships between variables in individual BDs; however, in such analysis, information about the sequence of intake and voiding events is lost. To preserve this information and analyze similarities in profiles of the variables, including co-occurrence of events, we used cross­correlation functions, calculated as explained in the Methods section.

Cross-correlation functions of dynamic BD variables for a typical participant A are presented in Figure 7 and for nine other typical participants (B-J) in Supplemental Figure S10. The maxima of the cross-correlation functions of BFR(t), F(t), and UrgR(t) for participant A occurred at zero lag and have high values of over 0.96 (highest possible value of cross­correlation function is 1), demonstrating the high level of similarity of the profiles and absence of time delay between these functions. This result corroborates the observation from Figure 2 that peaks of BFR(t), F(t), and UrgR(t) are co-occurring (e.g., labeled peaks at X=30, Figure 2). It also corroborates the result of the previous section that urinary frequency and UrgR are highly correlated with the BFR. The presence of two smaller maxima of these correlation functions at ±24 hours indicate the similarity of voiding patterns in days 1,2,3 of the BD, i.e., stability of the daily voiding pattern for this participant. Note that peaks of cross-correlation function at ±24 hours occur for some (B, D, G, H, I, J) but not for all participants in Supplemental Figure S10, indicating that similarity of voiding patterns of days 1,2,3 of the BD is not a universal, but a patient-specific property.

**Figure 7.**
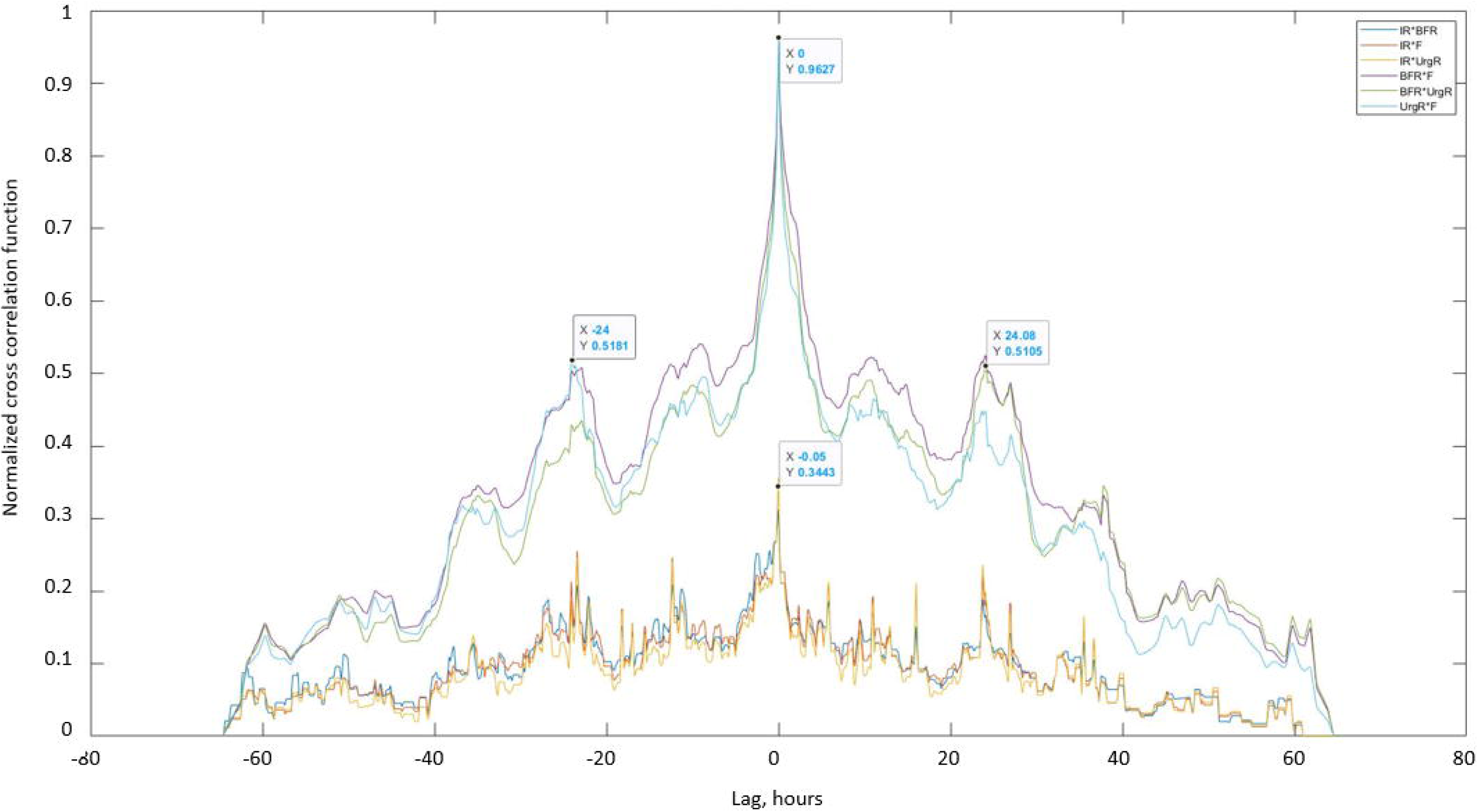
Cross-correlation functions between dynamic BD variables of typical participant A. Cross-correlation functions of BFR(t), UrgR(t), and F(t) have high maxima (Xcorrmax=0.96) at zero lag (X=0), indicating high similarity and absence of time delay between these functions. Two more maxima of these cross-correlation functions (X=±24 hour) indicate similarity of voiding patterns in days 1,2,3 of BD. Lower values of maxima of cross-correlation functions of frequency and UrgR with intake rate IR(t) (X=-0.05, Xcorrmax=0.34) indicate that peaks in intake volumes alone cannot predict peaks in voiding variables.

Cross-correlation functions of urinary frequency and UrgR with IR(t) demonstrate much lower values with maxima of about 0.34, indicating that peaks in intake volume alone cannot predict peaks in voiding variables. This corroborates with observations in Figure 2, where the highest peak (at X=30) co-occurred not with the highest intake volume but with the intake containing caffeine. Similar cross-correlation functions can be observed in Supplemental Figure S10 for the nine representative participants (B-J). For all of these participants, the maxima of cross-correlations of F and UrgR with BFR are in the range of [0.88, 0.98], which is three- to four-fold higher than their correlations with the IR(t). This observation makes sense, since IR(t) captures information only of the volumes and durations of the drinks, while urine production rate and therefore BFR may be affected by the composition of the drinks, e.g., osmolality, caffeine, and alcohol content. Also, importantly, BFR is affected not only by the volume of fluid consumed at the most recent intake but at the previous intakes as well, as seen for the peak of BFR (at X=68.3) following several peaks of intake (around X=66.8) in Figure 2, or similarly, for the peaks of BFR and UrgR (at X=14.23) following a series of intakes (at X=13.5) in Supplemental Figure S2. Therefore, analysis with cross-correlation functions confirmed conclusions of the analysis of intra-subject correlations on high correlations and co-occurrence of peaks of urinary frequency and UrgR with the BFR, but also demonstrated that more sophisticated methods of analysis are necessary to predict these peaks from the intake profiles.

#### Predicting BFR profile with the grey-box model of urine formation rate

As described in the Methods section and in Supplemental Material, we developed renal physiology-based grey-box models of urine formation rate for 197 individuals by fitting parameters of the model equations to minimize the difference of the urine formation rate predicted by the models and BFR calculated from the BDs of these individuals.

Figure 8 presents the comparison of BFR profiles derived from the BD with results of the grey-box models of urine formation rate for participants A (Figure 8A), B (Figure 8B), D (Figure 8C), and J (Figure 8D). Two versions of the model are presented, i.e., the “full model” and the model ignoring caffeine and alcohol contents of drinks (“no caf no alc”). As seen, the “full model” is capable of better describing the peaks of BFR especially collocated with intakes containing caffeine. The “no caf no alc” model predicts peaks collocated with intakes; however, these peaks are lower and broader than observed and predicted by the full model, as seen for participants A and C (Figures 7A, 8C). Figures 8B and 8D demonstrate that models ignoring caffeine and alcohol content of the drinks not only have problems in accurately predicting high peaks collocated with caffeine and alcohol consumption, but also sometimes create high false peaks collocated with drinks without caffeine and alcohol (e.g., around 7 pm on the first and third days in Figure 8B and on multiple locations in Figure 8D). This is the consequence of “no caf no alc” models not differentiating between different types of drinks and therefore attributing higher effects to the volume of the drinks. Although not ideal, “full models” are capable of better prediction of BFR profiles, with fewer peaks missed and fewer false peaks created. When comparing predicted and derived (from the BD) BFR profiles, it is important to remember that the later profiles are approximations as well, based on the assumption of BFR being constant during the interval between the voids, which is not necessarily correct and might be affected by multiple factors not recorded in the BD, e.g., heart rate, level of physical activity, consumption of salted food and/or diuretics.

**Figure 8.**
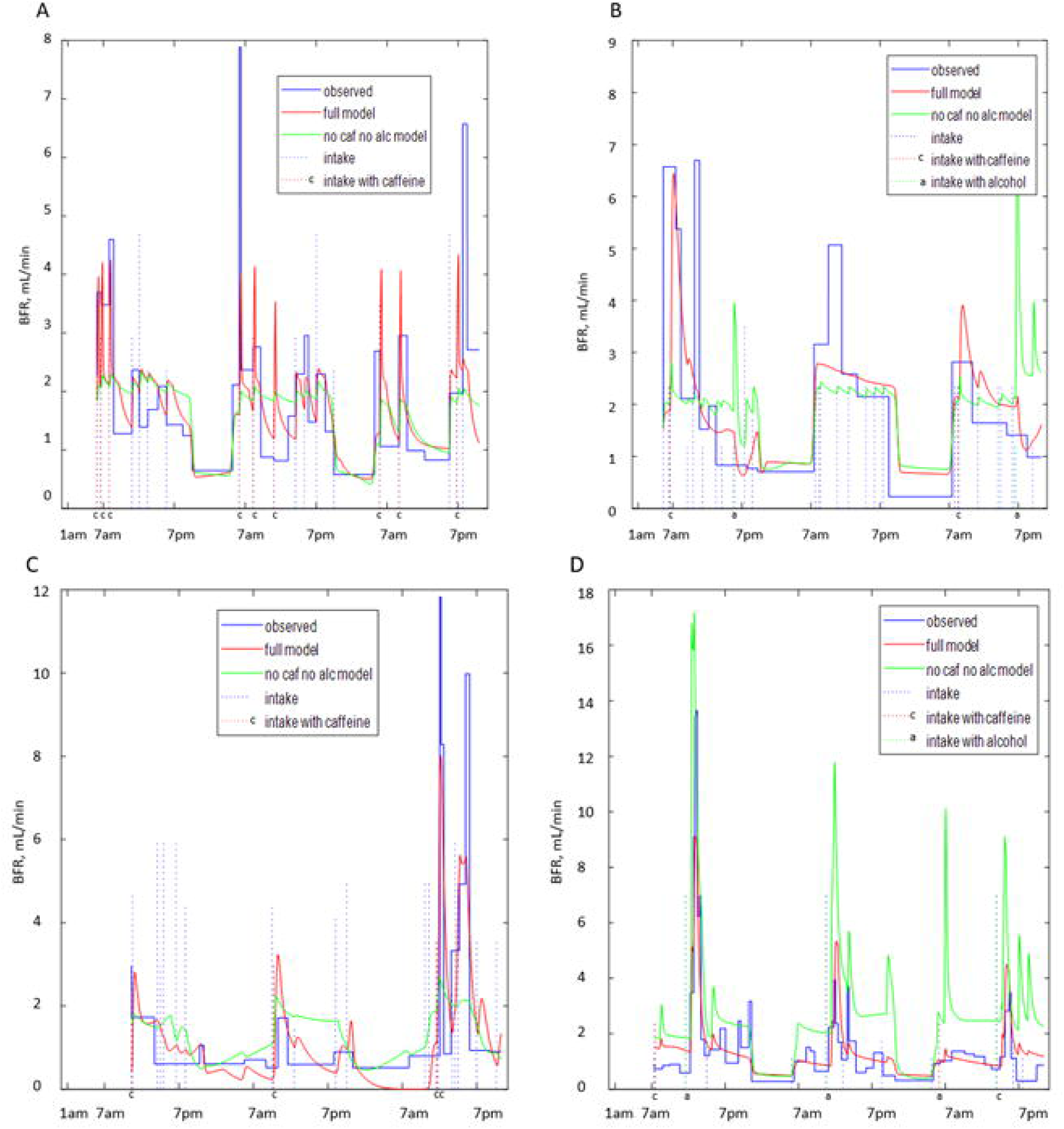
Comparison of BFR profiles derived from the BDs with the profiles predicted by the grey-box models of urine formation rate. Figure 8A: participant A. Figure 8B: participant B. Figure 8C: participant D. Figure 8D: participant J. Both “full models” (in red) and models ignoring caffeine and alcohol content of the drinks (in green) are presented.

#### Evaluation and selection of the BFR models

We evaluated the developed grey-box models by comparing BFR profiles predicted by the models and derived from the BD. We performed not only visual comparison (Figure 8) but also quantified similarities by using cross-correlation function (eq 1) and functions Peak Fit and Peak Lag (eqs. 3-4). As noted in the Methods section, an important quality of any model is its ability to describe/predict observations in the time range outside of the time range used for parameter fitting. To estimate this ability, we developed models using 3 days of BD data (3-day model), as well as models using only the first 2 days of BD data (2-day model). Then we used the parameters of the 2-day model and 3-day intake data to predict BFR(t) during all 3 days.

The two rows in Figure 9 serve to compare the similarity of the observed BFR(t) and the 3-days (Figure 9A-9C) versus 2-day models (Figure 9D-9F). As seen, the histograms are rather similar, indicating that 2-day model predictions are only slightly less accurate than 3-day models. For the majority of the 3-day and 2-day models, maximum of the cross-correlation function (max X_corr_) is above 0.8 (Figures 9A, 9D). For the majority of the models, Peak Fit>0.9 (Figures 9B, 9E) and Peak Lag <6.5 minutes (Figures 9C, 9F). We used the Peak Fit metric to select satisfactory models for further analysis. Specifically, we selected BDs for whom both 3-day and 2-day models have similar accuracy, i.e., Peak Fit >0.9 for 3-day and 2-day models. We found 145 such BDs. Table 5 provides the comparison of the 3-day and 2-day models for these selected BDs by showing mean values of maximum and lag of cross-correlation function, as well as mean values of Peak Fit and Peak Lag metrics. As seen, both 3-day and 2-day models provide satisfactory prediction of peak heights and positions, and the 2-day models are only slightly less accurate than the 3-day models.

**Figure 9.**
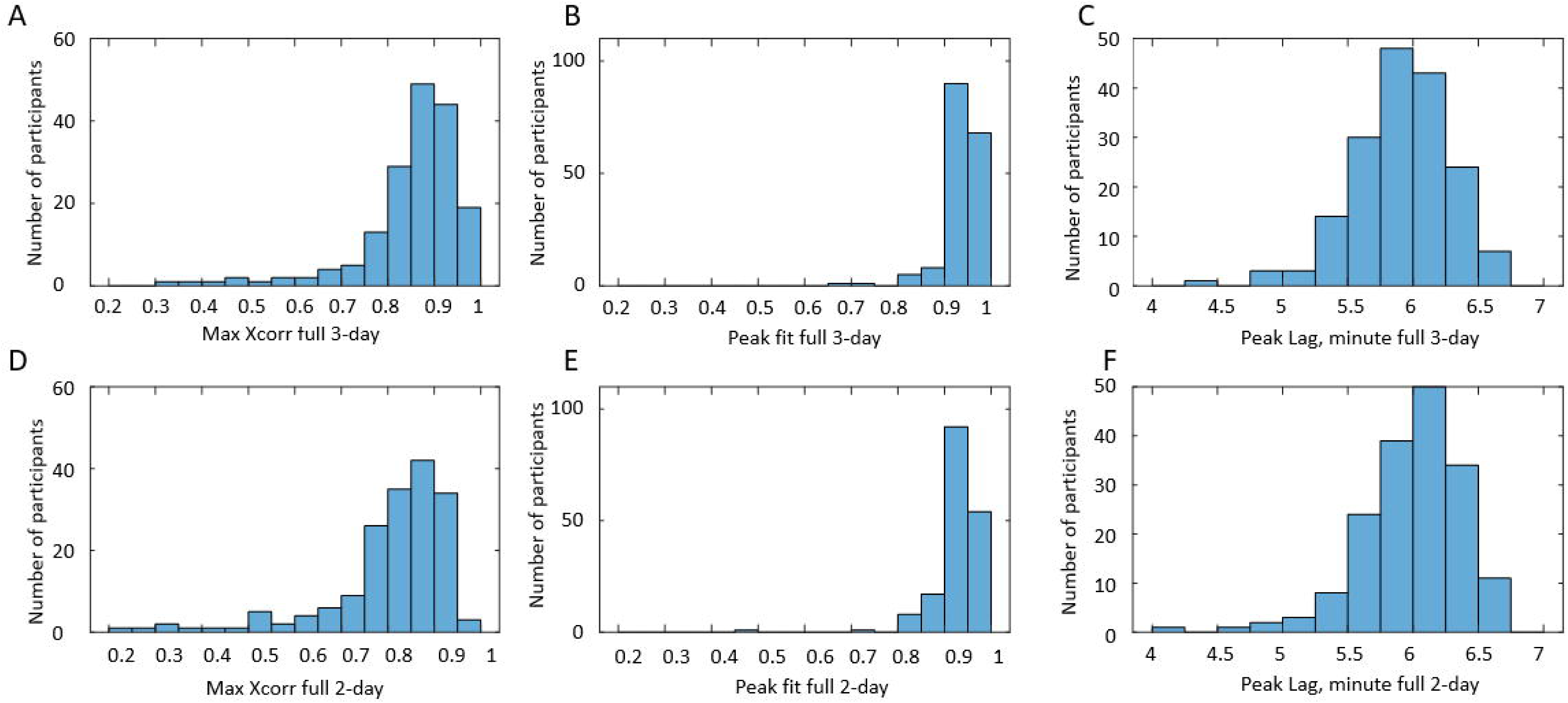
Distributions of metrics of similarity between modeled and observed (derived from BD) profiles of BFR for 197 participants. Figures 9A-9C: 3-day models. Figures 9D-9F: 2-day models. Figures 9A, 9D: distribution of maximum values of cross-correlation function. Figures 9B, 9E: distribution of Peak Fit values. Figures 9C, 9F: distribution of Peak Lag values.

**Table 5.** Metrics for comparison of 3-day and 2-day BFR full models with BFR derived from selected 145 BDs. Mean (std) values.

| | Peak Fit | Peak Lag, in minutes | $\max X_{\text{corr}}$ | $\tau_{\max}$, in minutes |
| --- | --- | --- | --- | --- |
| BFR full 3-day | 0.947 (0.017) | 5.91 (0.38) | 0.877 (0.07) | -2.32 (16) |
| BFR full 2-day | 0.943 (0.018) | 5.99 (0.39) | 0.844 (0.08) | 2.88 (60) |

#### Predictions of linear regression models of urinary UrgR

As described in the Methods section, we developed multivariable linear regression models of UrgR for each individual with the following three inputs: BFR, volume of urine in the bladder, and awake (yes/no or 1/0). Figure 10 provides the comparison of profiles of UrgR(t) derived from the BDs with those predicted by the developed multivariable linear regression models for four typical participants (A, B, E, and F).

**Figure 10.**
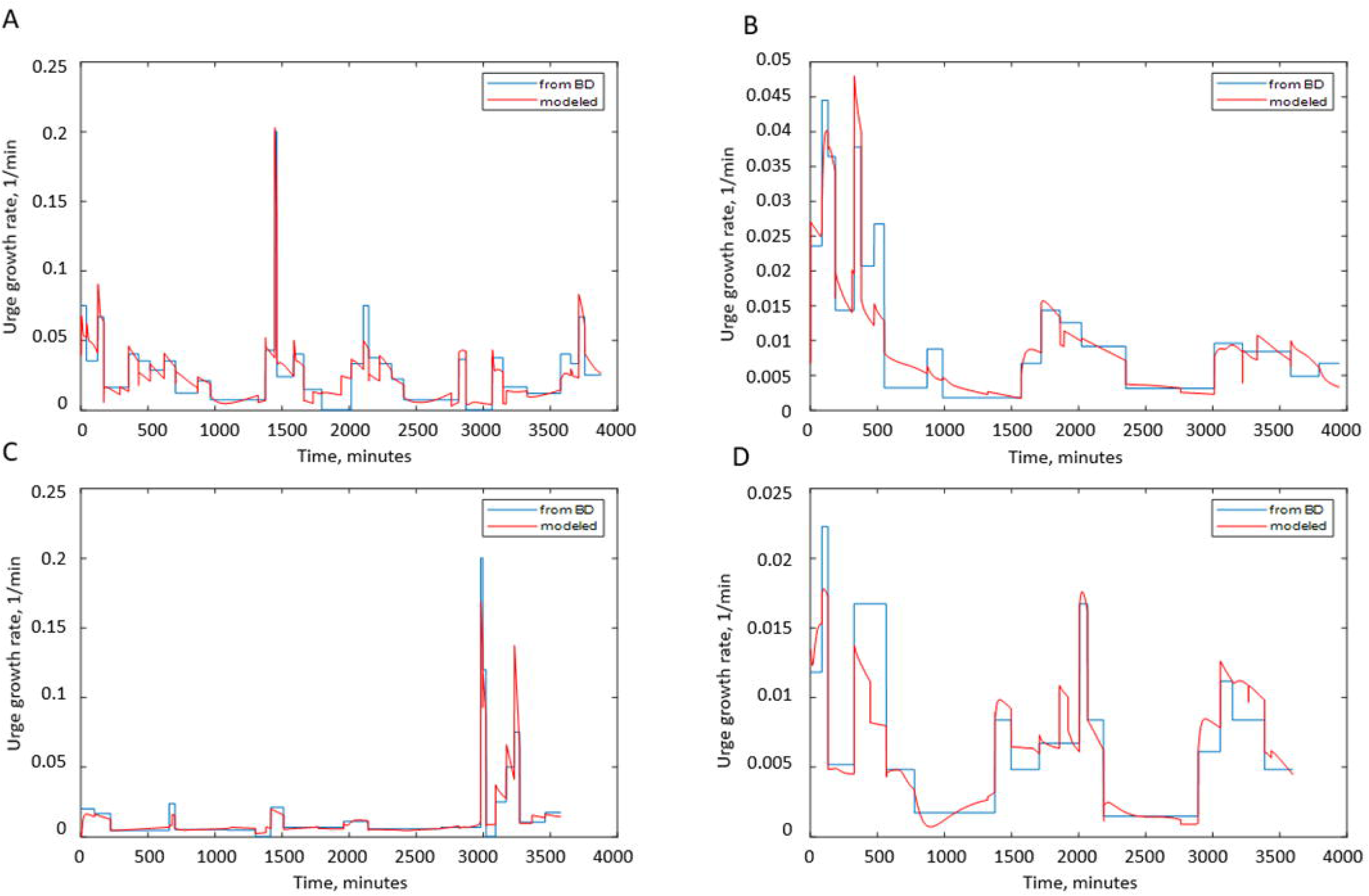
Comparison of profiles of UrgR predicted by the models and derived from the BDs for four typical participants. Figure 10A: participant A (Adj.R^2^ =0.88). Figure 10B: participant B (Adj.R^2^ =0.87). Figure 10C: participant E (Adj.R^2^ =0.86). Figure 10 D: participant F (Adj.R^2^ =0.77). Note that intake and voiding profiles for these participants are presented in Figure 2 and Supplemental Figures S1, S5, and S6.

The quality of the multivariable linear regression models (the proportion of variance explained by the model) is estimated by Adj.R^2^. The distribution of Adj.R^2^ for 145 participants is presented in Figure 11A (0.1< Adj.R^2^<0.95). There is no universal rule to judge the quality of the model by the value of Adj.R^2^. It is recognized that higher values of Adj.R^2^ are expected in physics and chemistry and lower in biology, medicine, and sociology. Typically, models of the processes involving human behavior are considered “good enough” if Adj.R^2^>0.3 [34]. We applied this threshold and selected for further analysis the 124 participants for whom Adj.R^2^>0.3 in the multivariable linear regression model of UrgR. These 124 participants are a subset of 145 participants for whom the satisfactory grey-box models (Peak Fit>0.9) of urine production rate were created. We used the predicted profiles of UrgR(t) to calculate predicted urge scores at the times of void by using eq. 7 and compared the urge scores predicted by the models and reported in BD, as described in Methods, to determine percentages of true positive and false positive predictions of urgency episodes during individual 3-day BDs. The distributions of true positives and false positives are presented in Figures 11B and 11C. For 87 (70%) of individuals, models correctly predicted more than 90% of urgency episodes. Similarly, models predicted less than 10% false positive episodes in 82 (66%) individuals. The mean percentage of true positive predictions across 124 participants is 84.6%, while the mean false positive is 10.9%.

**Figure 11.**
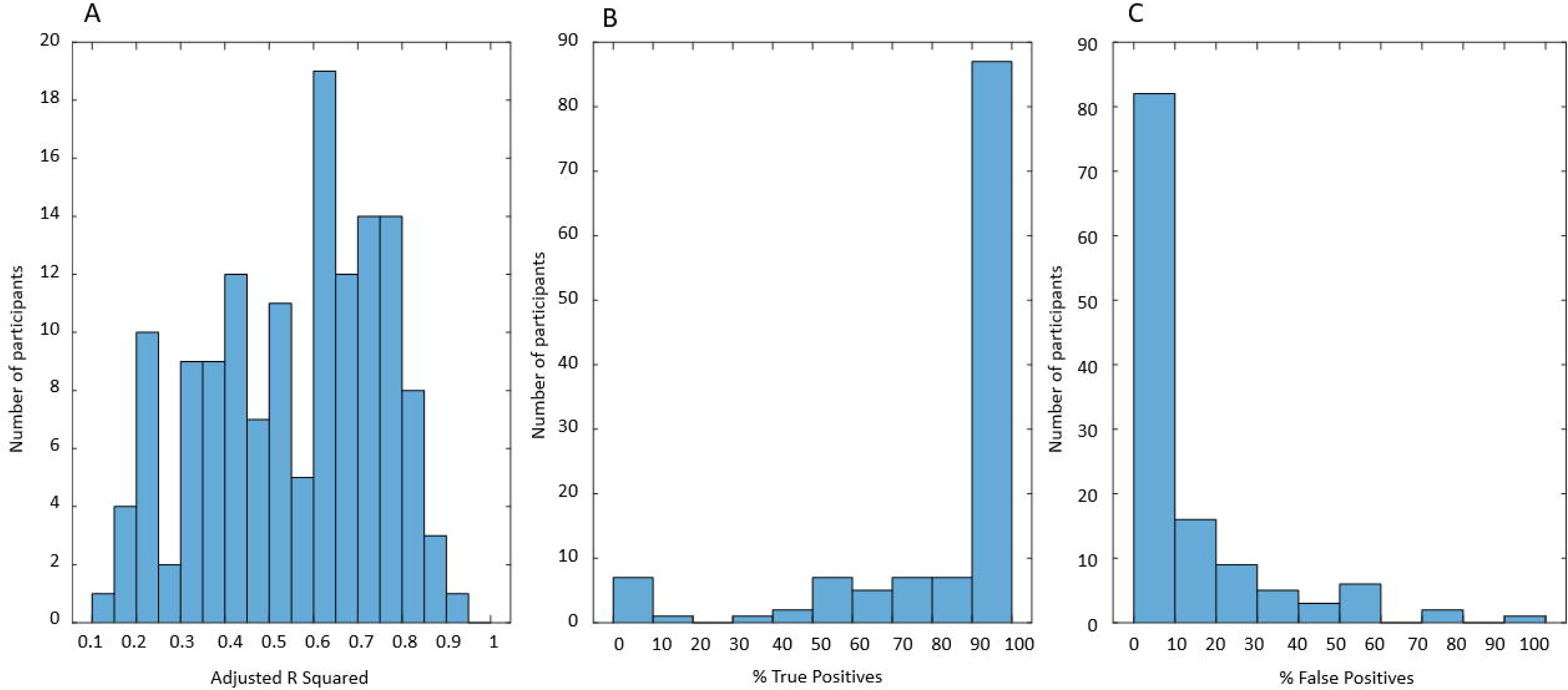
Quality measures of the multivariable linear regression model of UrgR and urge scores at voids. Figure 11A: Distribution of adjusted R squared values for 145 individuals’ UrgR model (mean Adj.R^2^ =0.67). Figure 11B: Distribution of percentage of true positive predictions of urgency episodes in subset of 124 participants with Adj.R^2^ >0.3. Figure 11C: Distribution of percentage of false positive predictions of urgency episodes in the same subset of 124 participants. There is a rather weak positive correlation between Adj.R^2^ of the model and percentage of true positive predictions of urgency episodes (R=0.3, p=0.006), and very weak negative correlation between Adj.R^2^ and false positive predictions (R=-0.17, p=0.054).

One goal of creating multivariable linear regression models was to investigate how BFR and urine volume affect the UrgR in various individuals. We compared the group of participants (n=106) experiencing at least one episode of urgency (URG≥2) with those without urgency (n=18). Table 6 provides the mean and standard deviation values of the linear model coefficients for these two groups.

**Table 6.** Comparison of coefficients of the linear model of UrgR for patients with and without urinary urgency.

|  | mean with urgency | std with urgency | mean without urgency | std without urgency | p-values |
| --- | --- | --- | --- | --- | --- |
| Intercept | 0.0124 | 0.0064 | 0.0061 | 0.0013 | 5.8E-05 |
| $\beta_1$ ($\text{BFR}^0$) | 0.0098 | 0.0054 | 0.0042 | 0.0021 | 3.6E-05 |
| $\beta_2$ ($V^0$) | -0.0022 | 0.0020 | -0.0010 | 0.0004 | 2.0E-02 |
| $\beta_3$ (awake) | -0.0014 | 0.0037 | -0.0005 | 0.0012 | 3.9E-01 |
| $\beta_{12}$ ($\text{BFR}^0 * V^0$) | -0.0003 | 0.0016 | -0.0003 | 0.0005 | 9.5E-01 |
| $\beta_{13}$ ($\text{BFR}^0 * \text{awake}$) | -0.0008 | 0.0065 | -0.0004 | 0.0022 | 8.1E-01 |
| $\beta_{23}$ ($V^0 * \text{awake}$) | 0.0008 | 0.0023 | 0.0001 | 0.0007 | 2.9E-01 |
| $\text{Adj.R}^2$ | 0.591 | 0.162 | 0.660 | 0.143 | 9.1E-02 |

Intercept is significantly higher in the group with urgency. Similarly, β_1_ is positive in both groups but is significantly higher in the group with urgency, indicating that BFR is a strong driver of urgency. Mean of ß_2_ is negative in both groups but has two-fold larger absolute value in the group with urgency. Mean negative value of the coefficient for *ß_2_* is in agreement with the negative mean intra-subject correlation of UrgR with urine volume (Table 4). The negative value of *ß_2_* might seem counterintuitive; one would expect urge to be higher with the higher urine volume when everything else is kept equal. However, it is important to remember that our model

(eq 6) predicts urge growth rate not urge level, meaning that with negative β_2_, urge is growing with the increase of urine volume but slower than linearly, suggesting possible saturation of the signal produced by the stretched bladder wall. Mean of β_3_ is negative in both groups but has nearly three-fold larger absolute value in the group with urgency, indicating that, when everything else is kept equal (i.e., BFR and urine volume), on average, UrgR is higher when participants are asleep than when they are awake in both groups, but to a larger extent in participants with urgency.

Distributions of the intercept and model coefficients for participants with and without urgency in Figure 12 corroborates this observation. It also demonstrates that distributions of intercept and all coefficients are broader for participants with urgency relative to those without urgency. Rephrasing Leo Tolstoy, one can say that all people without urgency are “non-urgent alike”, while each person with urgency has urgency in their own way. Note that, while intercept and β_1_ are positive for all 124 participants (both with and without urgency), the values of intercept and β_1_ are higher for participants with urgency; *β_2_* is negative for all participants without urgency and for the majority (107, or 92%) of participants with urgency. Coefficient β_3_ is negative for the majority of participants (85 with urgency and 12 without urgency), but is positive for some participants (31 with urgency and 6 without urgency), indicating that being awake could increase or decrease urgency growth rate in participants from both groups.

**Figure 12.**
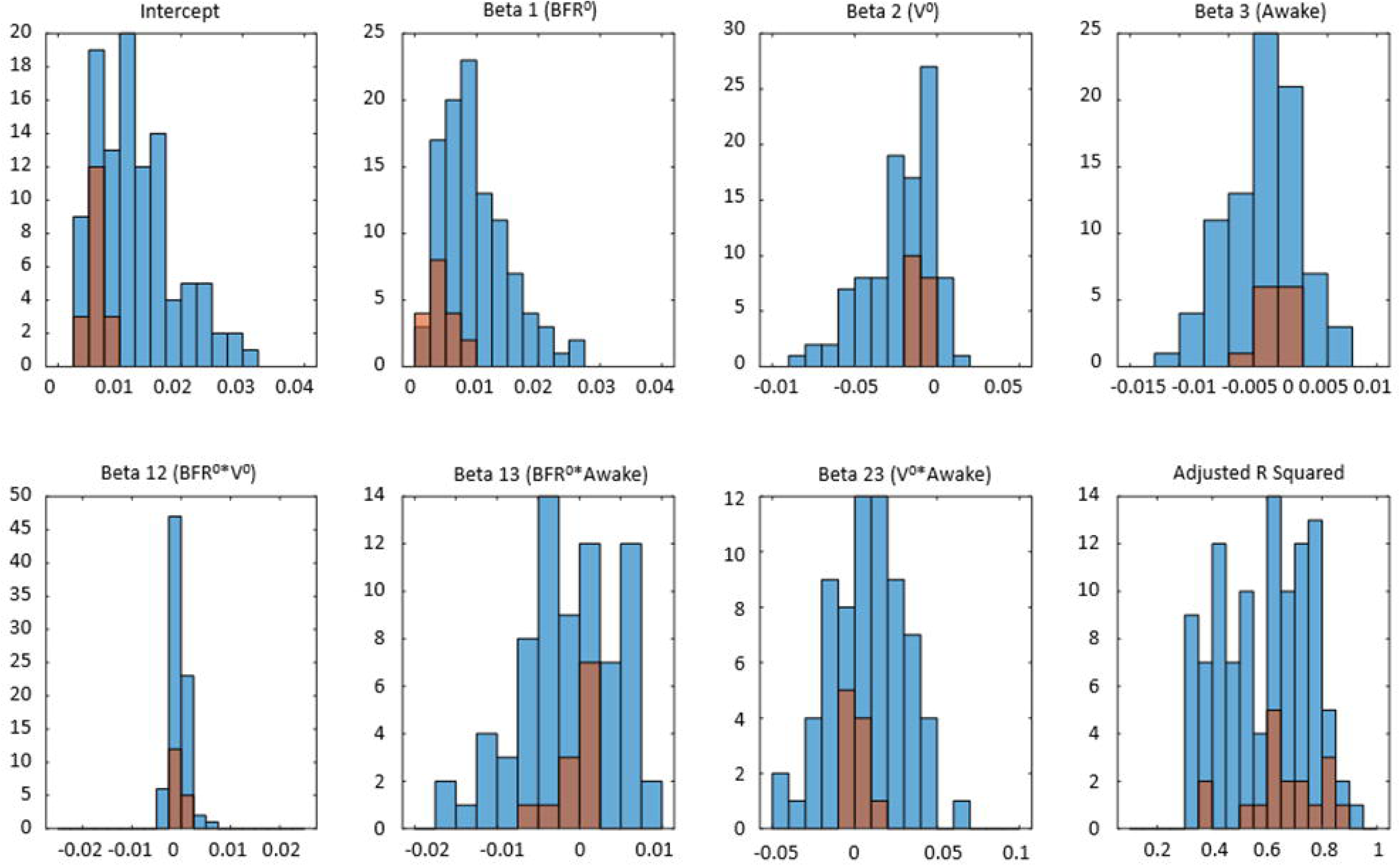
Distributions of the intercept and 6 coefficients of multi linear regression models of UrgR across 124 participants. Blue: participants with urgency (n=106). Red: participants without urgency (n=18). Bottom-right panel compares the distributions of Adj.R^2^ values for participants with and without urgency (no significant difference).

There are several metrics to estimate the severity of urgency in a patient; one possibility is to count the number of urgent voids. Table 7 provides correlations between the number of urgent voids with coefficients of the linear regression models across 124 participants.

**Table 7.** Inter-subject Pearson correlation coefficients between the number of urgent voids and coefficients of linear regression models of UrgR.

| Coefficient of variable | Correlation with number of urgent voids | p-values |
| --- | --- | --- |
| Intercept | 0.83 | <0.001 |
| $\beta_1$ | 0.54 | <0.001 |
| $\beta_2$ | -0.41 | <0.001 |
| $\beta_3$ | -0.32 | 0.002 |
| $\beta_{12}$ | -0.10 | 0.34 |
| $\beta_{13}$ | 0.09 | 0.39 |
| $\beta_{23}$ | 0.16 | 0.17 |

Number of urgent voids is strongly, significantly, and positively correlated with the intercept and coefficient ß1. Correlation with the intercept is obvious and trivial (it just means that participants with higher UrgR have higher number of urgent voids). Positive correlation with the coefficient β_1_ is more interesting and indicates that, not only the BFR is driving urge, but the participants with more frequent urgent voids are more sensitive to the increased BFR. Number of urgent voids is also significantly negatively correlated with β_2_, suggesting that participants with a high number of urgent voids are less sensitive to the increased volume of urine in the bladder relative to the increased BFR.

It is of interest to examine if the slopes (sensitivities to different factors) are correlated, e.g., are people more sensitive to the increase of BFR also more sensitive to the increase of urine volume or to being awake. Scatter plots of intercept and coefficients β_1_ and β_2_ in Figure 13 demonstrate that intercept and β_1_ are positively correlated (R=0.47, p<0.001), meaning that the larger the mean UrgR for the participant, the higher their sensitivity to the increase in BFR. On the contrary, correlation between β_1_ and β_2_ is negative (R=-0.33, p<0.001), indicating that participants more sensitive to increased BFR are less sensitive to increased urine volume. Note that correlation between β_1_ and β_2_ is different from the interaction term β_12_, which deals with interactions of BFR and urine volume within a model for each individual. Negative ß_12_ indicates slower than linear growth of UrgR with BFR. Correlation between β_1_ and β_2_ are across the individuals. Negative correlation indicates that participants more sensitive to increased BFR are less sensitive to increased urine volume.

**Figure 13.**
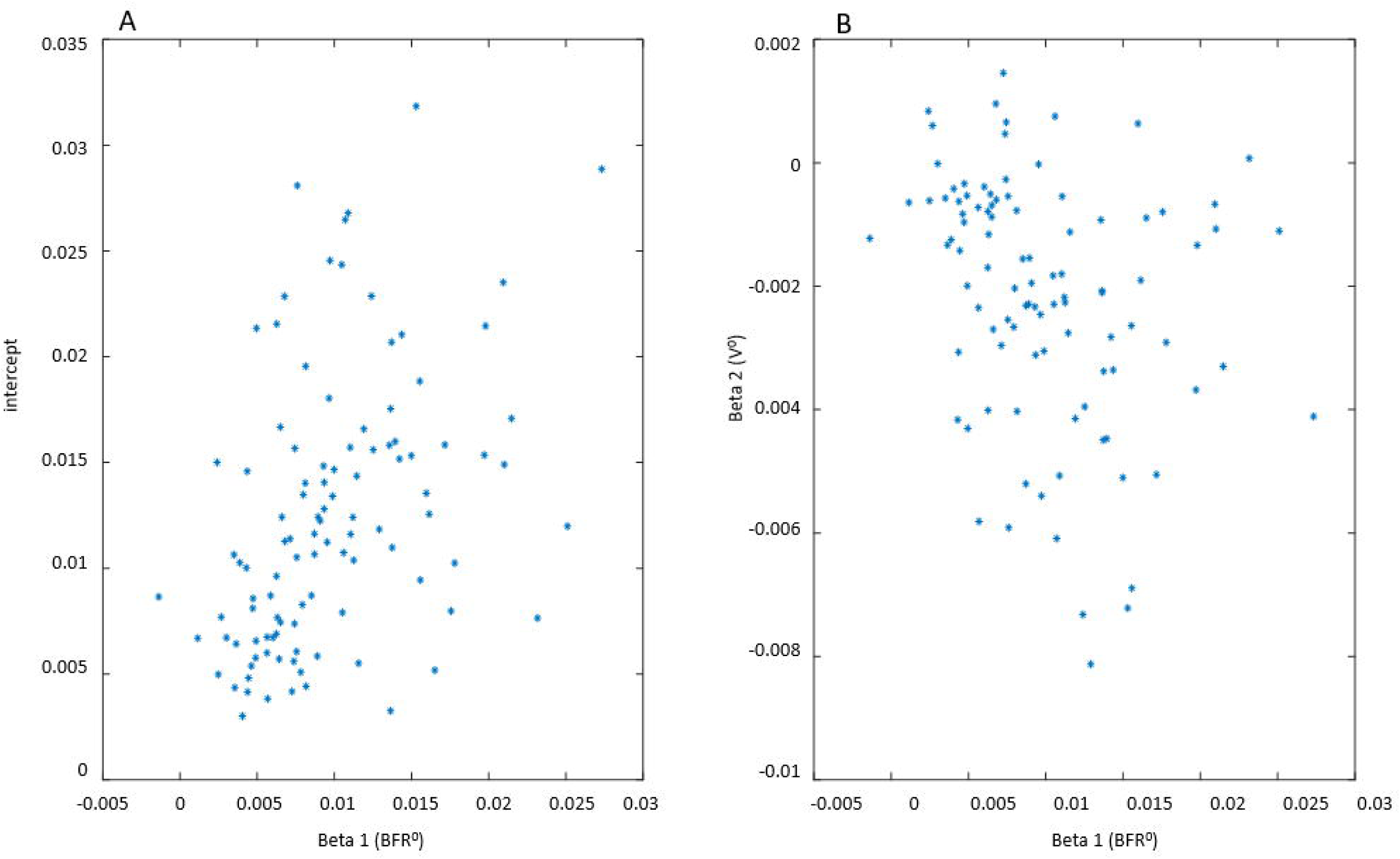
Scatter plots of intercept and two model coefficients for 124 multivariable linear regression models of UrgR. Figure 13A: intercept versus β_1_ (R=0.47, p<0.001). Figure 13B: ß2 versus ß1 (R=-0.33, p<0.001).

Of 21 correlations between 7 coefficients of the models, 12 are significant (Table 8) but the majority of them are weak to moderate. Of main interest are positive correlations between intercept and sensitivity to BFR (β_1_), and negative correlation between intercept and ß_2_ and β_3,_ indicating that participants with higher UrgR are more sensitive to BFR, less sensitive to urine volume, and have larger difference between day and night UrgRs. Correlations between β_i_ and β_ik_, and β_ij_ and β_ki_ are stronger; for instance, strong negative correlation of ß_2_ and ß_23_ indicates that people more sensitive to urine volume are also more sensitive to it during the night than during the day. Overall, the complex structure of correlations between coefficients of the multivariable linear regression models of UrgR indicates the possibility of different mechanisms of urge in different people. It is not clear (due to a limited sample size, n=124) if it is possible to identify distinct subtypes of patients with urgency based on these data, or if it is a continuum of different levels of sensitivities to various factors. Subtyping of patients with urgency would benefit from inclusion of data outside of BDs (patient-reported symptoms, physical examination, etc.) and would require consideration of the correlations across these variables [35]. Such study is of interest but is beyond the scope of the current paper.

**Table 8.** Significant inter-subject correlations between coefficients of UrgR models for 124 participants.

| First coefficient | Second coefficient | Correlation R | p-value |
| --- | --- | --- | --- |
| intercept | $\beta_1$ | 0.47 | <0.001 |
| intercept | $\beta_2$ | -0.53 | <0.001 |
| intercept | $\beta_3$ | -0.56 | <0.001 |
| intercept | $\beta_{23}$ | 0.21 | 0.014 |
| $\beta_1$ | $\beta_2$ | -0.33 | <0.001 |
| $\beta_1$ | $\beta_{12}$ | -0.22 | 0.013 |
| $\beta_1$ | $\beta_{13}$ | -0.57 | <0.001 |
| $\beta_1$ | $\beta_{23}$ | 0.25 | 0.015 |
| $\beta_2$ | $\beta_3$ | 0.48 | <0.001 |
| $\beta_2$ | $\beta_{23}$ | -0.79 | <0.001 |
| $\beta_3$ | $\beta_{23}$ | -0.48 | <0.001 |

#### Comparison of participants with satisfactory and unsatisfactory models

As described above, models of urine production rate and UrgR for 124 (63% of 197 participants) satisfied the selection criteria and were used for further analysis and simulation. It is of interest to evaluate how these 124 participants differ from 73 participants for whom satisfactory models were not created. Table 9 presents the comparison of the mean values and ranges of the BD variables for these two groups of participants. Variables are sorted starting with those with the most significant differences between the groups. Participants for whom satisfactory models were not created demonstrated significantly higher maximum across 3-day values of time-dependent instantaneous urinary frequency F(t) (Max F). They consumed more alcohol and high osmolality drinks. This demonstrates that our models have some difficulties in predicting extremely high peaks of F and in predicting the effects of alcohol and high-osmolality drink consumption. On the other hand, there are no differences in total intake and void volumes, maximum urge, number of voids and leaks, and sex of the participants. Differences between the groups in caffeine content of the drinks and mean urge levels are borderline significant.

**Table 9.** Comparison of the groups of participants with satisfactory (selected) versus unsatisfactory (filtered) models.

| Variables | Mean<br>Selected | Std<br>Selected | Min<br>Selected | Max<br>Selected | Mean<br>Filtered | Std<br>Filtered | Min<br>Filtered | Max<br>Filtered | p-value | Adjusted<br>p-value |
| --- | --- | --- | --- | --- | --- | --- | --- | --- | --- | --- |
| Max F, 1/day | 62.4 | 64.0 | 7.0 | 480.0 | 114.8 | 127.9 | 12.1 | 720.0 | <0.001 | 0.003 |
| ALC, mL | 19.1 | 44.9 | 0.0 | 215.2 | 75.7 | 212.2 | 0.0 | 1632.5 | 0.005 | 0.022 |
| OSM, mOsm | 175.4 | 165.6 | 0.0 | 934.1 | 252.7 | 220.2 | 1.2 | 895.4 | 0.006 | 0.022 |
| Mean URG | 1.9 | 0.8 | 0.8 | 3.9 | 1.7 | 0.7 | 0.0 | 3.8 | 0.026 | 0.083 |
| CAF, mg | 318.5 | 278.6 | 0.0 | 1300.0 | 410.7 | 311.4 | 0.0 | 1450.0 | 0.032 | 0.086 |
| Sum intake, mL | 5529.2 | 2246.1 | 1567.4 | 13722.3 | 6084.2 | 2437.0 | 2410.3 | 13101.3 | 0.105 | 0.240 |
| BFR ratio | 2.6 | 1.4 | 1.0 | 11.8 | 2.5 | 1.1 | 1.1 | 7.4 | 0.594 | 0.849 |
| Sum void, mL | 5299.3 | 2311.1 | 828.1 | 14195.5 | 5471.6 | 2261.5 | 1160.8 | 11740.9 | 0.609 | 0.849 |
| PVR, mL | 15.4 | 15.0 | 0.0 | 50.0 | 16.2 | 15.5 | 0.0 | 50.0 | 0.711 | 0.849 |
| Max void, mL | 462.9 | 212.8 | 103.5 | 1360.4 | 475.0 | 234.9 | 118.3 | 1419.6 | 0.712 | 0.849 |
| Max URG | 3.0 | 1.0 | 1.0 | 4.0 | 3.1 | 1.0 | 0.0 | 4.0 | 0.719 | 0.849 |
| LEAK | 2.0 | 4.3 | 0.0 | 26.0 | 2.2 | 4.8 | 0.0 | 33.0 | 0.743 | 0.849 |
| N voids | 25.9 | 8.9 | 8.0 | 51.0 | 26.2 | 7.8 | 12.0 | 50.0 | 0.861 | 0.918 |
| male, % | 47.6 |  |  |  | 55.4 |  |  |  | 0.999 | 0.999 |

### Simulation of behavioral modifications by using the developed models

We used the developed 124 satisfactory models of UrgR to simulate the effects of behavioral modifications. Of these participants with satisfactory models, 20 did not consume caffeine or alcohol (noCA), 29 consumed caffeine but not alcohol (yCnoA), 3 consumed alcohol but not caffeine (AnoC), and 72 consumed both caffeine and alcohol (yCA). Of these 124 participants, 106 had at least one urgency episode; distribution of these participants “with urgency” across “consumer groups” was the following: noCA=17, yCnoA=23, yAnoC=3, yCA=63. Since the goal of behavioral modifications is to minimize urgency episodes, we investigated the effects of behavioral modifications only on 106 participants with urgency. Two metrics of the effects of the modifications are of interest: first, the reduction of the mean (across the 3 days of the diary) UrgR for each individual, and second, reduction of the number of urgency episodes for each individual. The first metric translates into the expected reduction in urinary frequency (assuming that voids occur at the same levels of urge as without modification). The second assumes that timing of voids is the same as without modifications, but urgency is not reached due to the reduced UrgR. Table 10 provides the values of both metrics (separated by semicolon) averaged within the above groups of consumers.

**Table 10.**
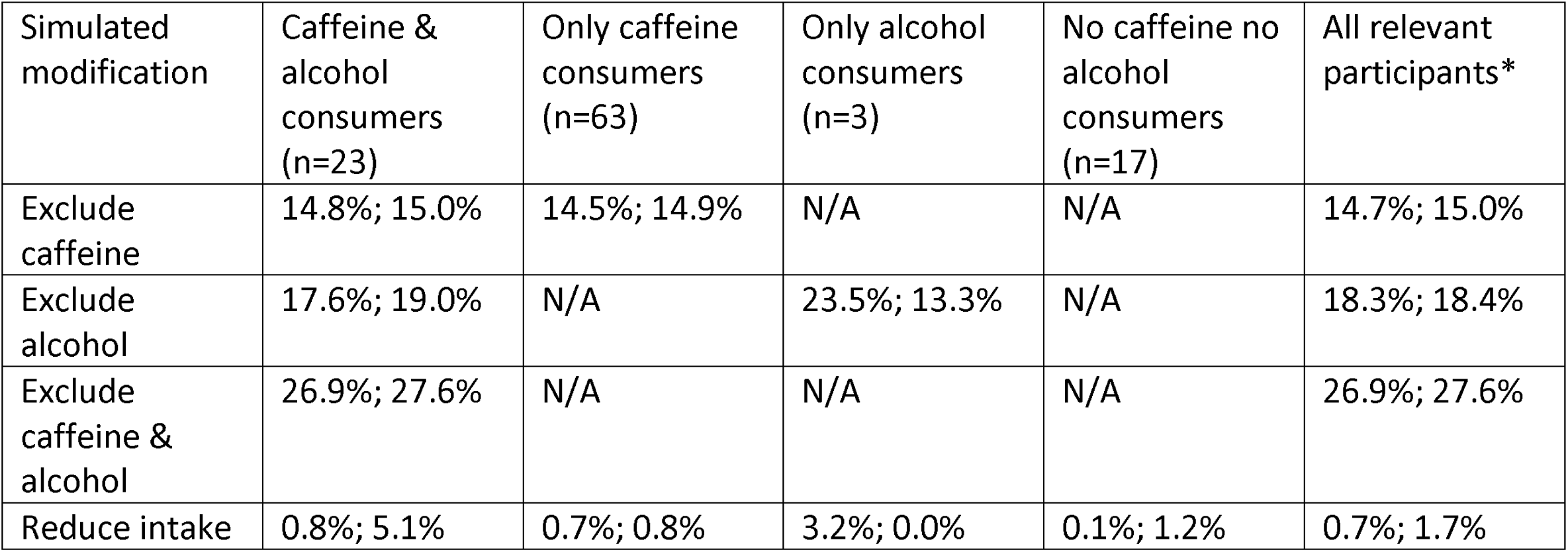
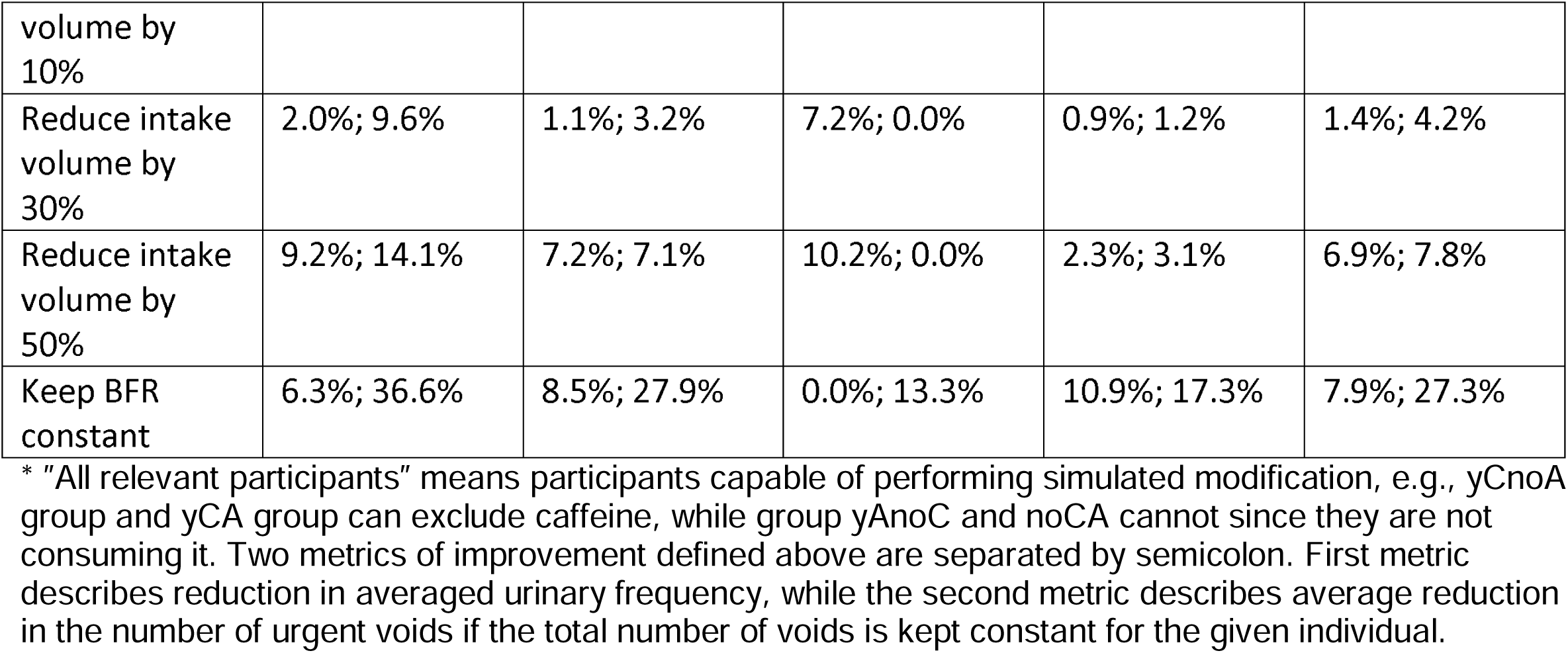
Improvements in mean UrgR and number of urgency episodes due to behavioral modifications.

Simulated exclusion of caffeine and alcohol predicted much higher effect in reduction of mean UrgR and reduction in the number of urgency episodes than reduction of intake volume. Exclusion of both caffeine and alcohol is more effective than exclusion of one of them. The most effective (in terms of reduction in the number of urgency episodes) simulated modification is presented in the last row of Table 10. It is the hypothetical case where BFR and urine volume in the bladder are somehow kept constant and equal to the median values of BFR and urine volume during the 3 days of the BD. Note that, in this case, the total intake volume is not reduced; however, the timings of intakes should be modified. Especially high is the reduction of the number of urgency episodes in all four groups of participants. The mean reduction in the number of urgency episodes is as high as 36.6% for participants consuming caffeine and alcohol and is 27.3% for all four groups on average. This result is consistent with our observation that peaks of UrgR are collocated with peaks of BFR (Figure 2 and Supplemental Figures S1-S9). It is also consistent with the strong intra-subject correlation of UrgR with BFR (R>0.7) in Table 4, and with the high level of similarity of the UrgR(t) and BFR(t) profiles demonstrated with cross-correlation functions (Figure 7 and Supplemental Figure S10).

Reactions of participants to the simulated modifications vary across individuals. For some participants, modifications resulted in more than 50% improvement in the number of urgency episodes, while for others, they resulted in no improvement at all. Histograms in Figure 14 present the distributions of relative improvement in the number of urgency episodes across participants capable of simulated modifications (see Table 10 footnote and Figure 14 caption). Simulated cases of excluded caffeine (Figure 14A), excluded alcohol (Figure 14B), intake volume reduced by 50% (Figure 14C), and a hypothetical case of constant BFR (Figure 14D) are presented. Keeping BFR constant appears to be the most effective simulated modification at the individual level as well; i.e., 23 (21.7%) of participants are predicted to experience more than 50% reduction in the number of urgency episodes, while for other modifications, 50% reduction was achieved for a much lower number and percentage of relevant participants: caffeine exclusion 8 (9.3%), alcohol exclusion 3 (11.5%), and 50% intake volume reduction 3 (2.8%).

**Figure 14.**
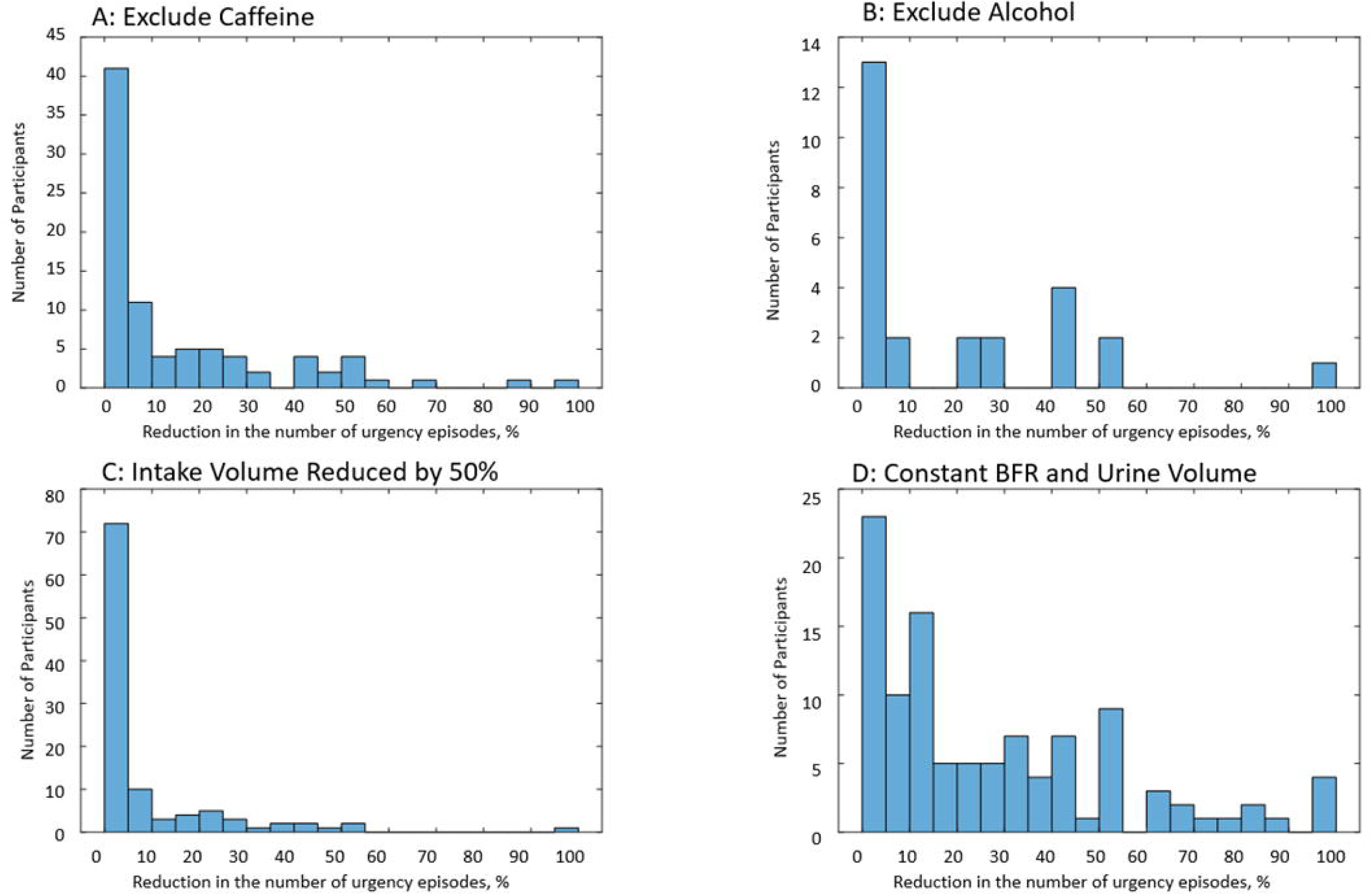
Relative improvement in the number of urgent voids due to simulated behavioral modifications. Figure 14A: caffeine excluded in groups of participants with urgency consuming caffeine without alcohol (n=63) and caffeine and alcohol (n=23). Figure 14B: alcohol excluded in groups of participants with urgency consuming caffeine and alcohol (n=23) and alcohol without caffeine (n=3). Figure 14C: intake volumes reduced by 50% without changing the timing of voids in all patients with urgency (n=106). Figure 14D: simulated BFR and urine volume are kept constant and equal to the median values of BFR and urine volume during 3 days (n=106).

To determine what kinds of participants benefit more from the simulated modifications, we calculated inter-subject correlations of the above two metrics of improvement with the following characteristics of the individual participants: total intake volume, total void volume, total caffeine consumed, total alcohol consumed, mean osmolality of drinks, number of voids, number of leaks, ratio of maximum BFR to median BFR, mean level of urge at voids, mean UrgR, maximum UrgR. We calculated the above correlations for each modification and group of participants separately. The decrease in percentage of urgency episodes due to excluding caffeine (in the group consuming caffeine and both caffeine and alcohol n=23+63) was found to be correlated with total amount consumed caffeine (R=0.21, p=0.06) and with urge level at voids

(R=-0.36, p=0.0008), indicating that patients with moderate urgency and high caffeine consumption would benefit from caffeine exclusion. The decrease in percentage of urgency episodes due to excluding alcohol (in the group consuming alcohol and both alcohol and caffeine n=3+63) was found to be correlated with the ratio of maximum BFR to median BFR (R=0.48, p=0.014), confirming the role of alcohol in creating peaks of BFR and UrgR, which can be avoided by excluding alcohol. The decrease in the percentage of urgency episodes due to decreasing the intake volume by 50% (in all patients with urgency n=106) was found to be correlated with the total void volume (R=0.24, p=0.015), mean osmolality of drinks (R=0.24, p=0.014), and negatively correlated with the mean urge level (R=-0.26, p=0.007). The decrease in the mean UrgR due to 50% intake reduction was correlated with the total void volume (R=0.30, p=0.002), maximum voided volume (R=0.24, p=0.014), and negatively correlated with consumed alcohol (R=-0.16, p=0.09). The reduction in the number of urgency episodes was negatively correlated with mean urge level at voids (R=-0.38, p=0.0001) and with the number of leaks (R=-0.27, p=0.057), indicating that this modification is more beneficial for patients with moderate level of urge and not too frequent episodes of incontinence. The above hypothetical case is likely not easy to implement for the majority of patients, it rather serves as an asymptotic solution demonstrating what is the maximum potential improvement in the percentage of urgency episodes that could be accomplished without decreasing the volume of drinks and without changing timings of the voids. It also provides some insights for selection of patients for whom such approach might be helpful, i.e., patients with moderate mean urge at void level and with rare incontinence episodes. One consistent observation can be made for all the above simulations, i.e., intake modifications seem to be beneficial for patients with moderate urgency. For these patients, the most beneficial medication could be the exclusion of caffeine and alcohol and/or modification of timings of the drinks to avoid peaks in BFR. Additionally, for participants with higher-level urinary urgency, multiple episodes of urgency, and incontinence, none of the simulated behavioral modifications was helpful.

## Discussion

The main premise of this paper is that BDs allow much more than just verification or more accurate presentation of the data similar to that available through the relevant self-reported responses on questionnaires. We believe that important additional information and insights could be extracted through the analysis of voiding and drinking patterns, where not only volumes of the voids and intakes and voiding sensations are recorded, but importantly, accurate timings of these events. To implement this program of pattern analysis, we proposed and developed a dynamic approach to the analysis of BDs, including seven steps or levels: 1) graphical representation and visual inspection of the patterns of intakes, voids, and sensations; 2) introduction of dynamic variables, such as BFR, UrgR, and instantaneous time-dependent frequency (reciprocal of time intervals between voids); 3) intra-subject correlations between the dynamic variables; 4) analysis of similarities and delays between patterns of dynamic variables using cross-correlation function; 5) grey-box models allowing for prediction of the BFR in an individual, given the types, volumes, and timings of intakes; 6) multivariable linear regression models predicting individual’s UrgR from their BFR and urine volume; 7) simulation of the effects of intake modifications on sensation of urge. Each next level provides deeper understanding of the relationship between patterns of intakes, sensations, and voids; however, each level requires more detailed and accurate data; therefore, the number of individuals to whom the proposed levels of analysis are applicable progressively reduced from 197 (levels1-4) to 145 (level 5) to 124 (levels 6-7). Consistency of the results established across the levels of analysis increases our confidence. There are two lines of inquiry and findings across all seven levels; the first is about the role of BFR as a driver of urge and frequency, while the second is about the role of caffeine and alcohol in stimulating BFR and therefore urinary urge and frequency.

Visual inspection of dynamic variables patterns, in particular BFR(t) (Figure 2, Supplemental Figures S1-S9) demonstrated that BFR could change dramatically during the day, and that peaks of BFR could be 10-12-fold higher than median BFR value for the given individual. Analysis of the distributions of max BFR/median BFR values confirmed this conclusion for the whole cohort (Figure 3). Furthermore, the patterns of instantaneous frequency (F(t)) and UrgR(t) demonstrated high peaks, which were collocated with peaks of BFR(t) (Figure 2, Supplemental Figures S1-S9). Analysis with cross-correlation function (Figure 7, Supplemental Figure S10) demonstrated a high level of similarity and absence of delay between BFR(t), UrgR(t), and F(t). Strong association of these patterns is corroborated by the high values of mean intra-subject correlation coefficients between BFR, UrgR, and F (Table 4) for full correlations and partial correlations calculated with fixed urine volume and IR(t). Strong positive correlations of UrgR and F with BFR were observed for the majority of individuals in the cohort, while correlations of UrgR and F with urine volume were mostly weak to moderate, with positive and negative correlations being almost equally prevalent (Figure 6). Consistent strong positive correlation of UrgR and F with BFR indicates that BFR rather than urine volume was the main determinant of urgency. The hypothetic mechanism is that BFR drives the urge growth rate and makes a person void when urge reaches certain level. One can argue that increased BFR increases the volume of urine in the bladder, and a person voids as soon as the certain fraction of the bladder volume is filled. This reasoning, however, contradicts the observation that multiple “very urgent” voids (URG≥3) occur when only a small fraction of the bladder is filled with urine (Figure 4D-4E) and the observation that the same level of urge in an individual could occur at different levels of bladder fullness (Figure 4F). Multivariable linear regression models of UrgR corroborated the conclusion of BFR being the dominant driver of UrgR and urge level; it also revealed that UrgR for most of the participants decreases with urine volume, meaning that urge is growing slower than linearly with urine volume. This result is in agreement with the observations of [27], where the reported urge during the natural bladder-filling experiments demonstrated sigmoidal dependence on urine volume. Further analysis of the models showed that participants with more frequent urgency episodes are more sensitive to the increased BFR. Simulation of the hypothetical behavioral modification of keeping BFR as constant as possible showed that it resulted in the more effective reduction of the number of urgency episodes than reduction of intake volume or just exclusion of caffeine and/or alcohol. Thus, results of the analysis at multiple levels consistently demonstrated the role of the peaks of BFR as drivers of urge and the benefit of the measures leading to avoidance of such peaks.

The results of our analyses are in concert with conclusions of the study by Redmond et al [36] of BDs of 24 women with OAB compared with 40 controls, where BFR was an independent predictor of the percentage of urgent voids in patients with OAB, but not in the control group. In that study, VVs and BFRs were stratified into three levels: low, medium, and high. All voids within the groups of women were combined together, ignoring possible heterogeneity of the individuals, of which the groups consist, including different bladder capacities. Our dynamic analysis was performed separately for each individual and is free of these limitations. Another study in concert with our observations reports on the bladder sensations in normal (n=12) and OAB (n=17) participants during the oral hydration by rapid consumption of 2L of Gatorade [37]. One conclusion was “this study showed fast filling can lead normal individuals to experience OAB sensations because sensation event patterns in normal participants during fast filling were similar to OAB participants during slow filling.” Although tempting to view this conclusion as a confirmation of our observations, it is worth noting that mean BFR was about 6.5 mL/min during “slow filling” and over 12 mL/min during “fast filling”.

While “fast filling” BFR is similar to observed during the peaks of BFR in our BDs, the “slow filling” BFR was about 6-fold higher than mean BFR (∼1 mL/min) across the 3-day BDs in our study. It would be of interest to see the results of oral hydration study with slower “slow filling”; in this case, differences in bladder sensations during “slow” and “fast” filling might be even higher than observed in [37].

Caffeine- and alcohol-containing drinks were reported as drivers of diuresis and urge [38,39]. Our second line of inquiry is about the role of caffeine and alcohol. Visual inspection (Figure 2, Supplemental Figures S1-S9) demonstrated that peaks of BFR are often collocated with intakes of caffeine- and alcohol-containing drinks. The assumption that certain peaks of BFR are caused not just by the intake volume but by the caffeine and alcohol content of the drinks was corroborated by the better fit to the data of the grey-box models of urine production rate, taking composition of the drinks into account relative to the models ignoring caffeine and alcohol content (Figure 8). Simulated exclusion of both caffeine and alcohol in a group of participants consuming them predicted to reduce by nearly 30% both the mean UrgR and the number of urgent voids. The effect of caffeine and alcohol exclusion is predicted to be much more substantial than that of intake volume reduction.

In summary, simulation predicted that the most beneficial modifications are those resulting in smooth profiles of BFR, which can be accomplished by exclusion of caffeine and alcohol and/or other measures (e.g., increasing number and decreasing volumes of intakes), which is consistent with some published recommendations [40]. It also demonstrated that, for some participants with severe and frequent urgency episodes, none of the simulated modifications was helpful. Such participants are likely beyond the behavioral modifications stage and require more radical medical treatment.

The above distinction – together with observed differences in the dependence of UrgR on urine volume, i.e., positive and negative intra-subject correlations of UrgR and F with VV (Figure 6), and both positive and negative values of coefficient ß_2_ (V_0_) of the linear regression model (Figure 13) – indicate that urinary urge might have different mechanisms in different participants. It is in line with the observation of urge dynamics in the natural-filling study, where some participants demonstrated sigmoidal and some linear dependencies of urge on the urine volume [27]. Recently, an expert panel [41] recommended to include some BD variables together with the results of physiological tests (e.g., maximum uroflow and PVR) into the classification of OAB patients. We believe that adding variables derived from the BDs using a dynamic approach, such as coefficients β_1_(BFR^0^) and β_2_(V^0^) might improve classification and provide additional insights into the mechanism of urge in various phenotypes.

Our paper is not free of limitations. The first group of limitations is related to the collected, or rather to the non-collected data; the second to the methodology and model assumptions. BDs are not perfect, there is always a chance of some intakes or voids being missed or recorded incorrectly [42]. We do not have data on the physical activity and heart rate that affects glomerular filtration rate. We do not have data on salt consumption with food that affects osmolality of the blood and, therefore, water reabsorption rate. We do not have data about fluids consumed with food (e.g., soup, watermelon). Participants reported the types of drinks, while information on alcohol and caffeine content is based on the literature data that is not necessarily accurate for all individual drinks. The absence or inaccuracy of these data negatively affects the accuracy of our models of urine production rate. We minimized the effects of these inaccuracies in the linear regression model of UrgR by using BFR(t) extracted from the BDs. The simulated changes in BFR(t) due to behavioral modifications were estimated as the difference in urine production rates predicted by the grey-box models with and without the above modifications, thus cancelling out some of the above inaccuracies.

The main assumption in defining the dynamic variables (BFR(t), F(t), and UrgR(t) was that they are constant during the time intervals between voids and change abruptly at the time of voids. This is accurate for F(t), defined as reciprocal of time interval between voids, but it is an approximation for BFR(t) and UrgR(t), which is the best estimation, given the data available in the BDs. Better approximation for BFR(t) may be derived from future studies where an observed rate growth of urine volume during natural bladder-filling (e.g., through ultrasound or fMRI measurements [43,44]) would be combined with BDs for the same participants. Similarly, accurate reporting of the level of urge during natural bladder-filling experiments [25] combined with BDs for the same patients could lead to better approximations of UrgR(t) profiles and more accurate predictions.

Despite the above limitations and approximations, our findings at different levels do corroborate each other. Deeper levels of analysis require more assumptions and approximations to provide more details on the codependence of the patterns of intakes, sensations, and voids, but are, in general, in concert with the visual observations at the first-level analysis, therefore increasing confidence in the obtained results. Obviously, our results should not be overgeneralized. They need to be validated in an independent cohort and require clinical verification. One possible way to evaluate clinical significance of dynamic approach to BD data is to start with implementing just the first step, i.e., graphically represent patterns of intakes, sensations, and voids, together with BFR and UrgR (Figure 2, Supplemental Figures S1-S9). This visual representation is useful since it clearly demonstrates the presence of peaks of BFR and urge growth and their collocation with certain intakes, which would enable both clinician and patient to generate and test hypotheses on the causes of urgency episodes, and propose and test behavioral modifications. Realized as a mobile application, it could be a useful educational and self-improvement tool for the patients.

Advancement and the growth of popularity of wearable devices measuring physical activity, together with the development of numerous apps for recording bladder and food diaries, generate optimism with regard to improvement of the quality of data required for dynamic analysis. Another potential way to advance the accuracy of model predictions is to collect data on urine composition. Studies have shown that urine osmolality can be reliably estimated by comparing the color of the urine with color charts [45]. Availability of such data would allow for fitting the model of urine production rate, not only by comparing model predictions with the BFR, but also with the osmolality profiles. Similarly advantageous would be at least occasional measurements of caffeine and alcohol concentrations in urine. Having both measured and simulated urine composition profiles would advance the accuracy of the UrgR models and could generate more insights potentially useful for clinical practice.

## Conclusion

A dynamic approach to the analysis of bladder diaries was proposed and developed, including visual representation of the patterns of intakes, bladder sensations, and voids, as well as analyzing intra-subject correlations of the introduced dynamic variables (bladder filling rate and urgency growth rate), and predictive modeling of these variables. The main results of the dynamic analysis performed on the 3-day BDs from 197 participants with LUTS include observations of the collocation of the BFR and UrgR peaks with the timings of intakes containing caffeine and alcohol and with the high-volume intakes. For a majority of participants, urgent voids were observed when only a small portion of their bladder was filled with urine. UrgR was strongly positively correlated with the BFR in the majority of participants, indicating that BFR, rather than urine volume, was the main determinant of their urge sensations. A dynamic approach to analysis of BDs and its results should be validated in an independent cohort. Although the results of the analysis should not be generalized prior to validation, the dynamic approach itself might be applied to the analysis of other physiological processes of interest where continuous measurements of the variables are available, as in continuous glucose or continuous heart rate monitoring and where the rate of change of the variables in response to the stimuli might be as important or even more important than their absolute values.

## Data Availability

The data that support the findings of this study are openly available in the NIDDK Central Repository at https://repository.niddk.nih.gov/. Please reference the acronym LURN.

## Acknowledgements

Heather Van Doren, Senior Medical Editor with Arbor Research Collaborative for Health, provided editorial assistance on this manuscript.

Authors are grateful to Drs. Ziya Kirkali, Jerry G. Blaivas, Jeffrey P. Weiss, Claire C. Yang, Robert M. Merion, Bruce M. Robinson, and Brenda W. Gillespie for interesting discussions.

The authors acknowledge the LURN study group for BD data. This LURN Ancillary Study is funded by the National Institutes of Health (NIH), grant number R21DK121065. LURN Data Coordinating Center (Arbor Research Collaborative for Health) and a clinical site at the University of Michigan are associated with this LURN Ancillary study.

The LURN study is supported by the National Institute of Diabetes & Digestive & Kidney Diseases through cooperative agreements (grants DK097780, DK097772, DK097779, DK099932, DK100011, DK100017, DK099879).

The following individuals were instrumental in the planning and conduct of LURN study at each of the participating institutions:

Duke University, Durham, North Carolina (DK097780): PIs: Cindy Amundsen, MD, J. Eric Jelovsek, MD; Co-Is: Kathryn Flynn, PhD, Jim Hokanson, PhD, Aaron Lentz, MD, David Page, PhD, Nazema Siddiqui, MD, Lisa Wruck, PhD, Todd Harshbarger, PhD; Study Coordinators: Paige Green, MSc, Magaly Guerrero, BSc, Stephanie Yu, Summer Granger

University of Iowa, Iowa City, IA (DK097772): PIs: Catherine S Bradley, MD, MSCE, Karl Kreder, MD, MBA; Co-Is: Bradley A. Erickson, MD, MS, Daniel Fick, MD, Vince Magnotta, PhD, Philip Polgreen, MD, MPH; Study Coordinators: Jean Walshire, AAS, Rachel Setting, BA

Northwestern University, Chicago, IL (DK097779): PIs: James W Griffith, PhD, Kimberly Kenton, MD, MS, Brian Helfand, MD, PhD; Co-Is: Carol Bretschneider, MD, David Cella, PhD, Sarah Collins, MD, Julia Geynisman-Tan, MD, Alex Glaser, MD, Christina Lewicky-Gaupp, MD, Margaret Mueller, MD, Francesca Farina, PhD, Richard Fantus, MD, Devin Boehm, BS; Study Coordinators: Hosanna An, Andrea Villegas, Melissa Marquez, MBA, Malgorzata Antoniak, PhD, Pooja Talaty, MS, Sophia Kallas, Jessica Thomas, Amelia Joblin. Dr. Helfand and Ms. Talaty are at NorthShore University HealthSystem.

University of Michigan Health System, Ann Arbor, MI (DK099932): PI: J Quentin Clemens, MD, FACS, MSCI; Co-Is: John DeLancey, MD, Dee Fenner, MD, Rick Harris, MD, Steve Harte, PhD, Anne P. Cameron, MD, Aruna Sarma, PhD, Giulia Lane, MD, Priyanka Gupta, MD, Whitney Horner, MD; Jannah Thompson, MD; Payton Schmidt, MD; Study Coordinators: Greg Mowatt, BA, Sarah Richardson, BS, Lydia Duong, Nailah Henry, BA.

University of Washington, Seattle Washington (DK100011): PI: Claire Yang, MD; Co-I: Anna Kirby, MD; Study Coordinators: Brenda Vicars, RN; Sreya Gutta.

Washington University in St. Louis, St. Louis Missouri (DK100017): PI: H. Henry Lai, MD; Co-Is: Joshua Shimony, MD, PhD, Fuhai Li, PhD; Study Coordinators: Linda Black, RN, Vivien Gardner, BSN, Patricia Hayden, BSN, Diana Wolff, Aleksandra Klim, RN, MHS, CCRC

Arbor Research Collaborative for Health, Data Coordinating Center (DK099879): PI: Robert Merion, MD, FACS; Co-Is: Victor Andreev, PhD, DSc, Brenda Gillespie, PhD, Abigail Smith, PhD; Project Manager: Jessica Durkin, M.Ed, MBA; Clinical Monitor: Melissa Sexton, BA, CCRP; Research Analysts: Margaret Helmuth, MA, Sarah Mansfield, MS. Project Associate: Julia Nashif, BA

National Institute of Diabetes and Digestive and Kidney Diseases, Division of Kidney, Urology, and Hematology, Bethesda, MD: Project Scientist: Ziya Kirkali MD; Project Officer: Christopher Mullins PhD; Project Advisor: Julie Barthold, MD

## Supporting Information

See separate Supplemental Materials document.

### Supplemental Material Text (Section Headings)

Urine Formation Rate Model

Supplemental References

### Supplemental Figures (Captions)

**Supplemental Figure S1.** Intake and voiding profiles recorded in the 3-day BD of participant B and dynamic variables derived from the profiles.

**Supplemental Figure S2.** Intake and voiding profiles recorded in the 3-day BD of participant C and dynamic variables derived from the profiles.

**Supplemental Figure S3.** Intake and voiding profiles recorded in the 3-day BD of participant D and dynamic variables derived from the profiles.

**Supplemental Figure S4.** Intake and voiding profiles recorded in the 3-day BD of participant E and dynamic variables derived from the profiles.

**Supplemental Figure S5.** Intake and voiding profiles recorded in the 3-day BD of participant F and dynamic variables derived from the profiles.

**Supplemental Figure S6.** Intake and voiding profiles recorded in the 3-day BD of participant G and dynamic variables derived from the profiles.

**Supplemental Figure S7.** Intake and voiding profiles recorded in the 3-day BD of participant H and dynamic variables derived from the profiles.

**Supplemental Figure S8.** Intake and voiding profiles recorded in the 3-day BD of participant I and dynamic variables derived from the profiles.

**Supplemental Figure S9.** Intake and voiding profiles recorded in the 3-day BD of participant J and dynamic variables derived from the profiles.

**Supplemental Figure S10.** Cross-correlation functions between dynamic BD variables of participants B-J.

**Supplemental Figure S11.** Box plots for the parameters of the urine formation rate models for 145 participants.

## References

1. Hofmeester I. Moderate agreement between bladder capacity assessed by frequency volume charts and uroflowmetry, in adolescent and adult enuresis patients. Neurourol Urodyn 2017;36:745–747.

2. Brown J. Measurement characteristics of a voiding diary for use by men and women with overactive bladder. Urol 2003;61(4):802–809.

3. Blaivas JG, Tsui JF, Amirian M, Ranasinghe B, Weiss JP, Haukka J, et al. Relationship between voided volume and the urge to void among patients with lower urinary tract symptoms, Scandinavian J Urol 2014;48(6):554–558.

4. Jimenez-Cidre MA. The 3-day bladder diary is a feasible, reliable and valid tool to evaluate the lower urinary tract symptoms in women. Neurourol Urodyn 2015;34:128–132.

5. Groutz A. Noninvasive outcome measures of urinary incontinence and lower urinary tract symptoms: a multicenter study of micturition diary and pad tests. J Urol 2000;164:698–701.

6. Ku J. Voiding diary for the evaluation of urinary incontinence and lower urinary tract symptoms: Prospective assessment of patient compliance and burden. Neurourol Urodyn 2004;23:331–335.

7. Homma Y. Assessment of overactive bladder symptoms: Comparison of 3-day bladder diary and the overactive bladder symptoms score. Urol 2011;77: 60–64.

8. Song M. Correlation of the overactive bladder symptom score, and the voiding diary and urodynamic parameters in patients with overactive bladder syndrome. LUTS 2014;6:180–184.

9. Brummen HJ. The association between overactive bladder symptoms and objective parameters from bladder diary and filling cystometry. Neurourol Urodyn 2004;23:38–42.

10. DiBona GF. Physiology in perspective: The wisdom of the body. Neural control of the kidney. The Walter B. Cannon Memorial Award Lecture. Am J Physiol Regul Integr Comp Physiol 2005;289: R633–R641.

11. Fontecave-Jallon J, Randall Thomas S. Implementation of a model of bodily fluids regulation. Acta Biotheoretica 2015;63(3):269–282.

12. Karaaslan F, Denizhan Y, Kayserilioglu A, Ozcan Gulcur H. Long-term mathematical model involving renal sympathetic nerve activity, arterial pressure, and sodium excretion. Ann Biomed Engineer 2005;33(11):1607–1630.

13. Thomas SR, Layton AT, Layton HE, Moore LC. Kidney modeling: Status and perspectives. Proceedings of the IEEE April 2006;94(4).

14. Guyton AC, Taylor AE, Granger HJ. Dynamics and control of body fluid. Philadelphia, PA: WB Saunders Company; 1975.

15. Layton AT, Edwards A. Mathematical modeling in renal physiology. NY: Springer; 2014.

16. Weinstein AM. Mathematical models of renal fluid and electrolyte transport: Acknowledging our uncertainty. Am J Physiol Renal Physiol 2003;284:F871–F884.

17. Yang CC, Weinfurt KP, Merion RM, Kirkali Z; LURN Study Group. Symptoms of lower urinary tract dysfunction research network. J Urol 2016;196:146.

18. Cameron AP, Lewicky-Gaupp C, Smith AR, Helfand BT, Gore JL, Clemens JQ, et al. Baseline lower urinary tract symptoms in patients enrolled in LURN: a prospective, observational cohort study. J Urol 2018;199:1023.

19. Bright E, Cotterill N, Drake M, Abrams P. Developing and validating the international consultation on incontinence questionnaire bladder diary. Eur Urol 2014:1–7.

20. Cameron AP, Wiseman JB, Smith AR, Merion RM, Gillespie BW, Bradley CS, et al. Are three-day voiding diaries feasible and reliable? Results from the Symptoms of Lower Urinary Tract Dysfunction Network (LURN) cohort. Neurourol Urodyn 2019;38(8):2185–2193.

21. McCusker RR, Goldberger BA, Cone EJ. Caffeine content of energy drinks, carbonated sodas, and other beverages. J Analytical Toxicology 2006;30:112–114.

22. Mettler S, Rusch C, Colombani PC. Osmolality and pH of sport and other drinks available in Switzerland. Schweizerische Zeitschrift für Sportmedizin und Sporttraumatologie 2006;54(3):92–95.

23. Coyne KS, Payne C, Bhattacharyya SK, Revicki DA, Thompson C, Corey R, et al. The impact of urinary urgency and frequency on health-related quality of life in overactive bladder: results from a national community survey. Value Health 2004;7(4):455–463.

24. FitzGerald MP, Butler N, Shott S, Brubaker L. Bother arising from urinary frequency in women. Neurourol Urodyn 2002;21(1):36–41.

25. Nagle AS, Speich JE, De Wachter SG, Ghamarian PP, Le DM, Colhoun AF, et al. Non-invasive characterization of real-time bladder sensation using accelerated hydration and a novel sensation meter: An initial experience. Neurourol Urodyn 2017;36(5):1417–1426.

26. Naimi HA, Nagle AS, Vinod NN, Kolli H, Sheen D, De Wachter SG, et al. An innovative, non-invasive sensation meter allows for a more comprehensive understanding of bladder sensation events: A prospective study in participants with normal bladder function. Neurourol Urodyn 2019;38(1):208–214.

27. De Wachter SG, Heeringa R, Van Koeveringe GA, Winkens B, Van Kerrebroeck PE, Gillespie JI. “Focused introspection” during naturally increased diuresis: description and repeatability of a method to study bladder sensation non-invasively. Neurourol Urodyn 2014;33(5):502–506.

28. Cow D. Diuresis. J Physiol 1914;48(1):1–17.

29. McCullagh P. Generalized linear models (2nd ed). UK: Routledge; 1983.

30. Peebles PZ. Probability, random variables, and random signal principles. NY: McGraw-Hill; 2001.

31. Bighamian R, Reisner AT, Hahn JO. A lumped-parameter subject-specific model of blood volume response to fluid infusion. Front Physiol 2016;7:390.

32. Janssen DAW, Schalken JA, Heesakkers JPFA. Urothelium update: how the bladder mucosa measures bladder filling. Acta Physiol 2017;220:201–217.

33. Bushman W, Blaivas J, Weiss J, Andreev VP. Physiologic determinants of urinary frequency and urgency in overactive bladder. Society for Biologic Research (SBUR) 2022 Annual Meeting; November 10-13, 2022; Orlando FL.

34. Linear regression models. Available at: https://people.duke.edu/~rnau/rsquared.htm. Accessed 2/2/23.

35. Andreev VP, Helmuth ME, Liu G, Smith AR, Merion RM, Yang CC, et al. Subtyping of common complex diseases and disorders by integrating heterogeneous data. Identifying clusters among women with lower urinary tract symptoms in the LURN study. PLOS ONE 2022;17(6):e0268547.

36. Redmond EJ, O’Kelly T, Flood HD. The effect of bladder filling rate on the voiding behavior of patients with overactive bladder. J Urol 2019;202(2):326–332.

37. Sebastian B, Swavely NR, Sethi D, Nagle AS, Thapa D, Vinod NN, et al. Comparison of sensation event descriptors in participants with overactive and normal bladders during non-invasive hydration studies. Arch Nephrol Urol Stud 2021;1(1):03.

38. Lohsiriwat S, Hirunsai M, Chaiyaprasithi B. Effect of caffeine on bladder function in patients with overactive bladder symptoms. Urol Ann 2011;3:14.

39. Eggleton MG. The diuretic action of alcohol in man. J Physiol 1942;101:172.

40. WebMD Editorial Contributors. Food and drink to tame an overactive bladder. Available at: http://www.webmd.com/urinary-incontinence-oab/food-drink. Accessed 12/21/22.

41. Blaivas JG, Li ESW, Dayan L, Edeson ME, Mathew J, O’Boyle AL, et al. Overactive bladder phenotypes: development and preliminary data. Can J Urol 2021;28(3):10699–10704.

42. Flynn KE, Wiseman JB, Helmuth ME, Smith AR, Bradley CS, Cameron AP, et al. Comparing clinical bladder diaries and recalled patient reports for measuring lower urinary tract symptoms in the symptoms of Lower Urinary Tract Dysfunction Research Network (LURN). Neurourol Urodyn 2022;41(8):1711–1721.

43. Pewowaruk R, Rutkowski D, Hernando D, Kumapayi BB, Bushman W, Roldán-Alzate A. A pilot study of bladder voiding with realtime MRI and computational fluid dynamics. PLOS ONE 2020;15(11):e0238404.

44. Nagle AS, Klausner AP, Varghese J, Bernardo RJ, Colhoun AF, Barbee RW, et al. Quantification of bladder wall biomechanics during urodynamics: A methodologic investigation using ultrasound. J Biomech 2017;61:232–241.

45. Kavouras SA, Johnson EC, Bougatsas D, Arnaoutis G, Panagiotakos DB, Perrier E, et al. Validation of a urine color scale for assessment of urine osmolality in healthy children. Eur J Nutr 2016;55(3):907–915.

