## Supplemental Materials for "Dynamic analysis of the individual patterns of intakes, voids, and bladder sensations reported in bladder diaries collected in the LURN study"

### Urine Formation Rate Model

#### Goal and foundation of the model

Our goal is to develop a parsimonious model capable of predicting urine formation rate from the bladder diary data (timings and volumes of drinks, as well as caffeine and alcohol content of drinks). Fluid entering the body is transported to the stomach and intestine, where it is absorbed into the blood. This added fluid is redistributed between blood and interstitial fluid to establish equilibrium, which is constantly disrupted by urine formation and new added fluid from drinks and food. Urine formation happening in the kidneys includes four stages: filtration, reabsorption, secretion, and excretion. Kidneys filter about 180 liters of blood plasma every day; only about 1.5 liters of urine is produced since more than 99% of filtered water is reabsorbed back to plasma in the renal tubules. Urine formation rate is a function of blood volume, osmolality, and concentrations of caffeine and alcohol. The exact percentage of not-reabsorbed water (~0.8%) and sodium strongly depends on blood osmolality and is regulated by arginine vasopressin (AVP) and atrial natriuretic peptide (ANP) hormones [1,2]. In addition to urine formation, some water is lost through perspiration and breathing. Data on physical activity, which affects perspiration, are not available in the current study and are not recorded in the food diaries. Therefore, we followed the usual approach and assumed volume of involuntary loss of water equal to volume of water consumed with solid food [3,4].

The core of our model is the modified model of fluid shift [5], which was originally developed to describe infusion and/or hemorrhage. According to [5], water added to blood (or removed from blood) is redistributed between blood and interstitial fluid to establish equilibrium ratio 1/(1+α), where α is around 3-5 and differs from person to person. The speed at which equilibrium is established is characterized by coefficient *K*, which also differs from person to person in the range about 0.03-0.08 min^-1^. Note that, in our model, water is added (drinks) and removed (urine) to and from blood multiple times, and there is not necessarily enough time for equilibrium to be established between the events of drinking and voiding. The equation describing dynamics of water redistribution is [5]:

$\frac{d\Delta V_{B}}{dt}=u\left( t \right)-v\left( t \right)-q(t)$	(s1)

Where $\Delta V_{B}=V_{B}-V_{B0}$ is the change of blood volume (mL), $u\left( t \right)$ – rate (mL/min) at which water is added to blood (in our case, absorbed from drinks in the stomach and intestine), $v\left( t \right)$ – rate (mL/min) at which water is removed from blood (in our case, as urine), $q(t)$ – describes redistribution (shift) of water between blood and interstitial fluid:

$q\left( t \right)=K\left\{ \Delta V_{B}-\frac{1}{1+\alpha}\cdot\int_{0}^{t} \left[ u\left( \tau\right)-v(\tau) \right]d\tau\right\}$	(s2)

Note that, in equilibrium: $\Delta V_{B}=\frac{1}{1+\alpha}\cdot\int_{0}^{te} \left[ u\left( \tau\right)-v(\tau) \right]d\tau$, assuming u(t_e_)-v(t_e_) = 0 at the moment of equilibrium t_e_. However, it is an intermediate equilibrium, since the global equilibrium is $\int_{0}^{t} \left[ u\left( \tau\right)-v(\tau) \right]d\tau=0$ (we urinate as much as we drink). The question is when, if ever, we reach the global equilibrium where:

$\frac{d\Delta V_{B}}{dt}$=0 and$V_{B}=V_{B0}$	(s3)

This question is of practical importance, since the answer determines how to define the initial equilibrium condition. In our model, we assumed that equilibrium is reached during nighttime sleep, although it might be not always the case.

Note that, in [5], both $u\left( t \right)$ and $v(t)$ are known inputs (infusion and hemorrhage), while $\Delta V_{B}$ is measured as dilution of hemoglobin ($K, \alpha, V_{0}$ are parameters of the model determined by parameter fitting). In our case, we know $u\left( t \right)$ – water added to blood from drinks, have no knowledge of $\Delta V_{B}$, while $v(t)$ – urine formation rate is the output of the model used for parameter fitting, and so it cannot be used in equation (s1) if we do not want to get trivial results (x=x). Instead, we need an equation that connects urine formation rate $v(t)$ with blood volume, osmolality, and concentrations of caffeine and alcohol in blood plasma. We also need equations to derive the above characteristics of blood from the bladder diary data, i.e., timing, volumes osmolality, caffeine, and alcohol content of the drinks. For this, we modeled dynamics of fluid transport and absorption in the stomach and intestine.

#### Structure and assumptions of the model

Our model consists of three compartments: stomach, intestine, and blood (connected to interstitial fluid through equations of [5]). We are modelling four variables in these three compartments: water volume (W), number of moles (E) of added electrolyte ions (osmolality of the drinks, which affects osmolality of the blood), number of mg (C) of caffeine, and of mL of alcohol (A).

Our model is based on the following main assumptions:

- Water, electrolytes, caffeine, alcohol, and food move through the stomach as a single bolus. It enters the duodenum with constant speed, limited by pylorus (gastric emptying). There is some limited absorption (first-order kinetics) of water, electrolytes, caffeine, and alcohol from the stomach to the blood, but most absorption is happening in the intestine [6].
- Water, electrolytes, caffeine, and alcohol entering the intestine from the stomach are absorbed to the blood (first-order kinetics). Concentrations of caffeine and alcohol are reduced by the liver prior to entering systemic circulation (first-pass metabolism) [7,8].
- Water and electrolytes are stable, while caffeine and alcohol are metabolized in systemic circulation. Caffeine is metabolized according to first-order kinetics, while alcohol is metabolized according to zero-order kinetics, limited by the amount of alcohol dehydrogenase [9].
- Electrolytes, caffeine, and alcohol are redistributed between blood plasma and interstitial fluid according to the same equation (s1) as water but with different values of parameters $\alpha$ and 𝐾.
- Urine formation rate $v(t)$ is a function of blood volume, osmolality, and concentrations of caffeine and alcohol. It may also depend on whether it is daytime or nighttime.

#### Equations of the model

##### Stomach

***Water***

$\frac{dW_{s}}{dt}=- k_{ge}\cdot\mathrm{erf} \left( \frac{W_{s}}{k_{ge}} \right)-k_{sw}\cdot W_{s}+intake(t)$	(s4)

Where *W_s_* – volume of water in the stomach, *k_ge_* – gastric emptying (mL/min) – volume of water that passes from the stomach to the intestine in 1 minute. We model gastric emptying (limited by a *k_ge_* constant volume per minute of drink/food exiting the stomach though pylorus) using nonlinear function: $\mathrm{erf} \left( x \right)=\int_{0}^{x} \exp\left( -t^{2} \right)dt$; $\mathrm{erf} \left( \frac{W_{s}}{k_{ge}} \right)$ = zero if *W_s_ = 0*, close to 1 if *W_s_> k_ge_ ,* $k_{sw}$ *–* coefficient of water adsorption in the stomach (1/min); $intake\left( t \right)= \frac{volume ofdrink}{duration of drink} , during the drink, zero otherwise.$

***Electrolytes***

Osmolality of the drinks varies from zero (distilled water) to 1000 (wine, beer) mmol/kg. Normal osmolality of blood plasma is 290 mmol/kg; it is the number of moles of electrolytes in 1 kg of fluid. Osmolarity is the number of moles of electrolytes in 1 liter of fluid. When considering the physiology of body fluids, the difference between osmolality and osmolarity is negligible because body fluids typically are dilute aqueous solutions [10]. We model how the amount of electrolyte moles added with the drink influence the amount of electrolytes in the stomach, intestine, and blood. We assume that electrolytes are moving through the stomach together with fluid due to gastric emptying and can be, to some extent, adsorbed in stomach. Source term = number of moles of electrolytes in the drink=*osmolarity(t) x intake(t).*

$\frac{dE_{s}}{dt}=$ $-k_{ge}\cdot\mathrm{erf} \left( \frac{W_{s}}{k_{ge}} \right)\cdot cE-k_{se}\cdot E_{s}+intake(t)\cdot cE$	(s5)

Where $E_{s}$ – number of moles of electrolytes (from the consumed drink) in the stomach, $cE$- *osmolarity(t)* of the consumed drink, $k_{se}$ – sorption coefficient for electrolytes in the stomach. Note that the first term in the right side of the equation is the volume of the drink moved (in 1 minute) from the stomach to the intestine, multiplied by the concentration of electrolytes in the drink.

***Caffeine***

Information on caffeine content for the drinks is typically available not as concentrations, but as number of mg per serving [11]. This requires attention for each caffeinated drink in the diary. For the sake of the symmetry of equations, let us write the source term as *intake(t) x (caffeine per serving)/(volume of the serving) = intake(t) x cC.*

We assume that caffeine is transported through the stomach together with water due to gastric emptying, partially adsorbed in the stomach but mostly in the intestine. Unlike water and electrolytes, which are stable, caffeine is metabolized (first-order kinetics) [12], which is assumed happening mostly in the blood versus the stomach and intestine.

$\frac{dC_{s}}{dt}=$ $- k_{ge}\cdot\mathrm{erf} \left( \frac{W_{s}}{k_{ge}} \right)\cdot cC-k_{sc}\cdot C_{s}+intake(t)\cdot cC$	(s6)

Where $C_{s}$ – number of mg of caffeine in the stomach, $cC$ – concentration of caffeine in the drink, $k_{sc}$ – caffeine in stomach sorption coefficient. Note that the first term in the right side of the equation is the volume of the drink moved (in 1 minute) from the stomach to the intestine, multiplied by the concentration of caffeine in the drink.

***Alcohol***

We assume that alcohol is transported through the stomach together with water due to gastric emptying, and is partially adsorbed in the stomach but mostly in the intestine. Unlike water and electrolytes, which are stable, alcohol is metabolized (zero-order kinetics) [13], which is assumed happening mostly in the blood versus the stomach and intestine.

$\frac{dA_{s}}{dt}=$ $- k_{ge}\cdot\mathrm{erf} \left( \frac{W_{s}}{k_{ge}} \right)\cdot cA-k_{sa}\cdot A_{s}+intake(t)\cdot cA$	(s7)

Where $A_{s}$ – number of mL of alcohol in the stomach, $cA$ – concentration of alcohol in the drink (usually provided in %, so should be divided by 100), $k_{sa}$ – alcohol in stomach sorption coefficient. Note that the first term in the right side of the equation is the volume of the drink moved (in 1 minute) from the stomach to the intestine, multiplied by the concentration of alcohol in the drink.

##### Intestine

***Water***

$\frac{dW_{i}}{dt}=k_{ge}\cdot\mathrm{erf} \left( \frac{W_{s}}{k_{ge}} \right)-k_{iw}\cdot W_{i}$	(s8)

Where *W_i_* – volume of water from the drink in the intestine. The first term in the right side of the equation is fluid transported from the stomach due to gastric emptying, $k_{iw}$ – coefficient of absorption of water in the intestine, which should be higher than in the stomach.

***Electrolytes***

$\frac{dE_{i}}{dt}=$ $k_{ge}\cdot\mathrm{erf} \left( \frac{W_{s}}{k_{ge}} \right)\cdot cE-k_{ie}\cdot E_{i}$	(s9)

Where $E_{i}$ – number of moles of electrolytes (from the consumed drink) in the intestine, $cE$- *osmolality(t)* of the consumed drink, $k_{si}$ – electrolytes in intestine absorption coefficient. Note that the source term in the right side of the equation is the volume of the drink moved (in 1 minute) from the stomach to the intestine, multiplied by the concentration of electrolytes in the drink.

***Caffeine***

$\frac{dC_{i}}{dt}=$ $k_{ge}\cdot\mathrm{erf} \left( \frac{W_{s}}{k_{ge}} \right)\cdot cC-k_{ic}\cdot C_{i}$	(s10)

Where $C_{i}$ – number of mg of caffeine in the intestine, $cC$ – concentration of caffeine in the drink, $k_{ic}$ – caffeine in intestine absorption coefficient. Note that the source term in the right side of the equation is the volume of the drink moved (in 1 minute) from the stomach to the intestine, multiplied by the concentration of caffeine in the drink.

***Alcohol***

$\frac{dA_{i}}{dt}=$ $k_{ge}\cdot\mathrm{erf} \left( \frac{W_{s}}{k_{ge}} \right)\cdot cA-k_{ia}\cdot A_{i}$	(s11)

Where $A_{i}$ – number of mL of alcohol in the stomach, $cA$ – concentration of alcohol in the drink (usually provided in %, so should be divided by 100), $k_{ia}$ – alcohol in intestine sorption coefficient. Note that the source term in the right side of the equation is the volume of the drink moved (in 1 minute) from the stomach to the intestine, multiplied by the concentration of alcohol in the drink.

##### Blood-interstitial fluid compartment

***Water***

Following [5], we substituted eq. (s2) into eq. (s1), which resulted in the second-order ordinary differential equation (ODE) for the volume ($\Delta V_{B}$) of water added to the blood plasma due to the intake:

$\frac{d^{2}\Delta V_{B}}{{dt}^{2}}+K\cdot\frac{d\Delta V_{B}}{dt}=\frac{du}{dt}-\frac{dv}{dt}+\frac{K\cdot\left[ u\left( t \right)-v\left( t \right) \right]}{\left( 1+\alpha\right)}$	(s12),

where $u\left( t \right)$ is rate at which water is absorbed to blood from the stomach and intestine:

$u\left( t \right)=k_{sw}\cdot W_{s}$+$k_{iw}\cdot W_{i}$	(s13)

while $v\left( t \right)$ is the rate at which water volume in blood is reduced due to urine formation.

***Urine formation rate***

We assumed the following functional dependences for urine formation rate:

$v\left( t \right)=A_{0}\cdot(V_{B0}+{\Delta V}_{B})\cdot F_{c}\cdot F_{a}\cdot F_{n}\cdot F_{p}\cdot F_{e}$	(s14),

where$V_{B0}$ is the volume of blood plasma at equilibrium; $A_{0}\cdot V_{B0}$ – glomerular filtration rate (GFR) at equilibrium (mL/min). $F_{c}$ – describes change in GFR due to the presence of caffeine in the blood. $F_{a}$ – describes change of GFR due to the presence of alcohol in the blood. $F_{n}$ – describes the change of GFR at night. $F_{p}$ – describes feedback loop for blood pressure regulation in response to increased blood volume. $F_{e}$ – describes reabsorption of water in renal tubules (about 0.8% is not reabsorbed in normal conditions when blood osmolarity = 290mmol/L = 0.29Mol/L, more is reabsorbed if osmolarity is higher).

We model $F_{e}$ as: $F_{e}=1-\mathrm{erf} \left[ 1.87+k_{osm}\cdot\left( CE-0.29 \right) \right] if CE>0.29$(s15)

$F_{e}=1-\mathrm{erf} \left( 1.87 \right) if CE<0.29$

$CE=\frac{E_{B}}{(V_{B0}+{\Delta V}_{B})}$ – osmolarity of blood, where $E_{B}$– number of moles of electrolytes in blood. Note that, if $\mathrm{CE}\leq$0.29, $F_{e}=1-\mathrm{erf} \left( 1.87 \right)=$0.008; $k_{osm}$ – coefficient determining the sensitivity of blood osmolarity regulation. Note that the $k_{osm}\cdot\left( CE-0.29 \right)$ term describes reabsorption of water due to the action of vasopressin, which forces aquaporin molecules (water channels) to move to the surface of the cells in renal tubules.

We model $F_{p}$ as: $F_{p}=1-\beta_{1}\cdot erf(\frac{{\Delta V}_{B}}{V_{B0}})$	(s16)

Note that $\beta_{1}$ is the maximum possible reduction in blood flow due to blood pressure regulation.

We model $F_{n}$ as: $F_{n}=1-k_{n}\cdot\sum_{n=1}^{N} (erf \left( \left( t-t_{bn} \right)/t_{s} \right)-erf(\left( t-t_{wn} \right)/t_{s}))/2$	(s17),

where N – number of nights covered by the voiding diary; $t_{bn}, t_{wn}$ are “going to bed” and waking times, and $t_{s}$ – time required to fall asleep and wake up.

We model $F_{c}$as: $F_{c}=1+$ $A_{caf}$ $\cdot$ $CC, if$ $CC<0.1;$ $F_{c}=1+$ $A_{caf}$ $\cdot$ 0.1, if $CC\geq0.1$ (s18),

where $CC=\frac{C_{B}}{(V_{B0}+{\Delta V}_{B})}$ – concentration of alcohol in blood, $C_{B}$ – number of mg of caffeine in blood.

We model $F_{a}$as:

$F_{a}=1+$ $A_{alc}$ $\cdot$ $CA, if$ $CA<0.002;$	$F_{a}=1+$ $A_{alc}$ $\cdot$ 0.002, if	$CA\geq0.002;$	(s19),

where $CA=\frac{A_{B}}{(V_{B0}+{\Delta V}_{B})}$ – concentration of alcohol in blood, $A_{B}$ – number of mL of alcohol in blood.

Eq. s12 includes term $\frac{dv}{dt}$ , which according to (s14) is equal to:

$\frac{dv}{dt}=A_{0}\cdot\left\{ \frac{d\Delta V_{B}}{dt}\cdot F_{c}\cdot F_{a}\cdot F_{n}\cdot F_{p}\cdot F_{e}+(V_{B0}+{\Delta V}_{B})\cdot\left[ \frac{dF_{c}}{dt}\cdot F_{a}\cdot F_{n}\cdot F_{p}\cdot F_{e}+F_{c}\cdot\frac{dF_{a}}{dt}\cdot F_{n}\cdot F_{p}\cdot F_{e}+F_{c}\cdot F_{a}\cdot\frac{dF_{n}}{dt}\cdot F_{p}\cdot F_{e}+F_{c}\cdot F_{a}\cdot F_{n}\cdot\frac{dF_{p}}{dt}\cdot F_{e}+F_{c}\cdot F_{a}\cdot F_{n}\cdot F_{p}\cdot\frac{dF_{e}}{dt} \right] \right\}$	(s20)

By using $F=F_{c}\cdot F_{a}\cdot F_{n}\cdot F_{p}\cdot F_{e}$ , eqs. s14 and s20 can be simplified to:

$v\left( t \right)=A_{0}\cdot F\cdot(V_{B0}+{\Delta V}_{B})$	(s21)

and $\frac{dv}{dt}=A_{0}\cdot F\cdot\left\{ \frac{d\Delta V_{B}}{dt}+(V_{B0}+{\Delta V}_{B})\cdot\left[ \frac{\frac{dF_{c}}{dt}}{F_{c}}+\frac{\frac{dF_{a}}{dt}}{F_{a}}+\frac{\frac{dF_{n}}{dt}}{F_{n}}+\frac{\frac{dF_{p}}{dt}}{F_{p}}+\frac{\frac{dF_{e}}{dt}}{F_{e}} \right] \right\}$	(s22)

Substituting eqs. (s13), (s21), and (s22) into eq. (s12) results in the second-order ODE for ${\Delta V}_{B}$:

$\frac{d^{2}\Delta V_{B}}{{dt}^{2}}=-\left( K+A_{0}\cdot F \right)\cdot$ $\frac{d\Delta V_{B}}{dt}$

$$-A_{0}\cdot F\cdot\left( \frac{K}{1+\alpha}+\frac{\frac{dF_{c}}{dt}}{F_{c}}+\frac{\frac{dF_{a}}{dt}}{F_{a}}+\frac{\frac{dF_{n}}{dt}}{F_{n}}+\frac{\frac{dF_{p}}{dt}}{F_{p}}+\frac{\frac{dF_{e}}{dt}}{F_{e}} \right)\cdot{\Delta V}_{B}$$

$$-A_{0}\cdot F\cdot V_{B0}\cdot\left( \frac{K}{1+\alpha}+\frac{\frac{dF_{c}}{dt}}{F_{c}}+\frac{\frac{dF_{a}}{dt}}{F_{a}}+\frac{\frac{dF_{n}}{dt}}{F_{n}}+\frac{\frac{dF_{p}}{dt}}{F_{p}}+\frac{\frac{dF_{e}}{dt}}{F_{e}} \right)$$

$+k_{sw}\cdot\frac{dW_{s}}{dt}+ k_{iw}\cdot\frac{dW_{i}}{dt}+ \frac{K}{1+\alpha}\cdot\left( k_{sw}\cdot W_{s}+k_{iw}\cdot W_{i} \right)$	(s23)

***Electrolytes***

Electrolytes are dissolved in the water of the drinks and in blood plasma. We are assuming that electrolytes added with drinks are redistributed between blood and interstitial fluid in the same way as water (but with different parameters *K_e_* and *α_e_* values):

$\frac{d\Delta E_{B}}{dt}=u_{e}\left( t \right)-v_{e}\left( t \right)-q_{e}(t)$	(s24)

where $\Delta E_{B}$ – change of the number of moles of electrolytes in blood plasma; $u_{e}\left( t \right)$ – number of moles of electrolytes adsorbed to blood from drinks in the stomach and intestine, $v_{e}\left( t \right)$ number of moles of electrolytes removed with urine, $q_{e}(t)$ – describes redistribution of electrolytes between blood and interstitial fluid. As noted above, reabsorption of electrolytes is different from reabsorption of water; the $k_{osm}\cdot\left( CE-0.29 \right)$ term in the equation (s15) for $v\left( t \right)$describes reabsorption of water due to the action of vasopressin, which forces aquaporin molecules (water channels) to move to the surface of the cells in renal tubules. This process does not facilitate reabsorption of electrolytes, caffeine, and alcohol. For electrolytes, $F_{e}$ in the equation for $v\left( t \right)$should be substituted by $F_{0e}$=$1-erf(1.87-k_{anp}\cdot(CE-0.29))$, which describes the reduced reabsorption of sodium in response to the increased osmolality of blood plasma. This process is facilitated by the ANP hormone synthesized when vasopressin concentration is increased in response to increased osmolality. This allows faster excretion of sodium from blood than just by increased water reabsorption. It is equivalent to substituting $F$ with $F_{1e}=F_{c}\cdot F_{a}\cdot F_{n}\cdot F_{p}\cdot F_{0e}$ . Note that this process is not involved in excretion of alcohol or caffeine, so for these, $F_{0}$=$1-erf(1.87)$, should be used instead of $F_{0e}$.

$u_{e}\left( t \right)$ =$k_{se}\cdot E_{s}+$ $k_{ie}\cdot E_{i}$	(s25)

$v_{e}\left( t \right)$ =$v\left( t \right)\cdot\frac{F_{0e}}{F_{e}}\cdot CE=$ $A_{0}\cdot F_{1e}(V_{B0}+{\Delta V}_{B})\cdot$ $\frac{E_{B}}{(V_{B0}+{\Delta V}_{B})}=$ $A_{0}\cdot F_{1e}\cdot$ $E_{B}=$ $A_{0}\cdot F_{1e}\cdot\left( E_{B0}+\Delta E_{B} \right)$	(s26)

$q_{e}\left( t \right)=K_{e}\left\{ \Delta E_{B}-\frac{1}{1+\alpha_{e}}\cdot\int_{0}^{t} \left[ u_{e}\left( \tau\right)-v_{e}(\tau) \right]d\tau\right\}$	(s27)

Leading to:

$\frac{d^{2}\Delta E_{B}}{{dt}^{2}}+K_{e}\cdot\frac{d{\Delta E}_{B}}{dt}=\frac{du_{e}}{dt}-\frac{dv_{e}}{dt}+\frac{K_{e}\cdot\left[ u_{e}\left( t \right)-v_{e}(t) \right]}{(1+\alpha_{e})}$	(s28),

where

$\frac{dv_{e}}{dt}=A_{0}\cdot F_{1e}\cdot\frac{d\Delta E_{B}}{dt}+$ $A_{0}\cdot\frac{dF_{1e}}{dt}\cdot\left( E_{B0}+\Delta E_{B} \right)$

$=$ $A_{0}\cdot F_{1e}\cdot\left\{ \frac{d\Delta E_{B}}{dt}+(E_{B0}+{\Delta E}_{B})\cdot\left[ \frac{\frac{dF_{c}}{dt}}{F_{c}}+\frac{\frac{dF_{a}}{dt}}{F_{a}}+\frac{\frac{dF_{n}}{dt}}{F_{n}}+\frac{\frac{dF_{p}}{dt}}{F_{p}}+\frac{\frac{dF_{0e}}{dt}}{F_{0e}} \right] \right\}$	(s29)

By substituting (s25), (s26), and (s29) into (s26), we get:

$\frac{d^{2}\Delta E_{B}}{{dt}^{2}}+K_{e}\cdot\frac{d{\Delta E}_{B}}{dt}=k_{se}\cdot\frac{dE_{s}}{dt}+ k_{ie}\cdot\frac{dE_{i}}{dt}$ +$\frac{K_{e}}{1+\alpha_{e}}\cdot\left( k_{se}\cdot E_{s}+k_{ie}\cdot E_{i} \right)$ $-\frac{K_{e}}{1+\alpha_{e}}\cdot A_{0}\cdot F_{1e}\cdot(E_{B0}+{\Delta E}_{B})$ - $A_{0}\cdot F_{1e}\cdot\left\{ \frac{d\Delta E_{B}}{dt}+(E_{B0}+{\Delta E}_{B})\cdot\left[ \frac{\frac{dF_{c}}{dt}}{F_{c}}+\frac{\frac{dF_{a}}{dt}}{F_{a}}+\frac{\frac{dF_{n}}{dt}}{F_{n}}+\frac{\frac{dF_{p}}{dt}}{F_{p}}+\frac{\frac{dF_{0e}}{dt}}{F_{0e}} \right] \right\}$	(s30),

and finally:

$\frac{d^{2}\Delta E_{B}}{{dt}^{2}}=-\left( K_{e}+A_{0}\cdot F_{1e} \right)\cdot$ $\frac{d\Delta E_{B}}{dt}$

$$-A_{0}\cdot F_{1e}\cdot\left( \frac{K_{e}}{1+\alpha_{e}}+\frac{\frac{dF_{c}}{dt}}{F_{c}}+\frac{\frac{dF_{a}}{dt}}{F_{a}}+\frac{\frac{dF_{n}}{dt}}{F_{n}}+\frac{\frac{dF_{p}}{dt}}{F_{p}}+\frac{\frac{dF_{0e}}{dt}}{F_{0e}} \right)\cdot{\Delta E}_{B}$$

$$-A_{0}\cdot F_{1e}\cdot E_{B0}\cdot\left( \frac{K_{e}}{1+\alpha_{e}}+\frac{\frac{dF_{c}}{dt}}{F_{c}}+\frac{\frac{dF_{a}}{dt}}{F_{a}}+\frac{\frac{dF_{n}}{dt}}{F_{n}}+\frac{\frac{dF_{p}}{dt}}{F_{p}}+\frac{\frac{dF_{0e}}{dt}}{F_{0e}} \right)$$

$+k_{se}\cdot\frac{dE_{s}}{dt}+ k_{ie}\cdot\frac{dE_{i}}{dt}+ \frac{K_{e}}{1+\alpha_{e}}\cdot\left( k_{se}\cdot E_{s}+k_{ie}\cdot E_{i} \right)$	(s31)

***Caffeine and alcohol***

We assume that caffeine and alcohol from drinks are redistributed between blood and interstitial fluid the same way as water but with different parameter values ($K_{c},\alpha_{c}, K_{A},\alpha_{A}, ).$ There are three additional differences in modeling caffeine and alcohol kinetics: (a) we assume that initial concentrations of caffeine and alcohol in blood are zero, so the third term in the right side of equation analogous to (s31) will disappear; (b) unlike water and electrolytes, caffeine and alcohol are not stable, but are metabolized by the body; (c) the amount of caffeine and alcohol in the blood will be reduced by the liver prior to entering systemic circulation (first-pass metabolism). So, additional terms describing reduction of the concentration of caffeine and alcohol should be added to the equations analogous to eq. (s12).

$\frac{d\Delta C_{B}}{dt}=u_{c}\left( t \right)-v_{c}\left( t \right)-q_{c}\left( t \right)-d_{c}\cdot\Delta C_{B}$	(s32)

which results in:

$\frac{d^{2}\Delta C_{B}}{{dt}^{2}}=- \left( K_{c}+A_{0}\cdot F_{1}+d_{c} \right)\cdot$ $\frac{d\Delta C_{B}}{dt}$

$$-A_{0}\cdot F_{1}\cdot\left( \frac{K_{c}}{1+\alpha_{c}}+\frac{\frac{dF_{c}}{dt}}{F_{c}}+\frac{\frac{dF_{a}}{dt}}{F_{a}}+\frac{\frac{dF_{n}}{dt}}{F_{n}}+\frac{\frac{dF_{p}}{dt}}{F_{p}} \right)\cdot{\Delta C}_{B}$$

$+(k_{sc}\cdot\frac{dC_{s}}{dt}+ k_{ic}\cdot\frac{dC_{i}}{dt}+ \frac{K_{c}}{1+\alpha_{c}}\cdot\left( k_{sc}\cdot C_{s}+k_{ic}\cdot C_{i} \right))$ $\cdot{fpm}_{c}$	(s33),

where ${\Delta C}_{B}$ is the change of the number of mg of caffeine in blood and $d_{c}$(min^-1^) is the coefficient of dissociation of caffeine by enzymes, ${fpm}_{c}$ – reduction of caffeine in systemic circulation due to first-pass metabolism, and $F_{1}=F_{c}\cdot F_{a}\cdot F_{n}\cdot F_{p}\cdot F_{0}$ .

For alcohol, dissociation is different since it is limited by the amount of alcohol dehydrogenase enzyme and, unlike the case with caffeine, cannot be described by first-order kinetics. We will describe dissociation of alcohol by the term ${- d}_{a}\cdot erf(\Delta A_{B}/d_{a})$ , which is zero when $\Delta A_{B}=0$ and ≈ ${- d}_{a}$when $\Delta A_{B}>d_{a}$, which results in:

$\frac{d^{2}\Delta A_{B}}{{dt}^{2}}=- \left( K_{A}+A_{0}\cdot F_{1}+exp(-\left( \Delta A_{B}/d_{a} \right)^{2}) \right)\cdot$ $\frac{d\Delta A_{B}}{dt}$

$$-A_{0}\cdot F_{1}\cdot\left( \frac{K_{A}}{1+\alpha_{A}}+\frac{\frac{dF_{c}}{dt}}{F_{c}}+\frac{\frac{dF_{a}}{dt}}{F_{a}}+\frac{\frac{dF_{n}}{dt}}{F_{n}}+\frac{\frac{dF_{p}}{dt}}{F_{p}} \right)\cdot{\Delta A}_{B}$$

$+ (k_{sa}\cdot\frac{dA_{s}}{dt}+ k_{ia}\cdot\frac{dA_{i}}{dt}+ \frac{K_{A}}{1+\alpha_{A}}\cdot\left( k_{sa}\cdot A_{s}+k_{ia}\cdot A_{i} \right))\cdot$ ${fpm}_{A}$	(s34),

where ${\Delta A}_{B}$ is the change in the number of mL of alcohol in blood and ${fpm}_{A}$ – reduction of alcohol in systemic circulation due to first-pass metabolism.

#### Model implementation

The model was implemented with System Identification Toolbox (MATLAB 2021a), using function idnlgrey.m , as a nonlinear grey-box model, which requires its representation as a state-space model – set of the first-order ODEs. The states (variables of the model) are the following: x(1)= $\Delta V_{B}$ – change of the blood volume, x(2) =$\frac{d\Delta V_{B}}{dt};$x(3) =$\Delta E_{B}$ – change of the number of moles of electrolytes in blood, x(4) =$\frac{d\Delta E_{B}}{dt}$; x(5) =$\Delta C_{B}$ – change of the number of mg of caffeine in blood, x(6) =$\frac{d\Delta C_{B}}{dt}$; x(7) = $\Delta A_{B}$ – change of the number of mL of alcohol in blood, x(8) =$\frac{d\Delta A_{B}}{dt}$; x(9) =$W_{s}$ – volume of water from drinks in the stomach (mL), x(10) =$E_{s}$ – number of moles of electrolytes from drinks in the stomach (mol), x(11) =$C_{s}$ – caffeine from drinks in the stomach (mg), x(12) =$A_{s}$ – number of mL of alcohol from drinks in the stomach (mL), x(13) =$W_{i}$ – volume of water from drinks in the intestine (mL), x(14) =$E_{i}$ – number of moles of electrolytes from drinks in the intestine (mol), x(15) =$C_{s}$ – caffeine from drinks in the intestine (mg), x(16) =$A_{i}$ – alcohol from drinks in the intestine (mL). The model consists of 16 ODEs. The set of ODEs include eight first-order eqs. (s4-s11) describing dynamics of added water, electrolytes, caffeine, and alcohol in the stomach and intestine, together with eight more first-order ODEs derived from second-order ODEs (s23), (s31), (s33), (s34) describing redistribution of water, electrolytes, caffeine, and alcohol between blood and interstitial fluid. The total of 30 parameters of these 16 equations were reduced to 22 free parameters by assuming $k_{sw}= k_{se}=k_{sc}=0.5\cdot k_{sa}$, and $k_{iw}= k_{ie}= k_{ic}=k_{ia}$, $t_{s}$ = 20 minutes, and estimating blood plasma volume from participants’ weight, height, and sex, by using the Nadler equation [14]. The values of free parameters were determined by the idnlgrey.m function that used the reflective Newton optimization algorithm to minimize the difference between the urine formation rate profile $v\left( t \right)$ predicted by the model and the bladder-filling rate profile determined from the bladder diaries (defined as FitPercent eq. 3 in the main text of the paper). In order to reduce the likelihood of finding local instead of global minima, the initial values of free parameters were randomly selected within the allowed physiologically reasonable intervals. Forty instances of idnlgrey.m with randomly selected initial values of the parameters were run in parallel on a 40-core processor using the spmd.m function; then the model with the best fit was selected.

As described in the main text, models for some (n=52) individuals did not pass our selection criterion of Peak Fit >0.9. For others (n=145), “satisfactory” models of urine production rate were selected for use in further analysis and simulation. Supplemental Figure S11 presents the box plots for parameters of the selected 145 models. To enable box plot comparison, parameter values are divided by their mean values calculated across 145 models. The highest variability is demonstrated by parameter $k_{n}$, introduced (eq. s17) to describe changes in GFR at nighttime versus daytime. Interestingly, although the allowed parameter range was $-1{<k}_{n}<1$, about the same number of models predicted increase and decrease of GFR during nighttime, leading to nearly zero mean parameter value and high parameter variability relative to its mean value. High variability is also demonstrated by parameters describing redistribution and metabolism of alcohol$K_{A}$,$\alpha_{A}$, and $d_{a}$ , which is in agreement with literature data [15] on the high variability of alcohol metabolism across individuals. Also, as seen in Supplemental Figure S11, variability in the coefficients $K_{W},K_{C}, K_{A}$describing the speed at which equilibrium distribution of water, caffeine, and alcohol between blood and interstitial fluid is established is much higher than variability in parameters $\alpha_{W}$,$\alpha_{C}$,$\alpha_{A}$ describing the level of the equilibrium.

### Supplemental References

1. Moss R, Thomas SR. Hormonal regulation of salt and water excretion: a mathematical model of whole kidney function and pressure natriuresis. *Am J Physiol Renal Physiol* 2014;306(2):F224-F248.
2. Karaaslan F, Denizhan Y, Kayserilioglu A, Gulcur HO. Long-term mathematical model involving renal sympathetic nerve activity, arterial pressure, and sodium excretion. *Ann Biomed Eng* 2005;33(11):1607-1630.
3. Adolph EF. Physiological regulations. Lancaster, PA: The Jacques Cattell Press, 1943.
4. Bossingham MJ, Carnell NS, Campbell WW. Water balance, hydration status, and fat-free mass hydration in younger and older adults. *Am J Clin Nutr* 2005;81(6):1342-1350.
5. Bighamian R, Reisner AT, Hahn JO. A lumped-parameter subject-specific model of blood volume response to fluid infusion. *Front Physiol* 2016;7:390.
6. Leiper JB. Fate of ingested fluids: factors affecting gastric emptying and intestinal absorption of beverages in humans. *Nutr Rev* 2015;73(Suppl 2):57-72.
7. Sharma R, Gentry RT, Lim RT, Lieber CS. First-pass metabolism of alcohol. *Digest Dis Sci* 1995;40: 2091-2097.
8. Alsabri SG, Mari WO, Younes S, Alsadawi MA, Oroszi TL. Kinetic and dynamic description of caffeine. *J Caffeine Adenosine Res* 2018;8(1):3-9; Erratum: *J Caffeine Adenosine Res* 2021;11(4):107.
9. Krebs HA, Perkins JR. The physiological role of liver alcohol dehydrogenase. *Biochem J* 1970;118(4):635-644.
10. DiBartola SP. Disorders of sodium and water. In: Fluid, electrolyte, and acid-base disorders in small animal practice (4th ed). Amsterdam: Elsevier; 2012. Chapter 3.
11. McCusker RR, Goldberger BA, Cone EJ. Caffeine content of energy drinks, carbonated sodas, and other beverages. *J Anal Toxicol* 2006;30(2):112-114.
12. Nehlig A. Interindividual differences in caffeine metabolism and factors driving caffeine consumption. *Pharmacol Rev* 2018;70:384-411.
13. Zakhari S. Overview: How is alcohol metabolized by the body? *Alcohol Res Health* 2006;29(4):245-254.
14. Nadler SB, Hidalgo JH, Bloch T. Prediction of blood volume in normal human adults. *Surgery* 1962;51(2):224-232.
15. Collins AC, Yeager TN, Lebsack ME, Panter SS. Variations in alcohol metabolism: Influence of sex and age. *Pharmacol Biochem Behav* 1975;3(6):973-978.

**Supplemental Figures**

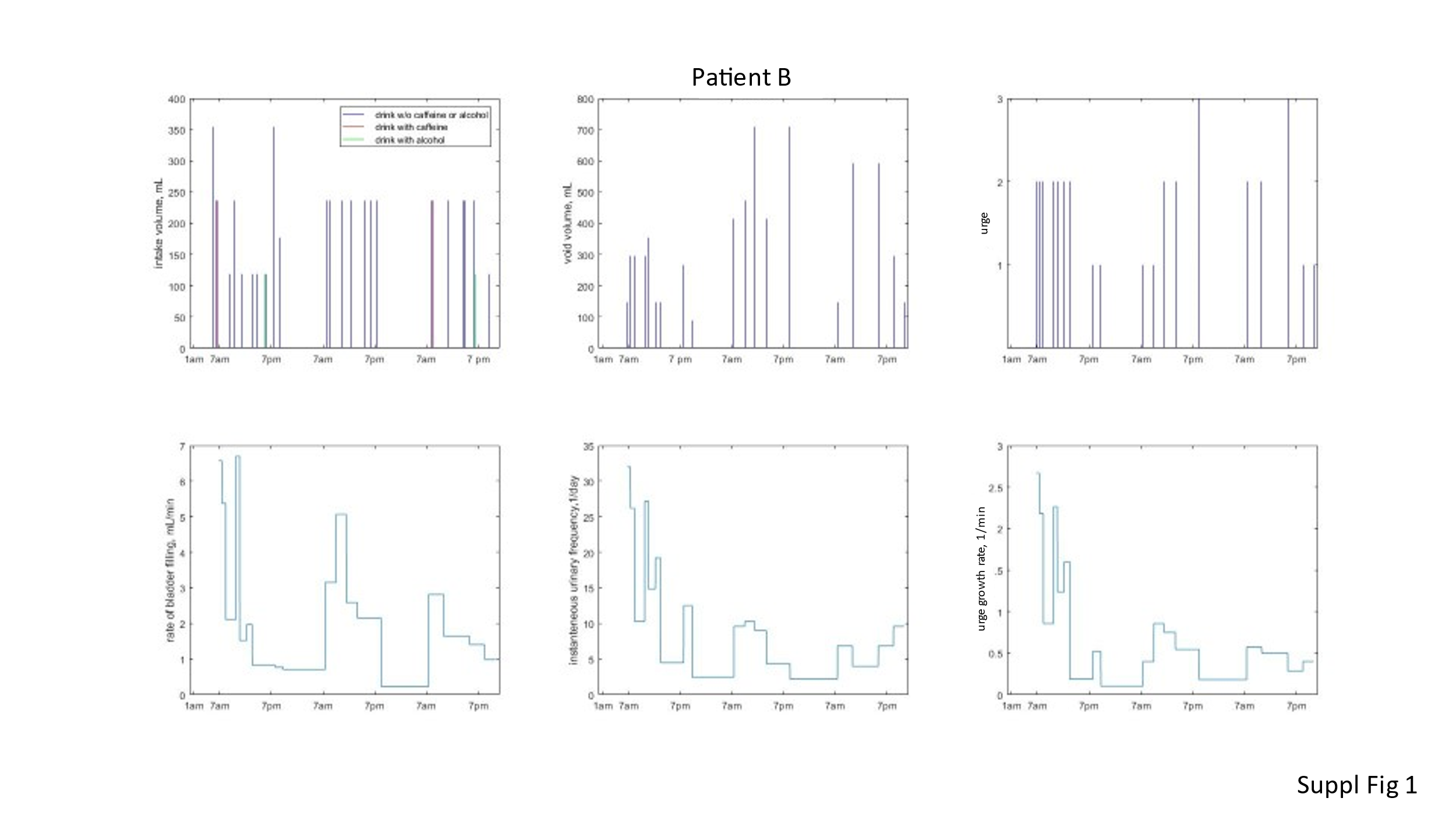

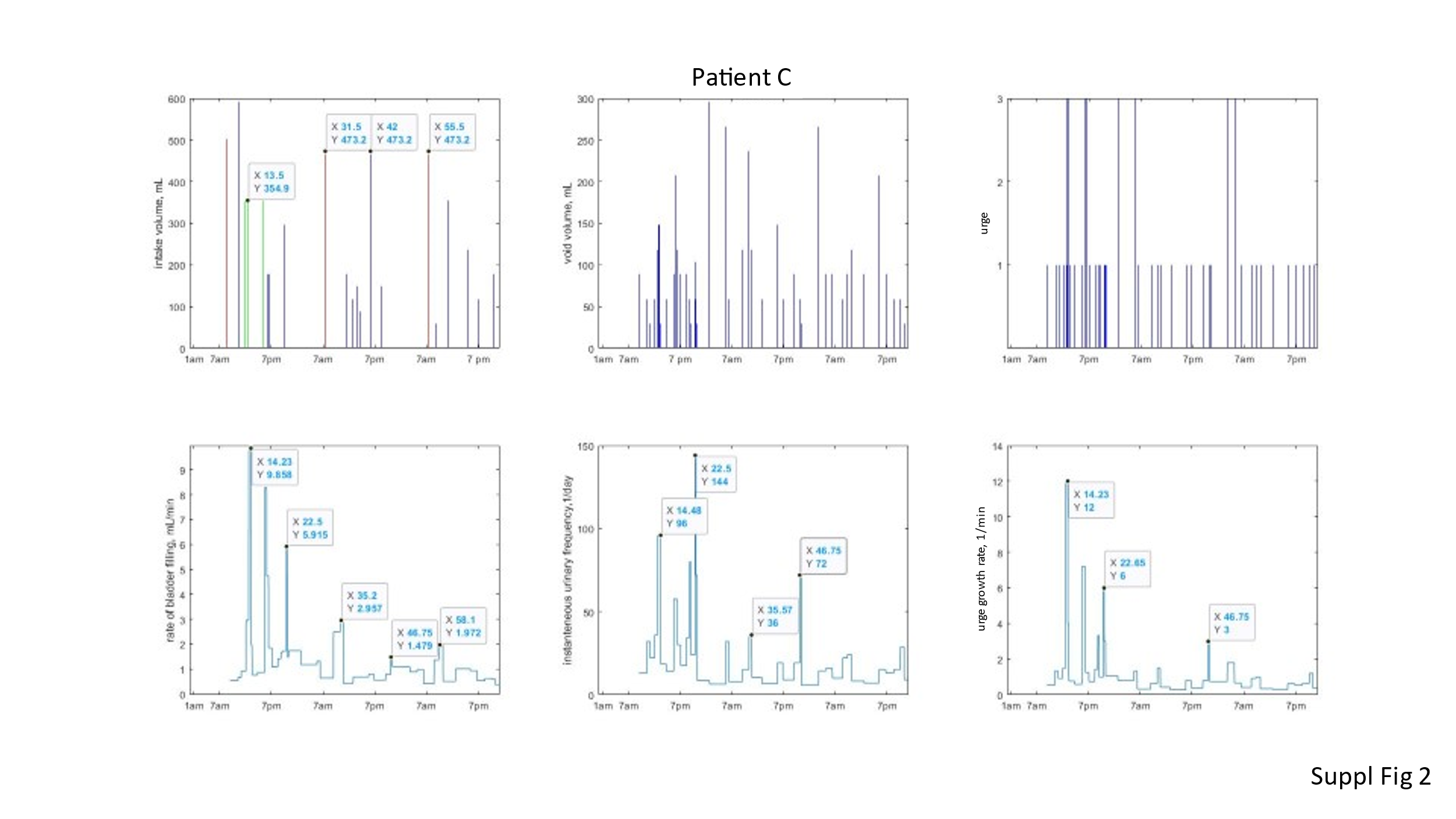

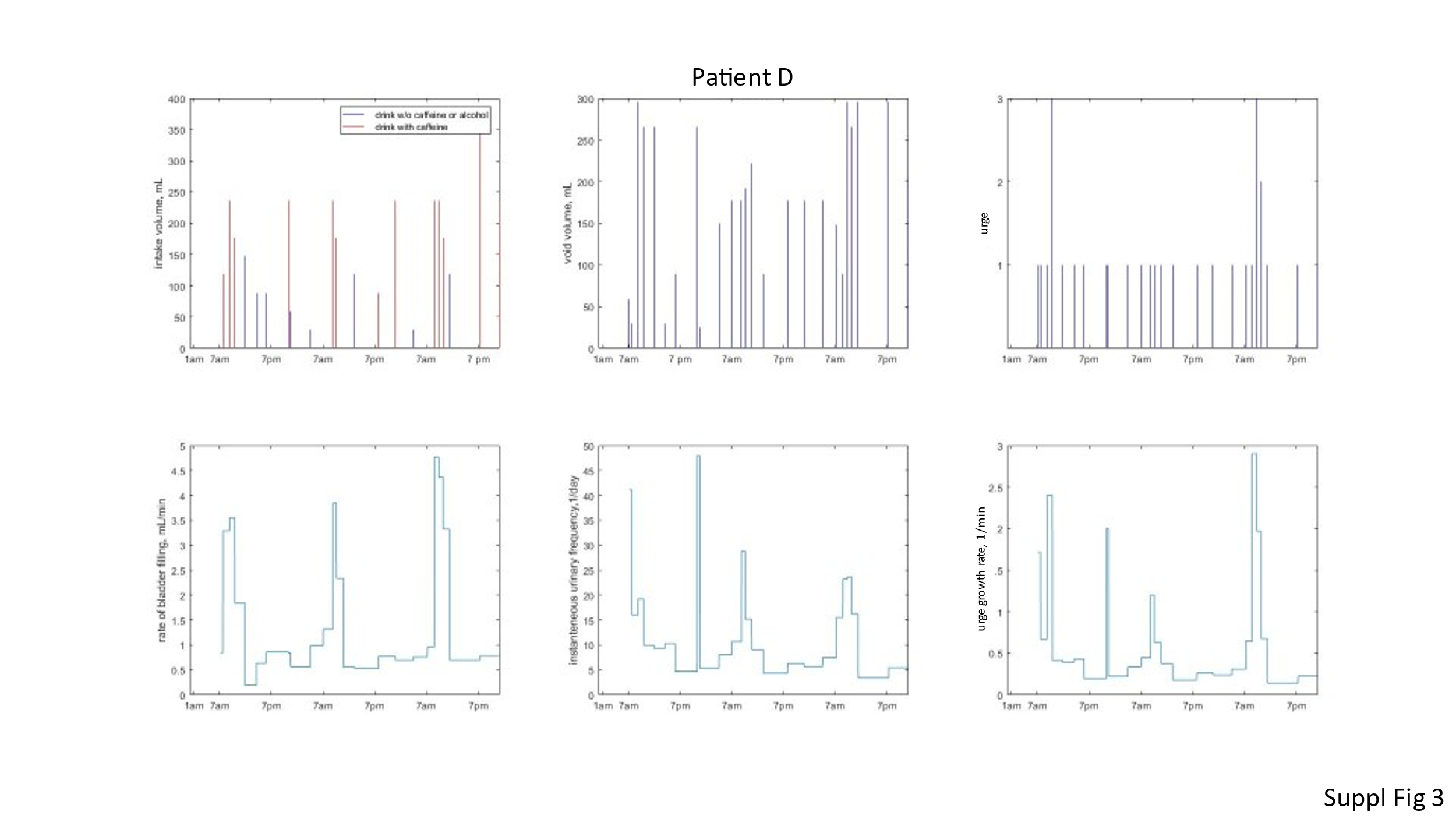

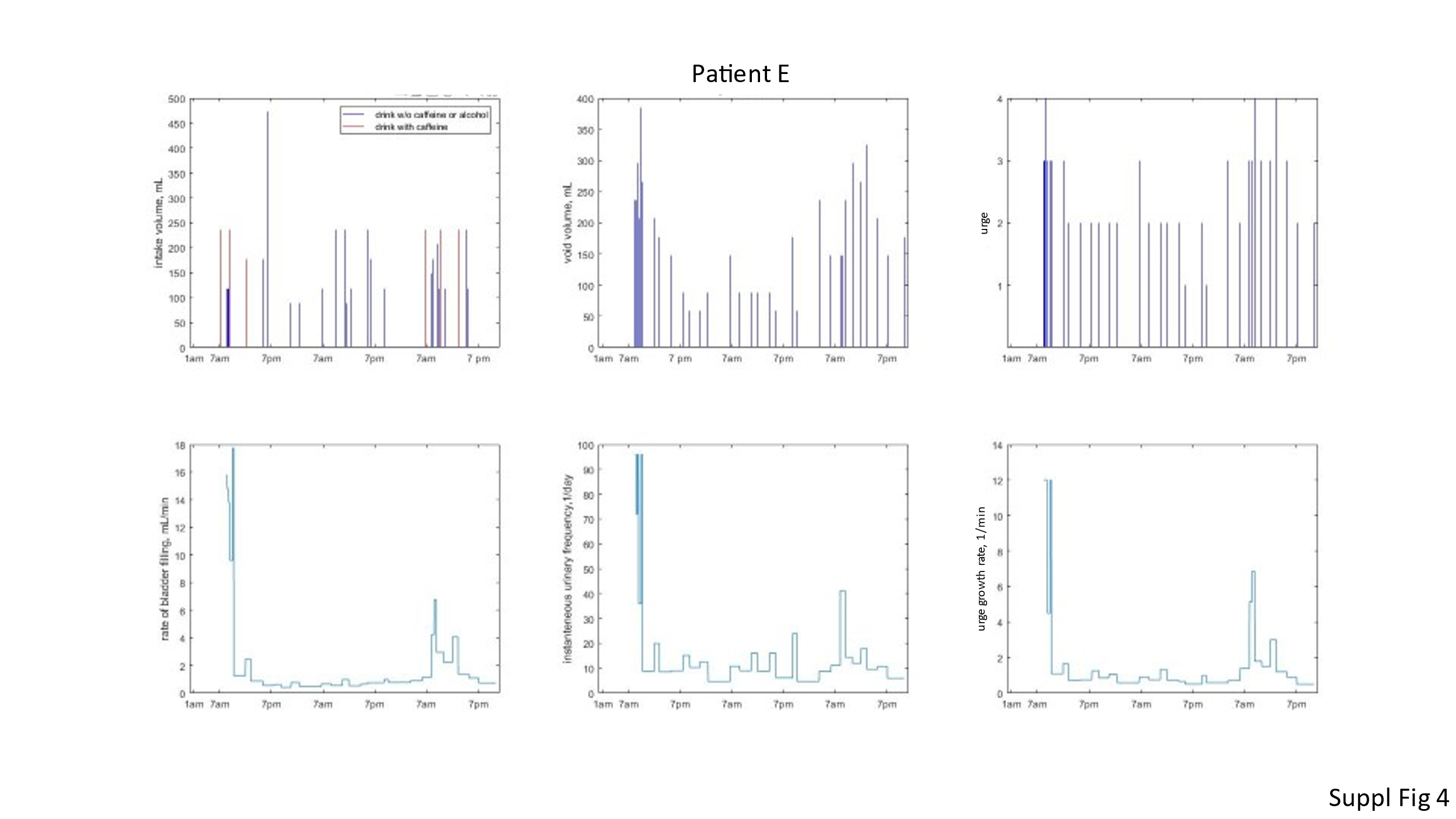

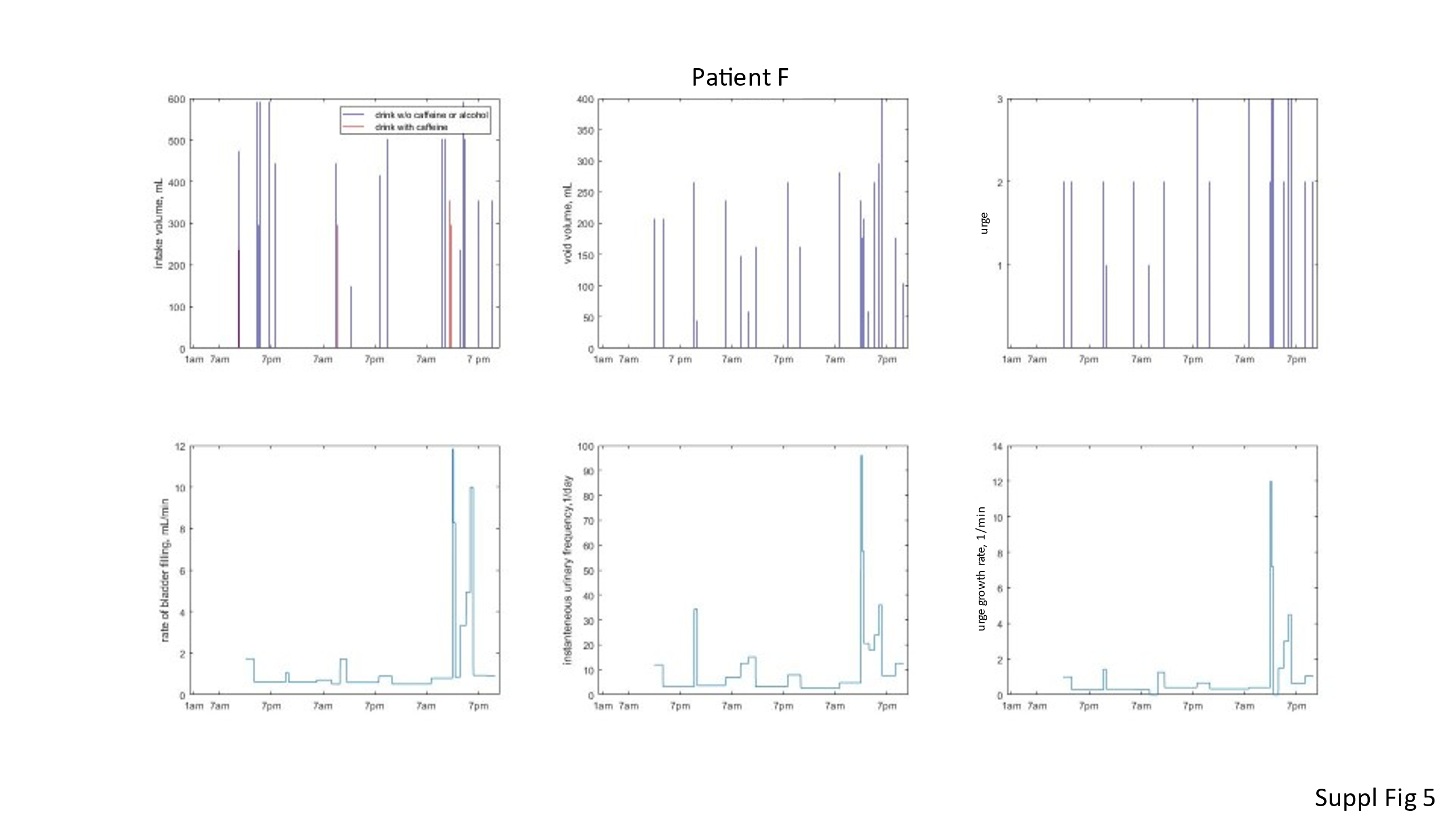

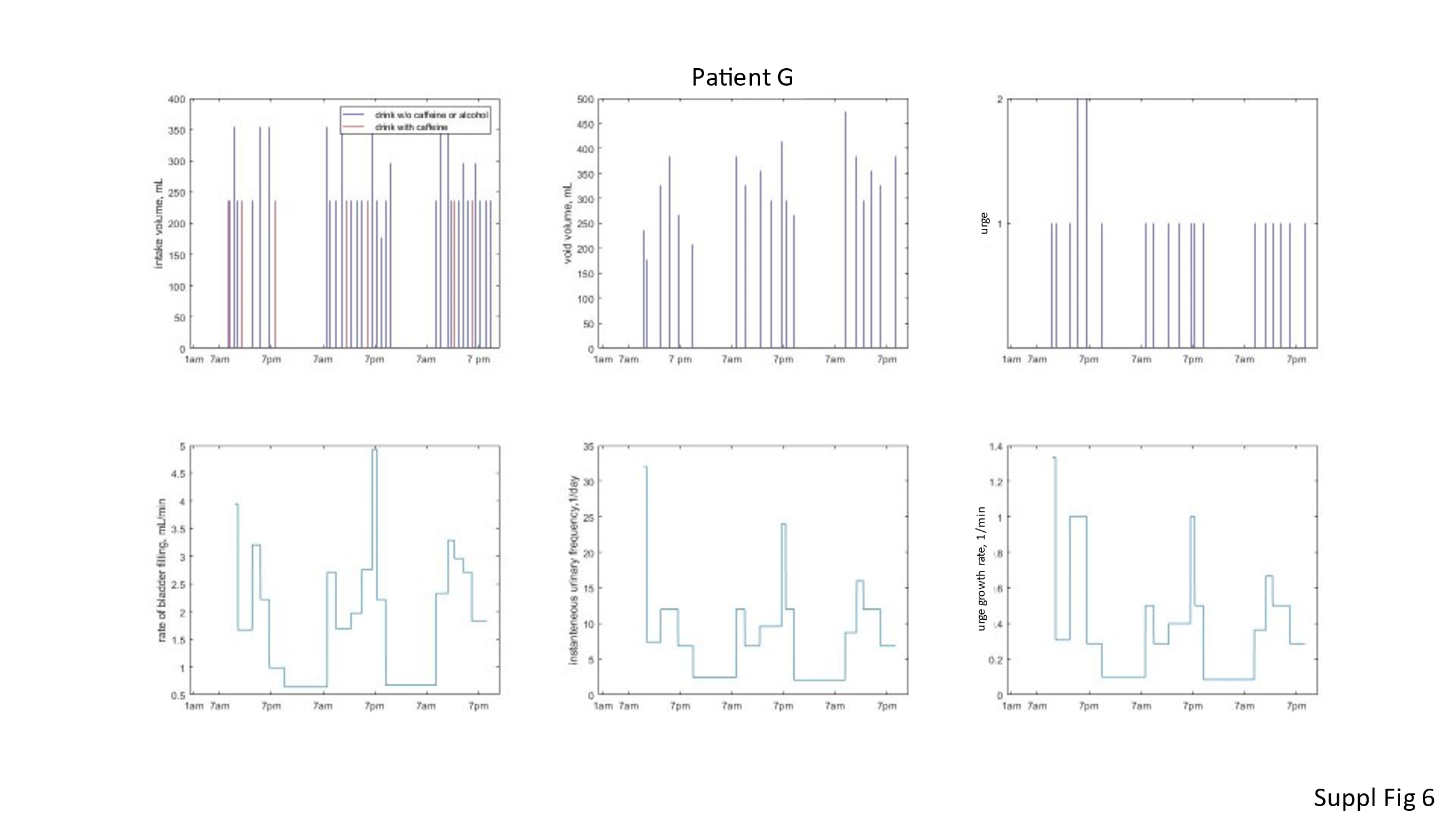

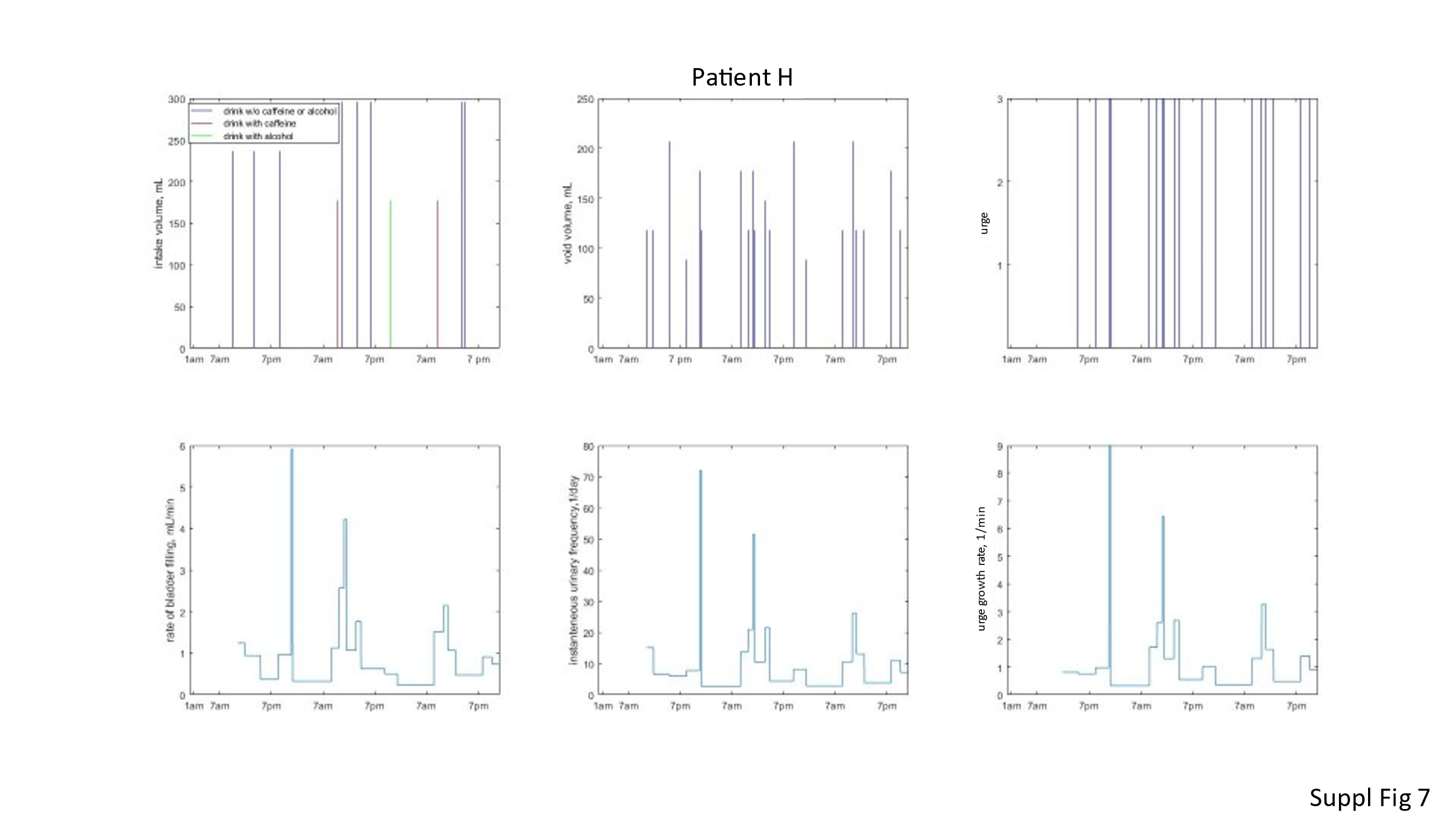

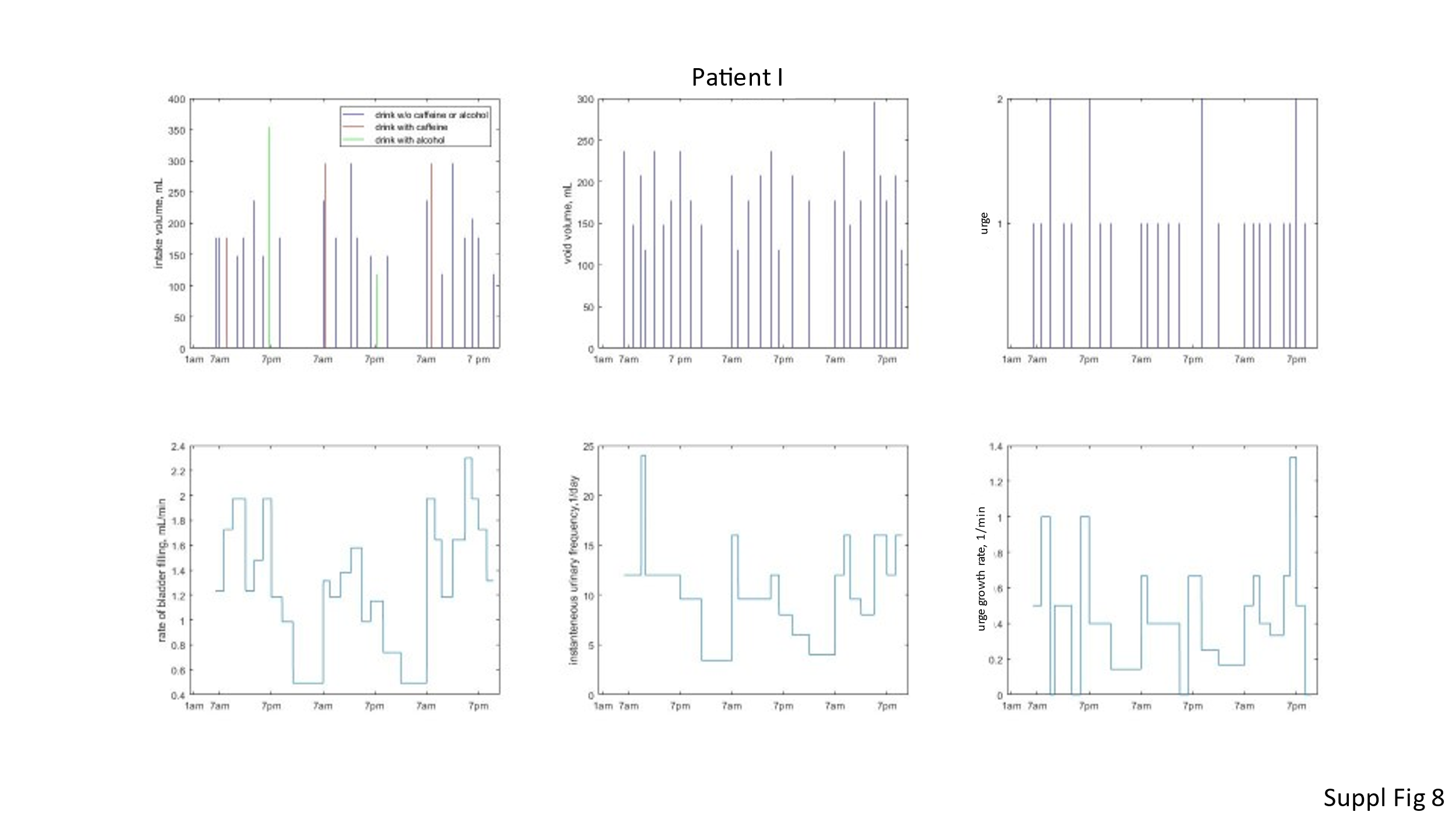

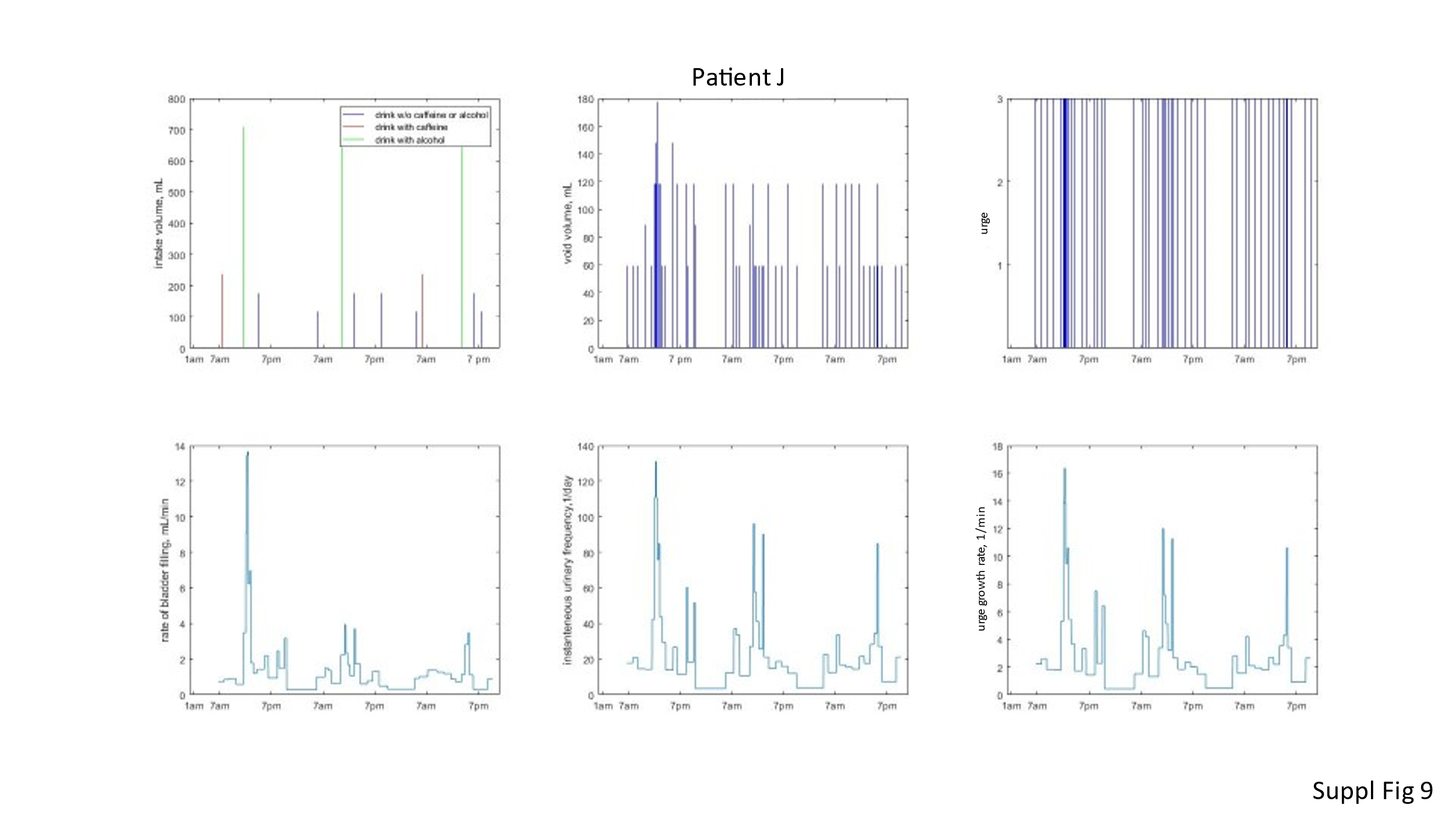

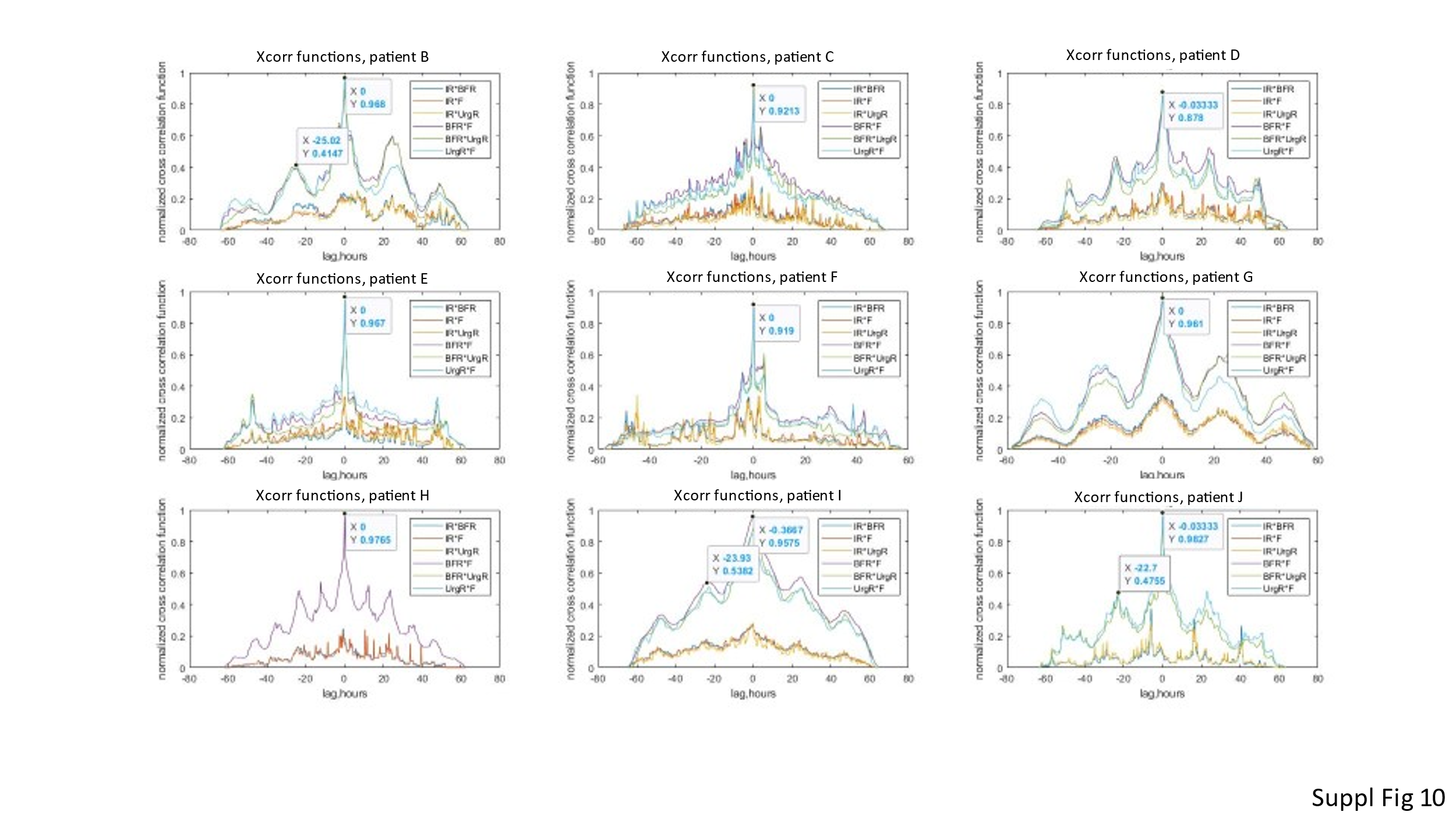

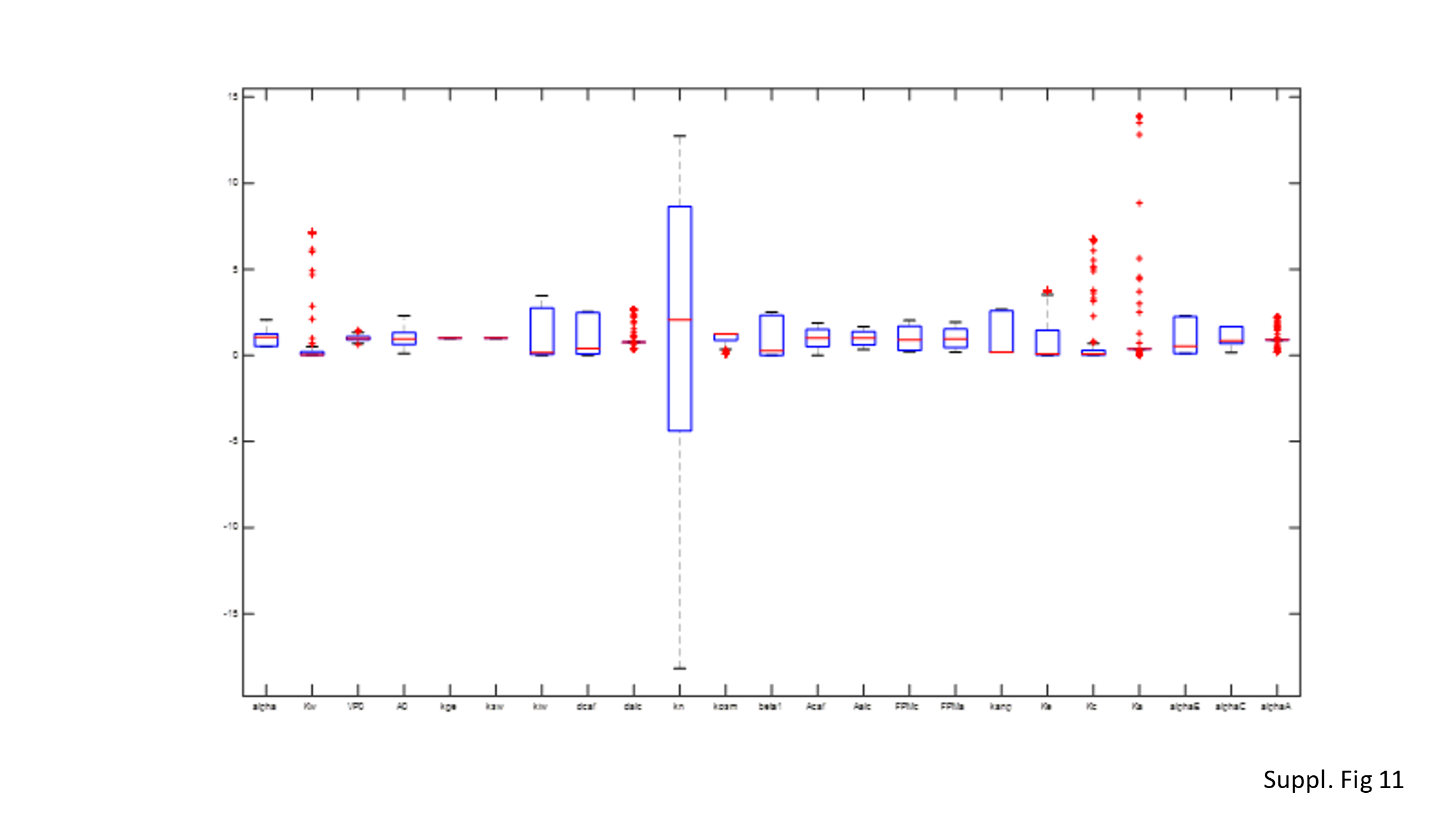
